# Artificial Intelligence in Depression – Medication Enhancement (AID-ME): A Cluster Randomized Trial of a Deep Learning Enabled Clinical Decision Support System for Personalized Depression Treatment Selection and Management

**DOI:** 10.1101/2024.06.13.24308884

**Authors:** David Benrimoh, Kate Whitmore, Maud Richard, Grace Golden, Kelly Perlman, Sara Jalali, Timothy Friesen, Youcef Barkat, Joseph Mehltretter, Robert Fratila, Caitrin Armstrong, Sonia Israel, Christina Popescu, Jordan F. Karp, Sagar V. Parikh, Shirin Golchi, Erica EM Moodie, Junwei Shen, Anthony J. Gifuni, Manuela Ferrari, Mamta Sapra, Stefan Kloiber, Georges-F. Pinard, Boadie W. Dunlop, Karl Looper, Mohini Ranganathan, Martin Enault, Serge Beaulieu, Soham Rej, Fanny Hersson-Edery, Warren Steiner, Alexandra Anacleto, Sabrina Qassim, Rebecca McGuire-Snieckus, Howard C. Margolese

## Abstract

**Background:** There has been increasing interest in the use of Artificial Intelligence (AI)-enabled clinical decision support systems (CDSS) for the personalization of major depressive disorder (MDD) treatment selection and management, but clinical studies are lacking. We tested whether a CDSS that combines an AI which predicts remission probabilities for individual antidepressants and a clinical algorithm based on treatment can improve MDD outcomes.

**Methods:** This was a multicenter, cluster randomized, patient-and-rater blinded and clinician-partially-blinded, active-controlled trial that recruited outpatient adults with moderate or greater severity MDD. All patients had access to a patient portal to complete questionnaires. Clinicians in the active group had access to the CDSS; clinicians in the active-control group received patient questionnaires; both groups received guideline training. Primary outcome was remission (<11 points on the Montgomery Asberg Depression Rating Scale (MADRS)) at study exit.

**Results:** 47 clinicians were recruited at 9 sites. Of 74 eligible patients, 61 patients completed a post-baseline MADRS and were analyzed. There were no differences in baseline MADRS (p = 0.153). There were more remitters in the active (n= 12, 28.6%) than in the active-control (0%) group (p = 0.012, Fisher’s exact). Of three serious adverse events, none were caused by the CDSS. Speed of improvement was higher in the Active than the Control group (1.26 vs. 0.37, p = 0.03).

**Conclusions:** While limited by sample size and the lack of primary care clinicians, these results demonstrate preliminary evidence that longitudinal use of an AI-CDSS can improve outcomes in moderate and greater severity MDD.

## Introduction

Major depressive disorder (MDD) is a leading cause of disability and socioeconomic burden^1^ impacting more than 300 million people worldwide^2^. Unfortunately, only a minority of patients will improve with the first treatment trial, and repeated treatment trials have diminishing probabilities of success^3^. Many patients undergo an arduous “trial and error” treatment selection approach, resulting in poorer outcomes, longer time in treatment, and greater patient and family burden^4^. To improve outcomes, it would be valuable to have a scalable point-of-care tool which can help personalize treatment choice without requiring expensive testing^5,6^.

There have been several efforts in recent years to use artificial intelligence (AI) to predict treatment outcomes in order to better match patients to specific treatments (see^7^. Most studies have differentiated between two treatments (e.g. two drugs) or treatment types (e.g. two types of psychotherapy), limiting clinical utility when many treatments are available. In addition, clinicians are often concerned about model bias and being able to interpret the outputs of AI predictive models which are often considered to be “black boxes”^8,9–10^ (providing predictions without clear reasoning). In addition, while improving predictions about treatment outcome may be helpful to personalize treatment, previous work has shown that treatments are often not managed in accordance with guidelines in terms of dosage and monitoring^11–13^. There is a need for a solution to both the treatment *selection* and the treatment *management* problems while integrating into existing clinical workflows^14^.

To address this, Aifred investigators developed the Aifred Clinical Decision Support System (hereinafter referred to as the CDSS). This is a digital platform which supports clinicians in the implementation of guidelines (2016 CANMAT depression guidelines^15^) and measurement-based care^15^ in order to solve the treatment *management* problem, and which includes an AI (deep learning) powered module to assist in baseline treatment *selection* by providing predicted probabilities of remission for 10 commonly used first line antidepressants and combinations of these. Extensive feasibility and ease of use testing of this CDSS was previously performed in both simulation center and *in vivo* feasibility studies^16–19^. With *in silico* testing demonstrating that the AI component should help improve remission rates^6,20–22^ and *in vivo* testing demonstrating that the platform was feasible, easy to use and likely safe^16–19^ the current study was undertaken with the main objective of determining the efficacy of the platform in improving depression treatment outcomes in patients with moderate to severe depression, as well as to assess platform safety.

## Methods

This study is reported as per the CONSORT-AI checklist ^23^ and was conducted in accordance with all relevant ethical regulations including the Declaration of Helsinki and the Tri-Council Policy Statement. The research ethics board of the Douglas Research Center gave ethical approval for this work and it was subsequently approved by central and local ethics review boards for each site. Written informed consent was obtained from all study participants. Full methods are available in the supplementary material.

### Design

The current study is a two-arm, cluster-randomized trial, with clinicians serving as the cluster. Clinicians rather than patients were randomized as they were the ones receiving the decision support intervention, and to avoid contamination ^24^. Patients entered the intervention arm of their treating clinician.

### Participants - Clinicians

Clinicians could include primary care doctors, psychiatrists, residents, nurse practitioners (with or without specialized mental health training), who saw at least one patient with depression per month, on average, before study start.

### Participants - Patients

Patient recruitment criteria were intended to be broad in order to replicate a naturalistic outpatient depression population with moderate to severe depression. Patients were recruited from the practices or hospital-based clinics of the participating clinicians. Inclusion criteria were as follows: 1) age 18 and over 2) diagnosed by their treating clinician with MDD using DSM-5 criteria ^25^ 3) MDD diagnosis confirmed via a blinded rater who completed the Mini Neuropsychiatric Interview (MINI) ^26^ and 4) at least moderate severity, as assessed by a blinded rater completing the Montgomery Asberg Depression Rating Scale (using a cutoff of 20) ^27^. Exclusion criteria were as follows: 1) age under age 18; 2) presence of bipolar disorder of any type; 3) inability or unwillingness to give informed consent; 4) inability to manage patient safely as an outpatient; 5) an active major depressive disorder was not the main condition being treated; and 6) an inability to use the tool (e.g. because of severe cognitive impairment). An active major depression meant that the depression, in the judgment of the treating clinician, required an initiation or a change in treatment. Psychiatric comorbidities (aside from bipolar disorder) were permitted.

### Settings

Eligible settings included any primary or secondary/tertiary public or private outpatient setting in the United States or Canada which provided outpatient care for patients presenting with MDD. A diverse array of sites joined the study. These included public sector psychiatric clinics in Canada and university-affiliated and Veteran’s Affairs mental health services in the U.S.

### Intervention - Aifred CDSS

The Aifred CDSS platform consists of the following elements. The *patient portal*, accessible by web browser or mobile phone application, allows patients to complete questionnaires, receive email reminders to complete questionnaires, visualize questionnaire scores and interpretations, and track treatments which they or their clinicians enter. The *clinician portal*, accessible by web browser, allows clinicians to see all the information patients enter, while at the same time giving them access to the *clinical algorithm* module. This module is a rule-based decision tree based on the CANMAT 2016 guidelines for depression treatment ^15^. This module presents the clinician with patient-specific, guideline derived information about treatment options based on the patient’s depression severity, change in depression severity over time (measured using the Patient Health Questionnaire (PHQ-9)^28^), and current treatments. The algorithm provides new information at each patient visit, based on patient progress. The AI component, which is separate from but housed within the algorithm module, is focused on assisting with treatment *selection* by generating remission probabilities for commonly used first line antidepressants based solely on baseline clinical and demographic details. The AI is a deep learning model trained on 9042 patients from depression treatment trials^29,30^; the AI, the data it intakes, and the report it provides clinicians is described further in the supplementary materials.

### Intervention - Patients

All patients received access to the patient portal of the CDSS. Patients remained in the study for 12 weeks from their first treatment visit, at week 2, weeks 4-6, week 8 and week 12.

### Intervention - Clinicians

There were two intervention groups: an Active group and an Active-Control group^31,32^. Clinicians in the Active-Control were provided with the results of questionnaires patients completed as well as training on the CANMAT guidelines^15^. Active group clinicians received guideline training, and were provided with full access to and training on the clinician portal of the Aifred CDSS. As all raw data provided to clinicians was the same in the Active and Active-Control groups, the only group differences consisted of the provision of the data processed by the clinical algorithm and AI model to the Active group. Clinicians were not required to use the information they were provided in any specific manner.

### Measures

At baseline, the blinded rater assessed the Mini International Neuropsychiatric Interview (MINI) and Montgomery–Åsberg Depression Rating Scale (MADRS). Patients were asked to complete a Patient Health Questionnaire (PHQ-9) and General Anxiety Disorder (GAD-7) weekly once they had accounts on the CDSS. The MADRS was administered by the blinded rater at screening, visit 3 (weeks 4-6 of treatment), visit 4 (week 8) and visit 5 (week 12). Trained study staff also administered the Brief Adherence Rating Scale (BARS) after every visit to assess treatment adherence ^33^. Further measures are detailed in the supplementary material.

### Outcomes

The pre-specified primary outcome of the study was remission of depressive symptoms, defined as a score of <11 ^34,35^ on the MADRS at study exit for those patients with at least two MADRS scores. Safety outcomes included an examination of the nature and number of adverse and serious adverse events in each group. Secondary outcomes included response (50% reduction in symptoms) on the MADRS, rate of change of the MADRS score, and medication adherence using the BARS score.

### Blinding

#### Patients

Patients were fully blinded to group assignment. They were told that they were entering a study where they would be using a new digital technology to provide information to their clinician and that there were two groups, but the nature of the groups was not revealed.

#### Clinicians

Clinicians were aware of their group assignments as they were the ones receiving the AI predictions. Clinicians were partially blinded to reduce expectation bias^36^: they were not told the study endpoints, and they were not informed of the expected effect sizes of the interventions.

#### Raters

Raters who collected the primary outcome (MADRS) and conducted the MINI were blind to group allocation.

### Statistical Analysis

Outcome data were analyzed, as prespecified in the Statistical Analysis Plan, on an intent-to-treat basis for patients who had at least two ratings of the MADRS (the Analysis set). Safety data were analyzed for the Safety population, pre-specified as all patients who attended at least the first treatment visit. Demographic and baseline clinical data are presented in Table 1. The primary outcome (MADRS remission) was assessed using a Fisher’s exact test. Secondary outcomes were compared using one-way ANOVAs, and proportions were compared using two-sided X^2^ or two-sided Fisher’s exact tests, as appropriate. Further pre-specified analyses are detailed in the Supplementary Material.

**Table 1:**
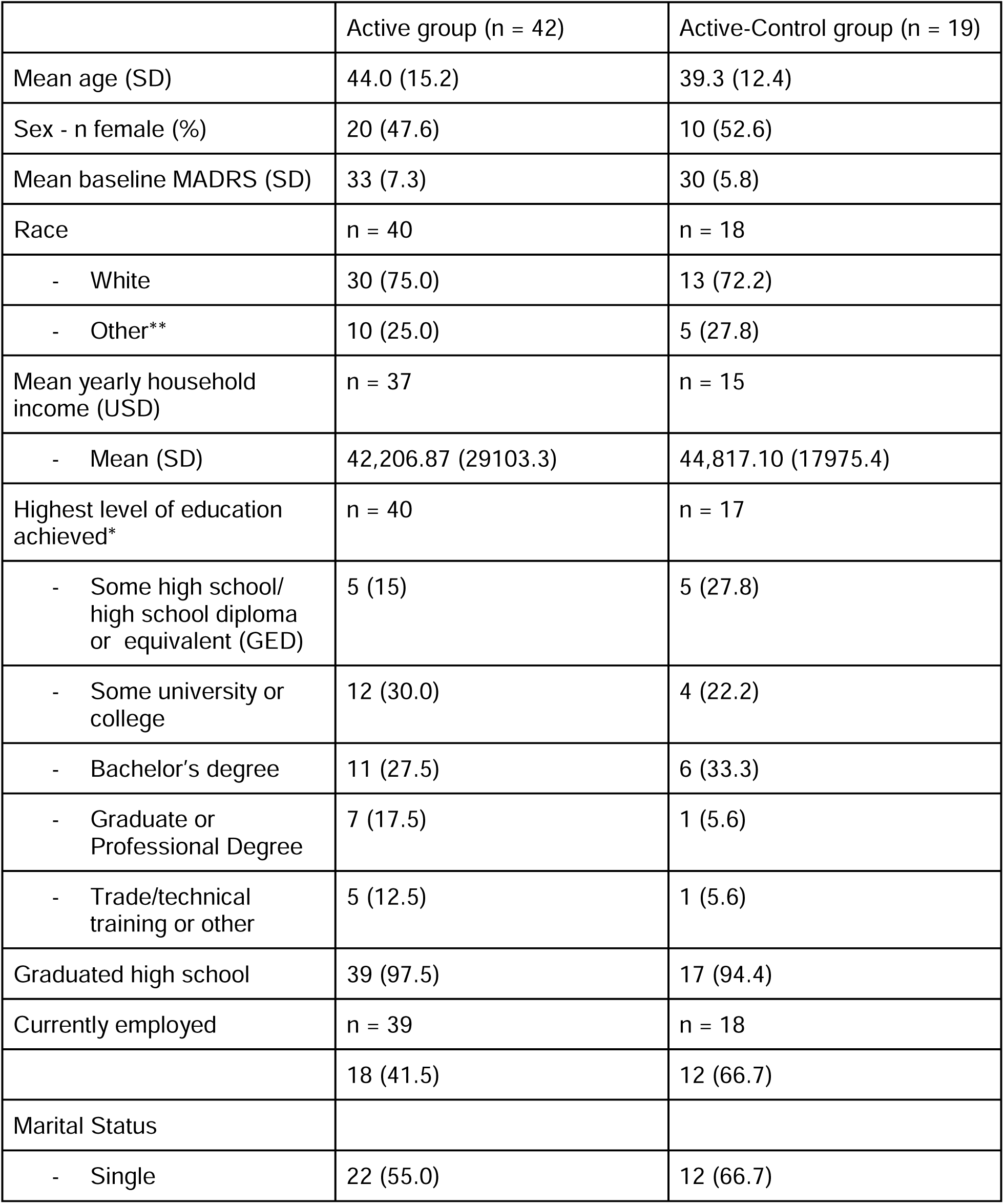
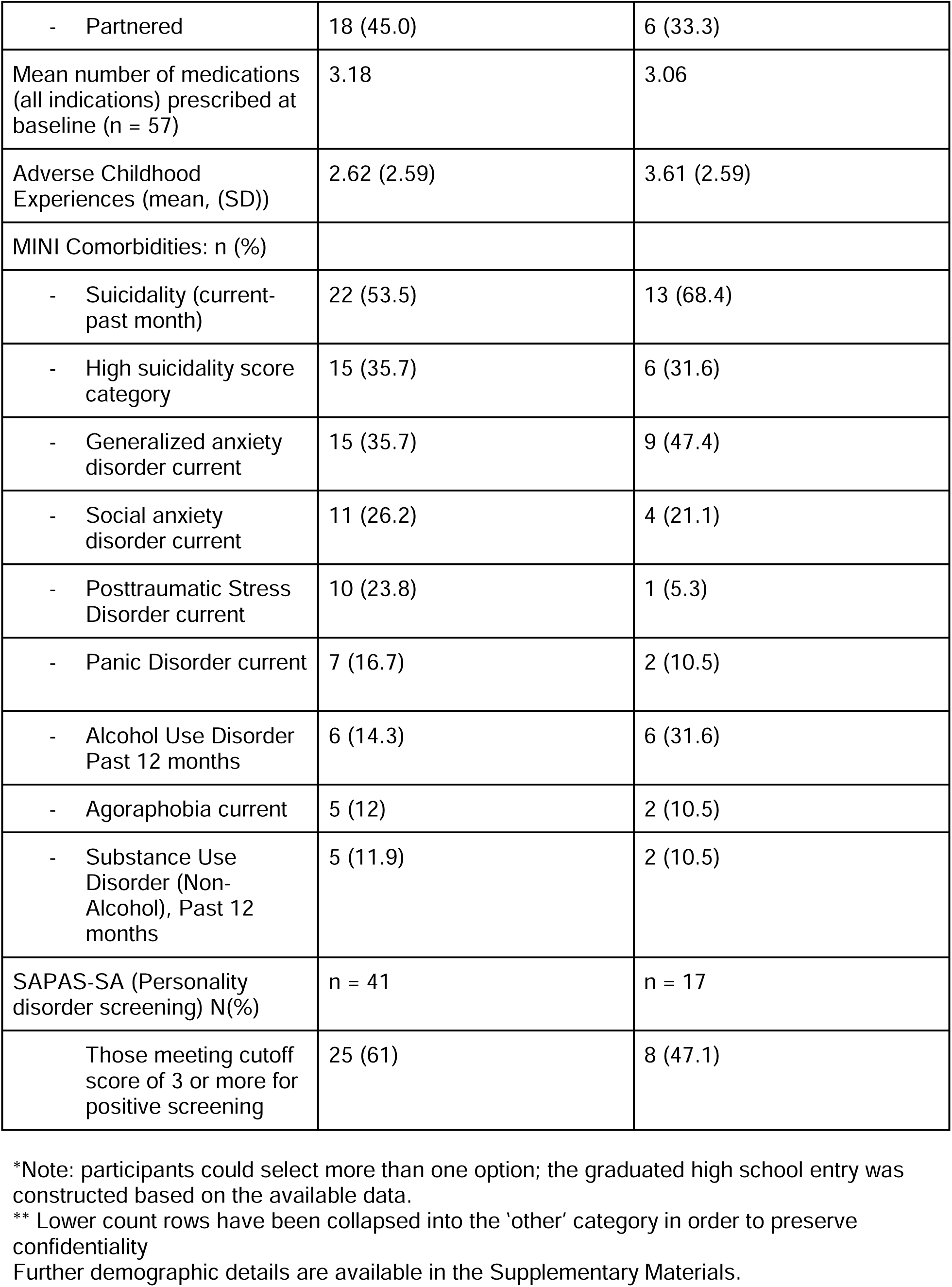
Baseline Clinical and Demographic Characteristics per Group.

### Early Study Termination

Unfortunately, due to lack of funding caused by delays related to the COVID-19 pandemic, the study was terminated early.

## Results

### Sites

Sites were located in Canada (5) and the United States (4) and included U.S.Veterans Affairs hospitals and mood disorders programs in university-affiliated psychiatric departments.

### Recruitment - Clinicians

Of the 47 clinicians recruited who were cleared to recruit patients prior to early study termination, 25 were randomized to the Active group and 22 to the Active-Control group. 27 clinicians recruited at least one patient (57%); 16 in the Active group (64%) and 11 in the Active-Control group (50%).

### Recruitment and Dropout - Patients

Patients were recruited between 2022-06-15 and 2023-11-16, a total of 17 months. The study was terminated early because of lack of funds due to delays in study initiation related to COVID-19. Recruitment and dropout are summarized in the CONSORT diagram **(Figure 1)**. Of the 74 eligible patients after screening and enrollment, 61 had at least 2 MADRS available, forming the Analysis set (n = 42 Active, n = 19 Active-Control). The groups did not differ in terms of 12 week completion (Active = 36/53 (68%); Active-Control = 18/21 (86%) (p = 0.15, Fisher’s exact test)).

**Figure 1.**
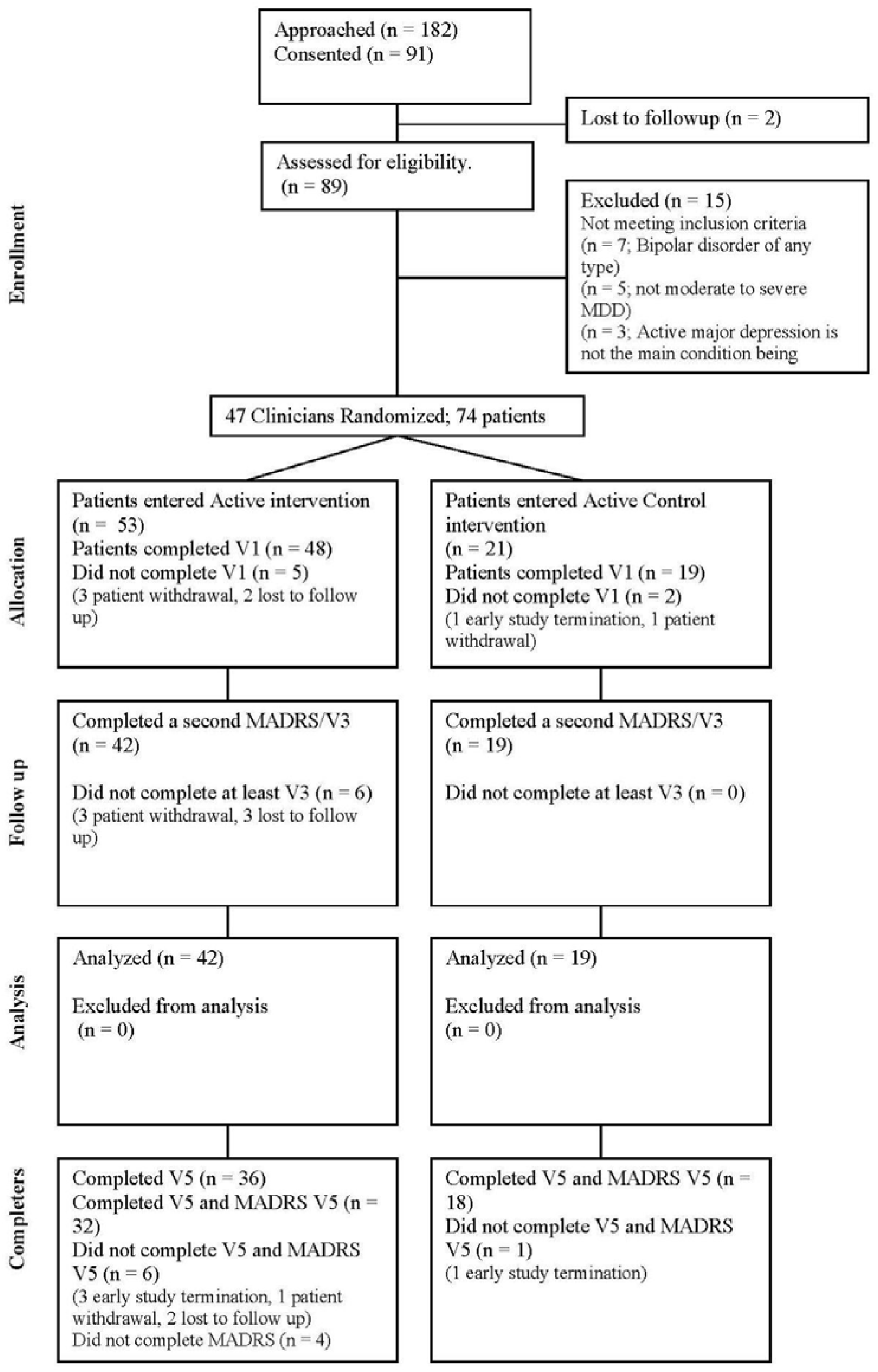
Patient Consort Diagram

### Demographics and baseline clinical characteristics

Intervention and Active-Control groups did not differ with regards to important clinical and demographic characteristics (Table 1). The group had substantial chronicity of illness with 33 Active patients (78.6%) and 13 Active-Control patients (68.4%) having recurrent MDD. Further details are available in the Supplementary Material.

### Treatment Outcome - Remission at Study Exit

On the primary outcome, remission, there were significantly more remitters in the Active (n = 12/42 (28.6%)) than in the Active-Control (0/19, (0%)) group (p = 0.01, Fisher’s exact test). Outcomes are summarized in table 2.

**Table 2:**
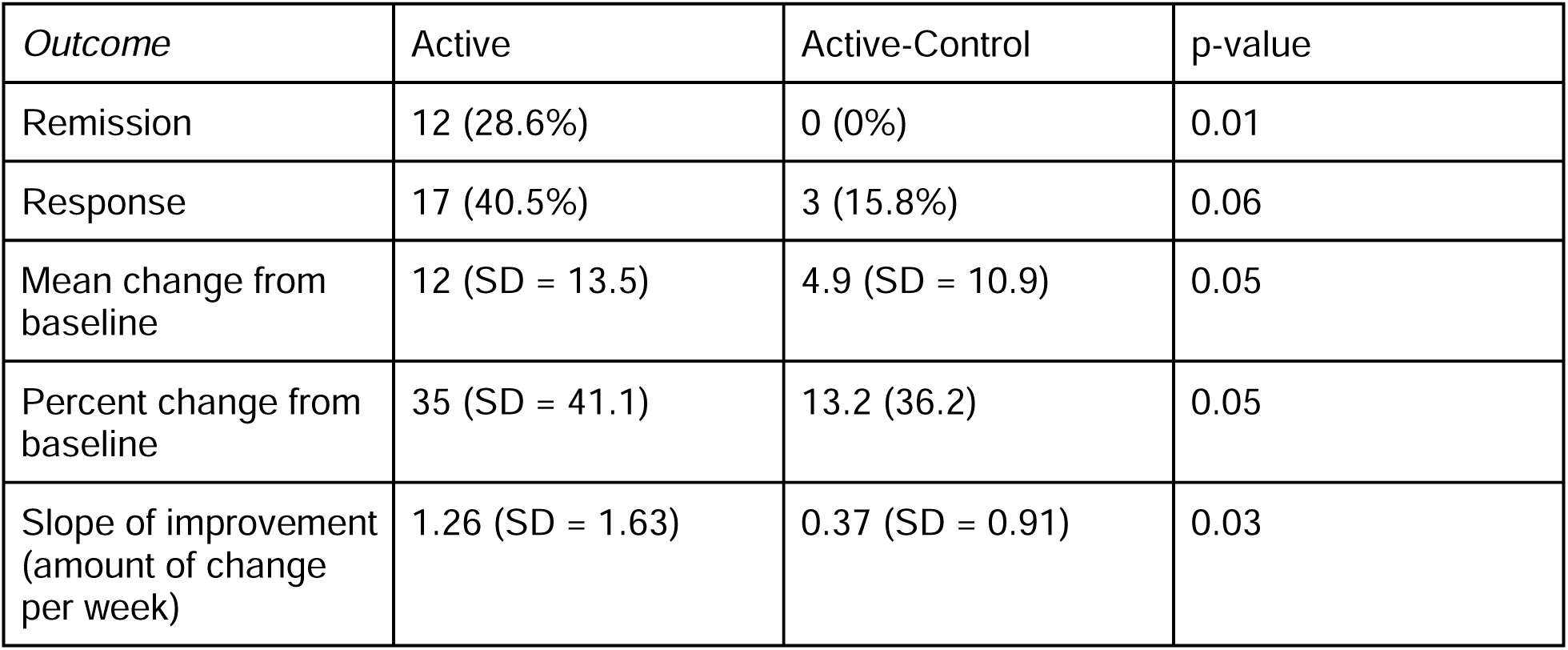
Summary of Outcomes.

### Treatment Outcome - Response and Change from Baseline

With respect to treatment response (defined as a 50% or greater decrease in total MADRS score between screening and study exit), 17 Active patients (40.5%) and 3 Active-Control patients (15.8%) responded at study exit. This was a large numerical difference, however, it did not reach significance (X^2^ = 3.6, p = 0.06). The proportion of responders was not significantly different at visit 3 or 5, but was significantly different at visit 4 (p = 0.04, Fisher’s exact test).

With respect to change from baseline to study exit, patients in the Active group experienced a mean 12.0 point improvement in MADRS score (SD = 13.5) while those in the Active-Control group experienced a change of 4.9 (SD = 10.9). Again, while a large numerical difference, it did not reach significance (F(1) = 4.006; p = 0.05). This corresponds to a between-group difference of 7.1 points, which exceeds the accepted threshold for a minimum clinically important difference between groups on the MADRS ^37^.

In terms of percent change of MADRS score from baseline to score at study exit, the Active group experienced a mean 35% change (SD = 41.1) and the Active-Control group experienced a mean 13.2% change (SD = 36.2); this difference was again numerically large but non-significant (F(1) = 3.95, p = 0.05).

### Treatment outcome - Rate of Change

Investigators observed a significantly faster rate of improvement in MADRS score (change in MADRS score divided by treatment weeks a patient spent in the study) in the Active group compared to Active-Control. The mean change in total MADRS score per week in the Active group was 1.26 points (SD = 1.63); in the Active-Control group this was 0.37 points per week (SD = 0.91), (F(1) = 4.99; p = 0.03; eta-squared = 0.08, 95% CI [0,0.23], ANOVA). See Figure 2.

**Figure 2:**
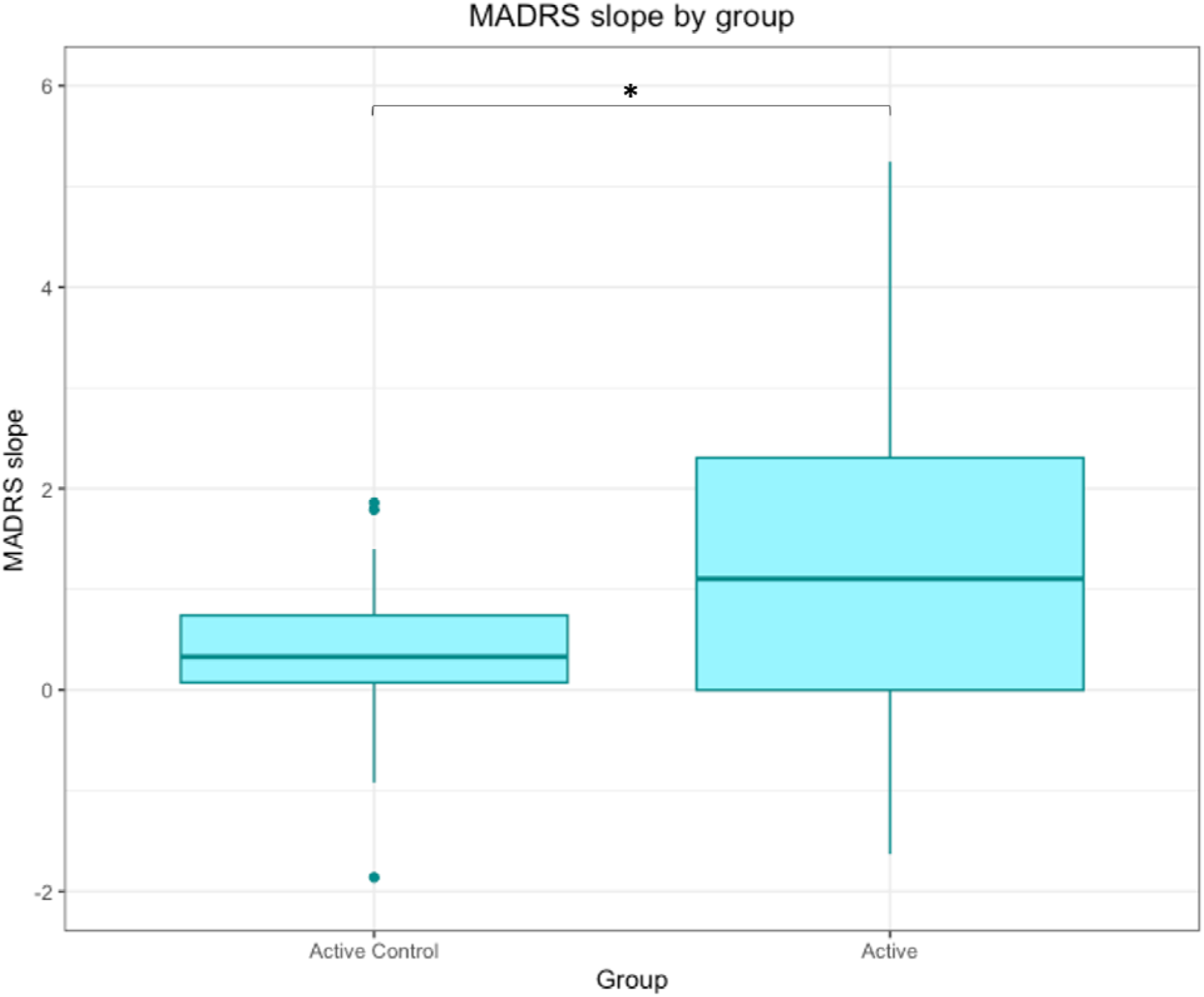
MADRS Slope by Group - bar plot displaying significantly different mean MADRS slope per group (F(1) = 4.99; p = 0.03; eta-squared = 0.08, 95% CI [0,0.23], ANOVA) Error bars represent standard deviation.

### Treatment Adherence

Patients reported high levels of treatment adherence across visits in the study on the BARS questionnaire (mean 96.4% adherence, SD = 13 in Active; 95% adherence, SD = 10.9 Active-Control, F(1) = 0.76, p = 0.40).

### Safety Outcomes

With respect to safety, adverse event rates (e.g., medication side effects) and serious adverse event rates (e.g., hospitalizations) were examined in the Safety population (48 Active, 19 Active Control). 89 adverse events were reported for the Active group, a rate of 1.9 events per patient, compared to 51 events (2.7 events/patient) in the Active-Control group. There were 3 serious adverse events in the Active group, and none in the Active-Control group. All three events were determined to have been unrelated to the CDSS by the site’s primary investigator. Further details can be found in the Supplementary Material.

### Patient Engagement

We examined PHQ-9 completion rates during the 12 treatment weeks (Fig. 3). The total PHQ-9 completion rate was 70% (67% in active, 77% in the Active-Control group). These completion rates are in line with our previous feasibility study^18,19^.

**Figure 3:**
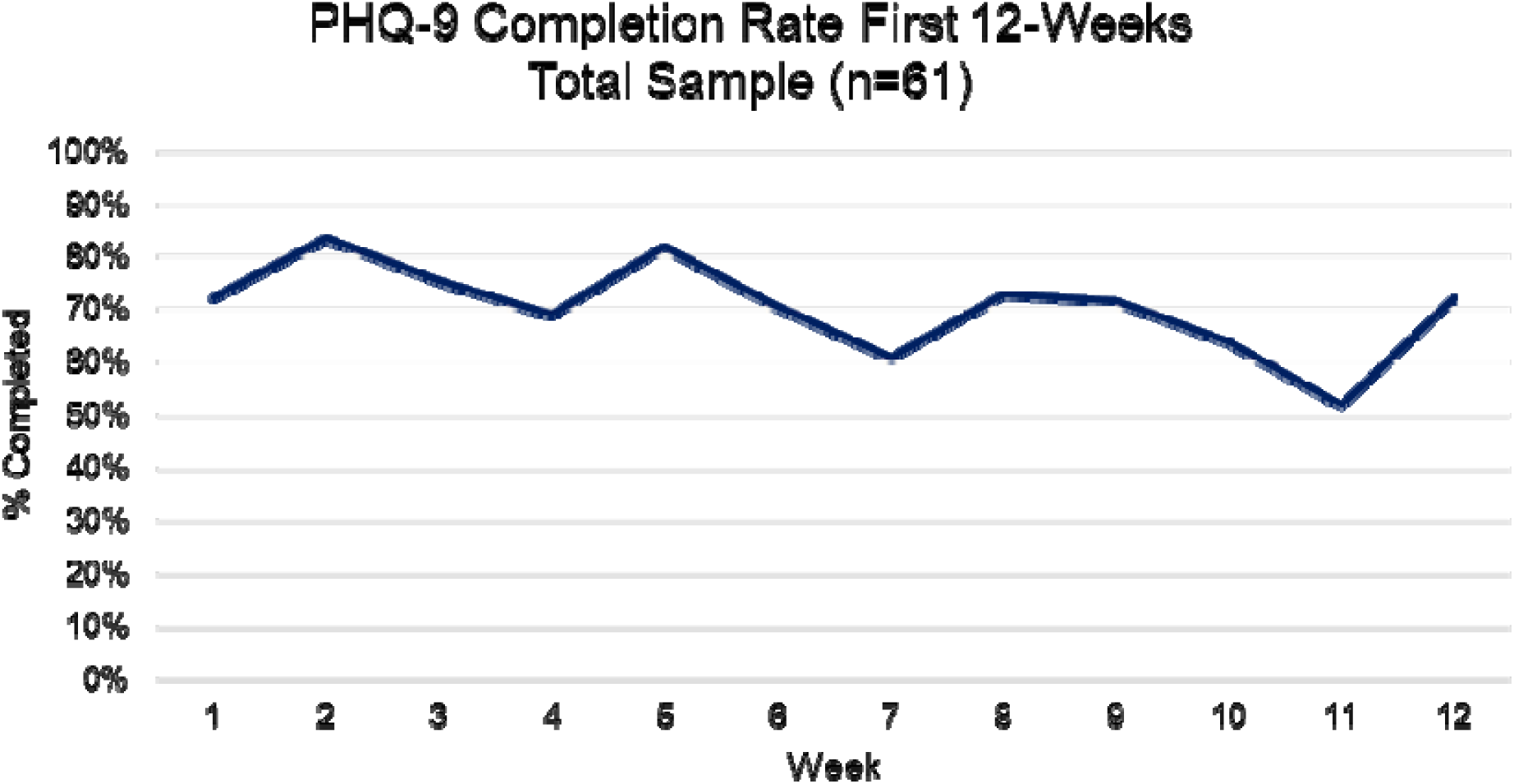
PHQ-9 completion rates for the 12 treatment weeks.

### Clinician Engagement

Over the course of the study, 81.25% of doctors in the Active group who recruited patients accessed the CDSS at least twice (e.g. at least once after training); 81.25% accessed the treatment algorithm at least twice (e.g. at least once after training), and 81.25% accessed the AI results at least once (note: AI results were not available during training). Per visit, we report the number of Active clinicians who accessed the application, and of these, the number that accessed the treatment algorithm and the AI results (Table 3).

**Table 3:**
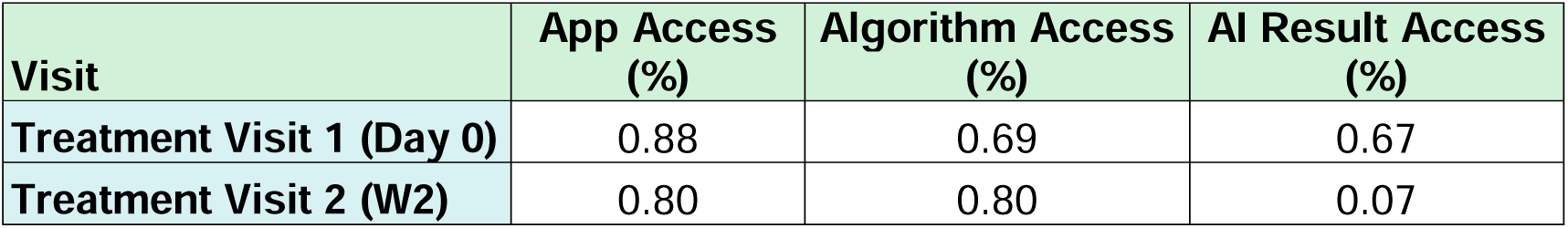
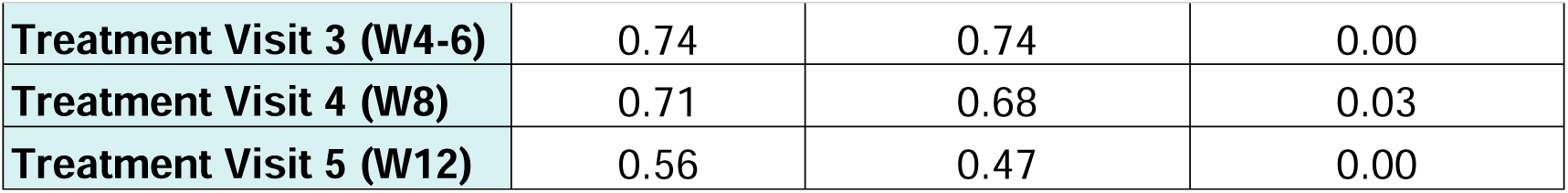
Clinician Platform Usage by Visit. App access = proportion of clinicians in the active group who logged into the app at each visit; Algorithm Access = proportion of clinicians who went beyond logging in and access the clinical algorithm module; AI Result Access = proportion of clinicians who accessed the AI results at the end of one session of the clinical algorithm, which was intended to occur at Visit 1 or 2 (one access later in treatment was a result of an error made by a clinician and logged as a protocol deviation).

## Discussion

This study is the first of its kind in mental healthcare to integrate an AI-powered CDSS in a longitudinal fashion to assist clinicians in making more effective clinical decisions about treatment selection and management while providing patients with more information about their own symptoms and trajectories. These initial results suggest that this CDSS is effective in improving remission rates (28.6% in the Active group compared to no remitters in the Active-Control group) and the rate of symptom improvement in adult patients with moderate or greater severity depression. In addition, engagement with the platform by both clinicians and patients was high throughout the study, and no adverse events were linked to the CDSS.

Despite both groups having the same baseline depression severity, similar clinical profiles, and the same treatment options permitted, patients in the Active group had significantly improved remission rates and more rapid improvement. Given this, we suggest that the CDSS may had a positive impact on clinical decision-making, demonstrating the potential for a CDSS which can organize information, present it at the clinically appropriate time at the point of care, and provide personalized treatment outcome predictions to significantly improve outcomes^38–40^. Further discussion of potential mechanisms of action for the CDSS is presented in the Supplementary Material.

This study has several strengths. The first is a design that intended to replicate realistic use of the CDSS, where clinicians and patients were not required to use the platform or adhere to it in any particular manner. Findings of improved outcomes in the Active group may therefore approximate benefits which would be derived in real clinical practice. The second keystrength is the comparison of the CDSS with an Active-Control group that approximated realistic best practices in-clinic today, suggesting that the CDSS may be able to improve outcomes over and above these best practices. Finally, the CDSS was simple to introduce into a diverse array of clinical environments and had reasoable patient and clinician engagement, which increases its potential for rapid adoption.

This study also has several limitations. The first is the smaller than intended sample size. This limits the power of subgroup and secondary analyses. It is reassuring, however, that significant clinical benefit was observed in line with the *a priori* estimated effect size. As discussed in the methods and in investigators’ companion paper <u>(Perlman et al. 2024)</u>, the AI model used in this study was limited to providing initial treatment outcome predictions and could not adapt to treatment failure, and it had a preference for escitalopram being predicted as the treatment most likely to be effective, while providing more variable predictions for the other medications (see^30^ for detailed discussion). In addition, first line treatments such as psychotherapy, vortioxetine and mirtazapine monotherapy could not be included in the AI model due to lack of data, though these were present in the treatment algorithm. Future versions of the model will continue to be improved by more and more diverse data, which will likely continue to improve the performance of the platform. Another limitation is the fact that the CDSS is a composite intervention, consisting of measurement based care, a rule-based algorithm, and the AI model. It would be important to be able to identify, in future work, which elements of the intervention are most responsible for the clinical improvements seen. Another limitation is the imbalance in the number of patients recruited into the Active and Active-Control groups. This may speak to the interest that Active group clinicians may have had in using the tool with patients.

## Conclusions

In this paper, we have shown preliminary data indicating the clinical effectiveness and safety, in a cluster-randomized study, of an innovative AI-powered CDSS to support clinical decision making in the treatment of adult patients with MDD of moderate and greater severity. Use of this and similar systems, which could be implemented rapidly into clinical practice with minimal training, has significant potential to improve the effectiveness and speed of treatment for MDD. Future work on CDSS systems like this which are intended to be integrated into longitudinal care may benefit from the study methods discussed here. Replication of this work in larger samples by independent investigators is also needed. Future work could also be directed at further expanding and improving the AI model implemented in the intervention, potentially using data collected during real-world use of the CDSS.

Registration at clinicaltrials.gov: NCT04655924

The full protocol and statistical analysis plan are available in the Supplementary Material.

### Funding

Funding was provided by Aifred Health; MEDTEQ FSISSS; Bell Let’s Talk and Brain Canada; Investissement Quebec; the Quebec Ministry of Economy and Innovation; the Mindstrong Foundation at the Jewish General Hospital; and the McGill Industry and Partnership grant. The study was co-designed and supervised by HM and Aifred Health. No other funder had a role in the development or reporting of this research.

## Supporting information

CONSORT AI Checklist

Protocol

Final SAP

## DISCLOSURES

### Declaration of Competing Interest

The authors declare the following financial interests/personal relationships which may be considered as potential competing interests:

DB, KW, MR, GG, KP, JM, RF, CA, SI, CP, AA, and SQ were employees and/or shareholders of Aifred Health Inc. and supported this research in the context of their work for Aifred Health. JFK, SP are members of Aifred Health’s scientific advisory board. SP has received honoraria from Aifred Health. DB, SG, EEMM, and SR receive a salary award from the Fonds de recherche du Québec – Santé (FRQS). EEMM is a Canada Research Chair (Tier 1) in Statistical Methods for Precision Medicine. BWD has received research support from Boehringer Ingelheim, Compass Pathways, Intra-Cellular Therapies, NIMH, Otsuka, Usona Institute, and has served as a consultant for Biohaven, Cerebral Therapeutics, Myriad Neuroscience, and Otsuka. SR receives grant funding from Mitacs (for a graduate student), is on a steering committee for Abbvie, and owns shares of Aifred Health. SK reports grants from the Labatt Family Innovation Fund in Brain Health (Department of Psychiatry, University of Toronto), the Max Bell Foundation, the Canadian Centre on Substance Use and Addiction, the Centre for Addiction and Mental Health Discovery Fund, the Ontario Ministry of Health and Long-Term Care (MOHLTC), the Canadian Institutes of Health Research (CIHR), and the International OCF Foundation (IOCDF). SK received honorarium for consultation from EmpowerPharm. JFK has been provided options in Aifred Health. HCM has received honoraria, sponsorship, or grants for his participation as a consultant, advisory committee member, and/or as a speaker at educational events for AbbVie, HLS Therapeutics Inc., Janssen, Lundbeck, Otsuka, Newron, Sunovion and Teva. He has received grants and/or research support from the MGH Hospital Foundation, SyneuRX and Aifred Health. SJ, TF, and YB recieved honoraria from Aifred Health for working on this paper. All other authors report no relevant conflicts.

### Code availability

The methods used to develop the AI platform are available in previous publications; however the final code for the CDSS and final trained model are not publicly available at this time. Previous versions of the pipeline used to construct the model are available here (https://github.com/Aifred-Health/VulcanAI) and a full version of the model trained using a pharmaceutical dataset is available here: (https://github.com/Aifred-Health/pharma_research_model).

### Data availability

Data requests will be considered by the study sponsor for non-commercial applications, on the condition that this publication is referenced in publications using the dataset. Requests for data access should be directed to and will be responded to within 30 days. Data requested will be transferred in a de-identified format using patient ID numbers.

## Acknowledgements

We thank patients, physicians, and local staff for participating in this study.

## Authorship statement

DB, SI, KP, MR, HM, conceived the study and revised the paper. DB conducted and supervised the study and analyses, and wrote the first draft of the paper. KW, MR, GG, SI, KP, CP, AA, and SQ assistant with data collection and manuscript review. KW, MR, GG, KP, SJ, TF, YB, RF and CP assisted with analysis and paper revision. JM, RF, CA, KP, and DB produced the machine learning model used in the study and revised the paper. JK, SP, SG, EM, ME, SR, FHE, and HM served on the steering committee and revised the paper. SG and EM served as statisticians who oversaw sample size calculations carried out by JS and reviewed the statistical analysis plan, which was approved by DB, MR and HM. AG, MF, SP, MS, SK, GP, BD, KL, MR, served as site primary investigators and revised the paper. SB, WS served on the data safety monitoring board and revised the paper. RMS provided the STAR questionnaire and revised the paper. HM provided supervision for the study, the analysis, and revised the paper.

## Notes

### Clinical Trial

NCT04655924

### Funding Statement

Funding was provided by Aifred Health; MEDTEQ FSISSS; Bell Lets Talk and Brain Canada; Investissement Quebec; the Quebec Ministry of Economy and Innovation; the Mindstrong Foundation at the Jewish General Hospital; and the McGill Industry and Partnership grant. The study was co-designed and supervised by HM and Aifred Health. No other funder had a role in the development or reporting of this research.

### Author Declarations

The study was conducted in accordance with all relevant ethical regulations including the Declaration of Helsinki and the Tri-Council Policy Statement. The research ethics board of the Douglas Research Center gave ethical approval for this work and it was subsequently approved by central and local ethics review boards for each site. The study was conducted in accordance with Good Clinical Practice. Written informed consent was obtained from all study participants.

### Summary of Updates

Submitted with updated supplementary materials.

