## Supplementary material for "Artificial Intelligence in Depression – Medication Enhancement (AID-ME): A Cluster Randomized Trial of a Deep Learning Enabled Clinical Decision Support System for Personalized Depression Treatment Selection and Management": Protocol

DocuSign Envelope ID: 3DB15B31-6E89-4135-81C3-F2DBD9C216BB

### A Randomized, Patient and Rater Blinded, Active-Controlled Trial of a Hybrid-Classic/Machine-Learning Enabled Clinical Decision Aid for Personalized and Individualized Pharmacological Depression Treatment Selection

**Protocol Number:** **IUSMD 18-04**

**Version:** 4.0
**Date:** 19 January 2022
Medical Monitor: David Benrimoh, M.D., C.M., M.Sc.
Chief Science Officer

Sponsor: Aifred Health Inc.
304-1600 rue Notre-Dame O
Montreal, QC H3J 1M1
Canada

DocuSign Envelope ID: 3DB15B31-6E89-4135-81C3-F2DBD9C216BB

**AID-ME: Artificial Intelligence in Depression – Medication Enhancement: A Randomized, Patient and Rater Blinded, Active-Controlled Trial of a Hybrid-Classic/Machine-Learning Enabled Clinical Decision Aid for Personalized and Individualized Pharmacological Depression Treatment Selection**

**Global Principal Investigator:** Dr. Howard C. Margolese, MD.,CM., MSc, FRCPC
Allan Memorial Institute, McGill University Health Centre (MUHC),
1025 Pine Ave W, Montreal, QC H3A 1A1, Canada


**Supported by:**
Aifred Health Inc.
IRAP Innovation Grant
McGill University Industry Partnership Grant

**Study Intervention Provided By:**
Aifred Health Inc.
www.aifredhealth.com
304-1600 rue Notre-Dame O
Montreal, QC H3J 1M1
Canada

**Protocol Revision History**

| Version Number | Version Date | Summary of Revisions Made |
|---|---|---|
| 1.0 | 27 January 2018 | N/A |
| 1.1 | 1 February 2018 | Revisions based on REB/IRB comments |
| 1.2 | 4 May 2018 | Revisions based on REB/IRB comments |
| 1.3 | 20 April 2020 | Added funding, corrected data retention time to 25 years, clarified recruitment process, revised patient evaluation schedule, fixed formatting |
| 1.4 | 6 August 2020 | Protocol formatting fixed<br>Sites added<br>Continuation study portion to be moved to another protocol<br>Added MADRS and MINI assessments for diagnostic clarification<br>Added that the study will be conducted in a patient-and-rater blinded manner, with some elements of physician blinding<br>Updated the name of the study<br>Updated sites<br>Updated primary investigator<br>Simplified and updated assessment schedule<br>Added week 8 visit<br>Removed biomarkers from this study<br>Clarify randomization for physicians and the fact that the study is cluster randomized<br>Added virtual visits because of COVID-19 |
| 1.5 | 24 October 2020 | Revisions based on REB comments following conditional approval of protocol version 1.4, including:<br>Update of study name<br>Declaration of interests<br>Plan to deal with a participant at risk of suicide or self-harm<br>Clarification of e-consent process<br>Clarification of Inclusion Criteria No. 4 for patients in sections 5.1.2<br>Clarification of ethical considerations<br>Compensation for patients and physicians for |

DocuSign Envelope ID: 3DB15B31-6E89-4135-81C3-F2DBD9C216BB

| | | |
|---|---|---|
| | | end of study interview<br>Other clarifications include:<br>Addition of Protocol Approval Page<br>Addition of Protocol Signature Page<br>Clarification on the number of physicians and patients and randomization process<br>Clarification of study objectives and hypotheses |
| 2.0 | 23 June 2021 | Make physical exams "as clinically indicated" in the context of the pandemic<br>Clarify age of eligible patients to ≥18<br>Update study initiation and completion dates<br>Deletion of references to a continuation study<br>Minor formatting corrections<br>Populated the list of abbreviations with a few missing terms<br>Additional details added regarding the electronic data capture process<br>Clarification regarding custom questionnaire and QIDS-SR16<br>Additional details added regarding the study steering committee and Data Safety Monitoring Board (DSMB)<br>Update Table of Contents page numbers<br>Harmonized all iterations of the study title<br>Updated information regarding the messaging feature of the CDA |
| 3.0 | 10 December 2021 | Updated Global PI from Dr Turecki to Dr Margolese.<br>Updated the description of the custom questionnaire including the specifications that the custom questionnaire is responded to in two parts: a clinician part at visit 1 and a patient part in the 2 weeks prior to visit 1. In addition, the interview guides the clinician will use to train for the clinician part are specified. |
| 4.0 | 19 January 2022 | Updated Aifred address from the Guy street address to the Notre-Dame O address.<br>Updated study duration in section 1.2<br>Added new section 12.0 about Circle of Care |

| | | |
|---|---|---|
| | | feature and updated the numbering of the remaining sections |

TABLE OF CONTENTS

*Page*

Protocol Revision History 3
LIST OF ABBREVIATIONS 9
DISCLOSURE STATEMENT 11
1. SYNOPSIS 14
1.1 Administrative Structure of the Study 14
1.2 Participating Study Sites and Study Duration 14
1.3 Study Title 15
1.4 Objectives 15
1.5 Design and Outcomes 15
1.6 Interventions and Duration 16
1.7 Sample Size and Population 17
2. BACKGROUND AND RATIONALE 18
2.1 Background 18
2.2 Study Purpose and Rationale 19
2.2.1 General Rationale 19
2.2.2 Study Specific Rationale 20
3. STUDY OBJECTIVES 20
3.1 Primary Objective 20
3.2 Secondary Objectives 21
4. STUDY DESIGN 22
5. SELECTION AND ENROLLMENT OF PARTICIPANTS 25
5.1 Inclusion Criteria 25
5.1.1 Inclusion Criteria for Physicians 25
5.1.2 Inclusion Criteria for Patients 25
5.2 Exclusion Criteria 26
5.2.1 Exclusion Criteria for Physicians 26
5.2.2 Exclusion Criteria for Patients 26
5.3 Study Enrollment Procedures 27
5.3.1 Physician Enrollment 27
5.3.2 Patient recruitment protocol 27
6. STUDY INTERVENTIONS 28
6.1 Interventions, Administration, and Duration 28

6.2 Concomitant Interventions 28
6.3 Adherence Assessment 28
7. STUDY PROCEDURES 30
7.1 Schedule of Patient Evaluations 30
7.2 Description of Evaluations 35
7.2.1 Measurements and study instruments 35
7.2.2 Enrollment, Screening, Baseline, and Randomization 36
7.2.3 Final Evaluation – Visit 5 (Week 12) 39
8. SAFETY ASSESSMENTS 41
8.1 Specification of Safety Parameters 41
8.2 Methods and Timing for Assessing, Recording, and Analyzing Safety Parameters 41
8.3 Adverse Events and Serious Adverse Events 41
8.4 Reporting Procedures 42
8.5 Follow-up for Adverse Events 42
8.6 Providing Support to Patients Experiencing Suicidality or Breaches in Continuity of Care 42
8.7 Notes on Ensuring Safety in the Context of COVID-19 when providing patients with tablets: 43
9. INTERVENTION DISCONTINUATION 43
10. STATISTICAL CONSIDERATIONS 44
10.1 General Design Issues 44
10.2 Sample Size and Randomization 44
10.3 Outcomes 45
10.3.1 Primary Outcome 45
10.3.2 Secondary Outcomes 45
10.4 Data Analyses 46
11. DATA COLLECTION AND QUALITY ASSURANCE 47
11.1 Data Collection Forms 47
11.2 Data Management 47
11.3 Quality Assurance 48
11.3.1 Training 48
11.3.2 Protocol Deviations 49
11.3.3 Monitoring 49
12. CIRCLE OF CARE FEATURE 49

DocuSign Envelope ID: 3DB15B31-6E89-4135-81C3-F2DBD9C216BB

13. PARTICIPANT RIGHTS AND CONFIDENTIALITY 50
  13.1 Informed Consent Forms 50
  13.2 Participant Confidentiality 50
  13.3 Ethical Considerations 51
14. COMMITTEES 51
15. PARTICIPANT COMPENSATION 52
16. PUBLICATION OF RESEARCH FINDINGS 52
17. REFERENCES 52

#### LIST OF ABBREVIATIONS

| | |
|---|---|
| ADs | Antidepressants |
| AI | Artificial Intelligence |
| AID-ME | Artificial Intelligence in Depression – Medication Enhancement |
| AUDIT | Alcohol Use Disorders Identification Test |
| AE | Adverse events |
| ACE | Adverse Childhood Experiences |
| BARS | Brief Adherence Rating Scale |
| CDA | Clinical decision aid |
| CRS | Clinician Rating Scale |
| DAST-10 | Drug Abuse Screening Test-10 |
| DSM-5 | Diagnostic and Statistical Manual of Mental Disorders, Fifth Edition |
| ER | Emergency Room |
| FIBSER | Frequency, Intensity, and Burden of Side Effects Rating |
| fMRI | Functional Magnetic Resonance Imaging |
| FoU | Frequency of Use |
| GAD-7 | Generalized Anxiety Disorder-7 |
| IRB | Institutional Review Board |
| LEC-5 | Life Events Checklist for DSM-5 |
| MADRS | Montgomery-Asberg Depression Rating Scale |
| MDD | Major depressive disorder |
| MDE | Major depressive episode |
| MINI | Mini International Neuropsychiatric Interview |
| PHQ-9 | Patient Health Questionnaire-9 |
| PRISE-20 | Patient Rated Inventory of Side Effects-20 |

| | |
|---|---|
| QIDS-SR 16 | Quick Inventory of Depressive Symptomatology Self-Report |
| RA | Research assistant |
| REB | Research Ethics Board |
| RCT | Randomized control trial |
| SAE | Serious adverse events |
| SAPAS | Standardized Assessment of Personality Abbreviated Scale |
| STAR-C | Scale to Assess Therapeutic Relationships - Clinician |
| STAR-P | Scale to Assess Therapeutic Relationships - Patient |
| WHODAS | World Health Organization Disability Assessment Schedule |

**DISCLOSURE STATEMENT**

Restricted Distribution of Documents

This document contains information that is confidential and proprietary to the study authors. This information is being provided to you solely for the purpose of evaluation and/or conducting a clinical trial for the study authors. You may disclose the contents of this document only to study personnel under supervision, ethics committees, or duly authorized representatives of regulatory agencies for this purpose under the condition that they maintain confidentiality. The contents of this document may not be used in any other clinical trial, disclosed to any other person or entity, and/or published without the prior written permission of the study authors. The foregoing shall not apply to disclosure required by any regulations; however, you will give prompt notice to the study authors of any such disclosure.

Any information that may be added to this document also is confidential and proprietary to the study authors and must be kept in confidence in the same manner as the contents of this document.

The Aifred Health Inc. protocol number for this study is IUSMD 18-04.

DocuSign Envelope ID: 3DB15B31-6E89-4135-81C3-F2DBD9C216BB

Protocol Number IUSMD 18-04 – Version 4.0

**AID-ME: Artificial Intelligence in Depression – Medication Enhancement:**

**A Randomized, Patient and Rater Blinded, Active-Controlled Trial of a Hybrid-Classic/Machine-Learning Enabled Clinical Decision Aid for Personalized and Individualized Pharmacological Depression Treatment Selection**

#### Sponsor Approval

DocuSigned by:
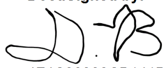
4F4C30388CB141B...

David Benrimoh, MD, CM, M.Sc.
Chief Science Officer

28/FEB/2022
Date: (DD/MMM/YYYY)

Protocol Number IUSMD 18-04 – Version 4.0

**AID-ME: Artificial Intelligence in Depression – Medication Enhancement:**

**A Randomized, Patient and Rater Blinded, Active-Controlled Trial of a Hybrid-Classic/Machine-Learning Enabled Clinical Decision Aid for Personalized and Individualized Pharmacological Depression Treatment Selection**

#### Investigator Signature Page

I have read the protocol and appendices and referenced documents. I understand the contents and intend to fully comply with all requirements. No changes will be made without formal authorization by Aifred Health Inc. in the form of a protocol amendment.

________________________
Print Name

________________________ ______________________
Signature of Investigator Date: (DD/MMM/YYYY)

### 1. SYNOPSIS

#### 1.1 Administrative Structure of the Study

Non sponsor parties, sponsor parties, study authors and service providers (as applicable) responsible for local management of the study are described in a separate document entitled "Administrative part of clinical study protocol".

**Global Principal Investigator:** Dr. Howard C. Margolese, MD.,CM., MSc, FRCPC
Address: Allan Memorial Institute, McGill University Health Centre (MUHC), 1025 Pine Ave W, Montreal, QC H3A 1A1, Canada


Co-Investigator: David Benrimoh, MD.,CM., MSc.
Address: 5910 David Lewis, Cote St. Luc, QC, CA, H3X 4A1

Key roles: Study design, data analysis, results reporting

Co-Investigator: Myriam Tanguay-Sela, PhD student
Address: 3640-1000, Sherbrooke St W, Montreal, QC, Canada, H3C 3P8

Key roles: Study design, data analysis, results reporting

Co-Investigator: Colleen Rollins, PhD Candidate
Address: Herchel Smith Building, University of Cambridge, Cambridge CB2 0SP, United Kingdom

Key roles: Results reporting, data analysis, quality assurance

Co-Investigator: Sonia Israel
Address: 600-1250 Guy St, Montreal, QC, Canada, H3H 2L3

Key roles: Results reporting, data analysis, write-up

Co-Investigator: Christina Popescu, MSc.
Address: 24-2325 Boulevard de Maisonneuve West, Montreal, QC, Canada, H3H 1L6

Key roles: Study coordination, RA management

Site directors listed in the list of investigators are also primary investigators at their sites.

#### 1.2 Participating Study Sites and Study Duration

Up to 75 physicians will participate in this study, for a total of up to 11 (2-10 physicians per site – planned average of 5) sites located in 2 countries (Canada and the USA). The list of investigators and participating physicians for each country is given in separate documents entitled "List of investigators".

**Study Duration :**

- Study duration for the participant: 12 weeks.
- Study initiation date: Quarter 1 (Q1) 2022
- Recruitment period: up to 6 months
- Study completion date: Quarter 3-4 (Q3-Q4) 2022

##### 1.3 Study Title

AID-ME: Artificial Intelligence in Depression – Medication Enhancement: A Randomized, Patient and Rater Blinded, Active-Controlled Trial of a **Hybrid-Classic/Machine-Learning Enabled Clinical Decision Aid for Personalized and Individualized Pharmacological Depression Treatment Selection**

##### 1.4 Objectives

The primary objective of this study is to conduct a naturalistic trial in order to establish the clinical utility, safety, and potential effectiveness of a machine learning based clinical decision aid for physicians treating patients with depression.

The secondary objective will be to glean insight into patterns of physician use of the clinical decision aid and to examine any secondary effects of use of the clinical decision aid on clinical practice, such as quality of record keeping, frequency of use of standardized rating scales, physician knowledge of guidelines and safety information, and utilization of non-pharmacological treatments, among others.

We will use the effect sizes found here to inform the power and sample size calculation for future studies including further randomized control trials aimed at testing specific aspects of the tool.

##### 1.5 Design and Outcomes

This will be a patient-and-rater blinded and partially physician blinded, physician randomized, prospective controlled trial to test the effectiveness and safety of the use of a clinical decision aid (CDA) by physicians treating adults with major depressive disorder (MDD). After baseline measurement of psychiatric symptomatology (see Section 7.2 Description of Evaluations), physicians who diagnose a patient with depression and who have been randomized to the clinical decision aid group will have the choice to use the CDA. Physicians not randomized to the clinical decision aid group will proceed according to usual guideline-informed practice with the benefit of patient symptom questionnaires provided to them by paper or secure, site-approved electronic means. Assessments of patient symptoms will be carried out as part of usual follow up visits, which will be scheduled within two weeks of the first visit, then at 4-6 weeks, 8 weeks and 12 weeks (total follow-up of 12 weeks). Primary effectiveness and safety outcomes will be change in patient symptoms, as well as adverse events and serious adverse events (including emergency room visits and hospitalizations). Secondary outcomes will be physician use patterns of the clinical decision aid, use of predictions made by the clinical decision aid, and effect of using the clinical decision aid on physician practice and on healthcare outcomes (see objectives list).

##### 1.6 Interventions and Duration

From the initial visit, at which point the patient will be selected into the study, the patients will be followed for 12 weeks, with follow-up appointments scheduled at 2 weeks, 4-6 weeks, 8 weeks and 12 weeks. Additional visits can be added as clinically appropriate and documented appropriately.

Physicians in both physician groups will determine, on an individual patient basis, whether to initiate treatment and what that treatment will be. The physicians in the CDA group will have access to the CDA and while they will be required to open the tool, they will be free to choose whether or not to use the information and results presented in it as part of their medical decision making. As such, no specific interventions are specified or required by the study other than opening the CDA.

The CDA will come packaged in the "study software". This software will have the following features (for both groups, except the predictive feature (the CDA itself) which will only be available to clinicians in the intervention group):

- A clinician-facing side, which will include all the clinician-administered questionnaires detailed in the appendix as well as a summary page for each patient detailing current and previous active treatments and care plans.
- Pages with symptom tracking information that allows clinicians and patients to see how symptoms have been tracked over time.
- The predictive feature (the CDA itself, only available to clinicians in the intervention group).
- A patient-facing side, where patients will be able to input their data and see their symptoms over time as well as their active treatments and care plan (but they will not have access to the CDA or its predictions).
- Patients will be identified in the software only by their participant code, not by name or medical record number.
- Integration with the electronic medical record at study sites is not currently planned.
- Secure messaging services allowing communication between patients and physicians. This is a feature of the existing software, not a part of this study, and the messages being sent will not be analyzed or viewed by study authors.
- All answers to self-reported or clinician-administered questionnaires as well as information about treatment and outcome entered into the study software is stored by that software on a secure Microsoft Azure server located in Canada (see below) and will be analyzed in the course of the statistical analysis of the study's outcomes.

DocuSign Envelope ID: 3DB15B31-6E89-4135-81C3-F2DBD9C216BB

The CDA itself is a predictive model which takes as input individual patient characteristics (called ‘features’) which are inputted by the clinician or by patient self-report, and outputs a list of all possible treatments, with each treatment associated with a predicted efficacy (likelihood to achieve response and likelihood to achieve remission, each expressed as a percentage). The treatments (which may include any approved treatment for depression) will be ordered by efficacy and presented to the clinician. In terms of predictions, in this study the application will only provide predictions for a set of first-line and combinations of first-line pharmacological treatments for depression, but the software will still include the ability to select other treatments (such as psychotherapy or neurostimulation) as the clinician sees fit.

Features inputted will include sociodemographic information (see demographics questionnaire), clinical information and medical history, and responses to validated symptom rating scales. The CDA is a predictive model based on deep-learning technology [1,2]. Deep learning is a form of artificial neural network (ANN) technology. ANN’s are networks of artificial computing units (called ‘neurons’ as they are inspired by elements of neuronal functioning) which are networked together in layers. The connections (called “weights”) between these units can be tuned to sculpt the network in response to feedback as the network tries to predict certain outcomes. As the network is trained on large sets of ‘labelled’ data (i.e. data with both baseline measurements and outcomes) it will sculpt itself, learning to use the set of input features to produce optimal predictions. It will then be ready to make predictions on individual data points it has not already trained on, thus becoming a predictive model that can use the features of an individual case (or patient) to make a prediction about that case. The network can be made deep - i.e. more layers can be added - to allow it to make use of more abstract or complex relationships between input features in order to make more accurate predictions; this is made more efficient by allowing the network to train in an unsupervised manner on the data set - allowing it to learn about associations between different input features without having to make predictions. Predictions made by the model can be interrogated by looking at which input features were most important in making a given prediction, as well as looking at how input features may naturally cluster with respect to most effective treatments (representing patient subgroups who respond preferentially to certain treatments). This information will be presented in a report to the clinician to accompany the prediction.

##### 1.7 Sample Size and Population

This study involves two populations: physicians and patients. All physicians who are either psychiatrists, family doctors or other primary care physicians (a role that is performed by internists at some U.S. sites) and who regularly treat depression in their practices, defined as at least one case of depression per month, will be eligible to enter the study. At the request of some sites, residents from the above specialties may also enter the study if they are being supervised by a clinician who is also part of the study. They will be randomized to either the “treatment as usual group” practice group (control group) or to the clinical decision aid group (intervention group). We plan to recruit between 30-50 physicians, up to a maximum of 75.

All patients 18 years or older of either sex who are treated by any of the physicians in the study for a new onset, recurrent, or chronic depression will be recruited into the study, for a target of 350 patients to meet our primary effectiveness endpoint. Up to a maximum of 500 patients could be recruited based on the maximum potential capacity of the sites identified thus far.

#### 2. BACKGROUND AND RATIONALE

##### 2.1 Background

Depression is estimated to affect more than 300 million people worldwide [3] and has a lifetime prevalence of 11.1% in Canada [4]. Symptoms of the disorder include a persistent feeling of sadness, emptiness, or hopelessness, anhedonia, reduced energy levels, sleep disturbances, appetite changes, cognitive impairments, psychomotor retardation, and recurrent thoughts of death or suicide [5]. In addition to the major toll that this depression takes on the lives of those affected and their families, the socioeconomic burden of the illness is enormous, costing $210.5 billion per year in the United States [6] and $32.3 billion per year in Canada [7].

While a range of effective treatments for depression exist, these are not equivalently effective for all patients and some patients can spend years finding the right choice from the dozens of medications, multiple psychotherapies, and five neurostimulation techniques currently available. Most patients and their physicians are forced to go through a “guess and check” approach to finding the right treatment [8]. For a patient with depression, trying a new treatment means several weeks of therapy or medication titration to determine if there is a positive effect. This is time lost in the patients’ lives - time that is potentially away from work and when they are not able to be fully present in their families’ lives. Inadequately treated depression also leads to risks of suicide and self-harm [3]. Moreover, many patients with depression will not improve after the first treatment. In the STAR*D trial, only about one third of patients improved after their first treatment trial, with decreasing response rates after further trials [9]. This means that the decision about which treatment to try is one that has significant consequences.

Aifred’s aim is to optimize treatment efficacy in psychiatry by leveraging a data-driven approach: making use of machine learning techniques along with a patient’s individual physiological profile to select the most effective interventions for that given patient’s depression. By training artificial neural networks on a vast quantity of naturalistic patient data, Aifred will learn to predict the best treatment or treatment cocktail at optimal dosages, while filtering out safety concerns such as drug-disease or drug-drug interactions. This in turn will lead to reduced treatment trials as compared to the “trial and error” approach, faster recovery or/and remission, while reducing the potential dangers that arise due to delayed treatment response, such as suicide. We hypothesize that this enhancement in treatment efficacy will increase patient treatment adherence, with the patient results feeding back into Aifred’s database to improve its predictive power. Considering the fact that depression is projected by the World Health Organization to be the leading cause of disease

DocuSign Envelope ID: 3DB15B31-6E89-4135-81C3-F2DBD9C216BB

burden by 2030 [10], helping those suffering from depression obtain remission is an utmost priority.

#### 2.2 Study Purpose and Rationale

##### 2.2.1 General Rationale

Major depressive disorder (MDD) is a serious mental illness that globally affects 11.1% of people over the course of their lives and is projected to be responsible for the majority of Disability Adjusted Life Years (DALYs) lost by 2030 [3, 10]. Mental health professionals -- psychiatrists, family doctors, therapists, nurses, counsellors -- and the patients they work with face another challenge: choosing the best treatment for a given patient. While a range of effective treatments do exist, these are not equivalently effective for all patients and some patients can spend years finding the right choice of treatment [9]. Currently, most patients and their physicians are forced to go through the current "trial-and-error" paradigm for treatment selection, where treatments are tried until something works [8]. For a patient with depression, trying a new treatment means several weeks of therapy or medication titration to start seeing if there is a positive effect, leading to time lost in the patient's life and increased risk for self-harm or suicide. Though existing guidelines do categorize the large array of treatment options into first-, second-, and third-line treatments [11]; and clinical experience has taught mental health professionals that certain types of medications or psychotherapy approaches work best in certain kinds of patients, there is not a systematic, evidence-based tool that can help clinicians choose treatments in a way that is personalized to a given patient.

A growing body of literature has identified potential predictors of treatment response across domains of neuroimaging, genetics, immunology, endocrinology, clinical profile and sociodemographics, among others [12-14]. Yet, to date, very few of them have entered into clinical practice, and we lack the clinical tools to systematically assess predictors to match patients with the treatments most likely to lead to response or remission [15]. Motivated by this critical gap between research and clinical practice, our research group developed a predictive model of individualized patient response to depression treatments using deep learning, a machine learning method based on layered artificial neural networks. The advent of this kind of application of machine learning to personalized medicine has already proven effective, for instance, in predicting which patients may benefit from preventive treatment for cardiovascular disease [16], in classifying depression severity based on resting-state fMRI data [17], and in predicting risk for suicide attempt based on demographic and clinical variables [18].

In light of the multitude of studies emphasizing the potential of biological, clinical, and sociodemographic markers to predict treatment response to antidepressants (ADs) in patients with MDD, it is probable that a clinical decision aid tool that integrates the most robust predictors of response to depression treatment with advanced machine learning techniques will provide specific, novel, and valuable information to guide clinical decision making, enable individualized treatment, reduce costs and side effects, and enhance clinical outcome.

##### 2.2.2 Study Specific Rationale

Now that we have trained a working model and tested it under artificial conditions in our initial ease of use study [19], we must validate it under clinical conditions. Initial safety and useability/feasibility was confirmed by a feasibility study. The next step is to further test safety and to begin to establish effectiveness in a realistic clinical setting of the study software as a whole. This is the purpose of the current study. Future studies would build on this study to further interrogate the value of the AI model in isolation and in longitudinal, recurrent treatment situations, but the current study should be able to demonstrate safety and effectiveness of the tool as a whole in the early phases of an initiation or change of treatment. While physicians randomized to the intervention group will be required to open the tool during every visit with their participating patients, the design of our study is intended to give physicians the freedom to use or ignore the CDA's information and features. Information about physician use patterns will be invaluable in the assessment of the technology and the design of future studies.

#### 3. STUDY OBJECTIVES

##### 3.1 Primary Objective

The primary objective is to assess the safety and potentially effectiveness, of a clinical decision aid for clinicians treating patients with major depressive disorder when compared to usual guideline-informed practice, and assessing that outcomes are not worse when using a clinical decision aid.

The primary endpoint of this study is to evaluate the remission rate of patients treated by physicians using usual guideline-informed practice (control group) as compared to patients treated by physicians using the CDA software.

**Objective 1.** To evaluate remission rates between groups. The hypothesis is that physicians using the CDA software will produce superior patient outcomes in terms of remission to those physicians using usual guideline-informed practice, as measured by rating scales (Montgomery-Asberg Depression Rating Scale (MADRS) at study exit using an intent-to-treat analysis). This is the primary endpoint for which the study is powered.

**Objective 2.** To evaluate safety and clinical outcomes compared to usual guideline-informed care. The hypothesis is that the clinical outcome (depressive symptomatology score as measured by the MADRS and the Quick Inventory of Depressive Symptomatology Self-Report (QIDS-SR16)) and rate of adverse and serious adverse events of patients whose physicians were randomized to the CDA group will not be worse than the outcome of patients whose physician used usual guideline-informed practice.

DocuSign Envelope ID: 3DB15B31-6E89-4135-81C3-F2DBD9C216BB

##### 3.2 Secondary Objectives

The secondary objective is to explore the usage of the CDA under natural conditions and impact on other healthcare outcomes (such as outpatient, ER, and inpatient visits). These should be considered exploratory in nature; the study is powered for the primary endpoint described in Objective 1.

**Objective 3.** To evaluate time to remission between groups. The hypothesis is that time to remission will be shorter in the CDA compared to the control group (both the MADRS and QIDS-SR16 remission definitions will be investigated).

**Objective 4.** To evaluate response rate between groups. The hypothesis is that the response rate (defined as 50% improvement in symptoms) and time to response, will be higher and lower, respectively, in the CDA compared to the control group (both the MADRS and QIDS-SR16 remission definitions will be investigated).

**Objective 5.** To evaluate between-group differences in patient functional outcomes. The hypothesis is that the physicians using the CDA software will produce superior patient outcomes to those physicians using usual guideline-informed practice, in terms of patient function as measured by a rating scale (World Health Organization Disability Assessment Schedule - WHODAS).

**Objective 6.** To evaluate physician engagement with the CDA. The hypothesis is that the physicians will use the CDA (beyond just opening it) in more than half of clinical interventions, as assessed by Frequency of Use (FoU) questionnaires to be completed after every visit.

**Objective 7.** To evaluate physician adherence to CDA suggestions. The hypothesis is that physicians will consider and adhere to the CDA suggestions at least 70% of the time as assessed by Frequency of Use (FoU) questionnaires to be completed after every visit.

**Objective 8.** To evaluate how physician behavior in the CDA group affects outcomes. The hypothesis is that improved patient outcomes will be present in the CDA group but may not depend entirely on high-frequency use of the model.
**Objective 9:** To evaluate patient engagement in providing data to the CDA. The hypothesis is that patients will answer their weekly questionnaires in the CDA greater than 67% of the time.

**Objective 10:** To evaluate between-group differences in hospital service utilization. The hypothesis is that visits to the emergency department and inpatient admissions and readmissions will be lower in the CDA group.

DocuSign Envelope ID: 3DB15B31-6E89-4135-81C3-F2DBD9C216BB

**Objective 11:** To evaluate between-group differences in medication adherence rates. The hypothesis is that medication adherence rates, as measured by the Brief Adherence Rating Scale (BARS) questionnaire, will be higher in the CDA group compared to the control group.

**Objective 12:** To evaluate between-group differences in outpatient appointment number. The hypothesis is that the number of completed outpatient appointments will be no different, or lower, in the CDA group compared to the control group.

Other exploratory data analyses will be conducted as applicable.

#### 4. STUDY DESIGN

This is a two-arm, patient-and-rater blinded, partially physician blinded, active-controlled, prospective, randomized trial to test the effectiveness and safety of the use of a CDA by physicians treating adults with major depressive disorder (MDD). Physicians will be randomized between CDA and control groups. Randomization will attempt to control for physician time of entry into the study and for physician type (i.e. primary care versus psychiatry) as this factor will have the greatest impact on the patient population recruited. Patients will be recruited from these physicians' practices and will not be randomized.

Patients will be blinded in the following way: they will be informed that they will have equivalent interactions with the software as patients in the other group but that physicians will use the data they provide in a different manner to manage their treatment. Patients in both groups will have equivalent interactions with the study software. As such, instead of informing patients in detail about the two arms of the study prior to them starting, we will inform patients that they will be "entering a study where new technology will be used by your clinician to help manage your depression." This is true in both patient groups because in the control group patients will be using the application to complete questionnaires, and physicians will get this information and can use it to tailor care. This method also allows physicians in both groups to engage in shared decision making effectively, while ensuring the patients have equivalent expectations about the "new technology" and ensuring that patients do not know if they are in the control or active arm. Note that all patients use the CDA to answer questionnaires and will have identical CDA experiences regardless of group assigned. As such, patients will be blinded to whether they are in the placebo or active group, and the experience they have interacting with the tool will be identical.

The study will include partial physician blinding in an effort to help reduce bias [20]. This will include blinding physicians to specific hypotheses and to the specific outcome measures (MADRS score at end, time to remission) and refrain from providing physicians with effect size estimates for the AI and control group interventions (to reduce expectation-related bias). As such, physicians will operate with a sense of equipoise regarding the intervention and will not have specific expectations generated by training for the study. In addition, as noted, the physicians in the control group would be an active control which has been previously proven to improve outcomes (following guidelines and measurement-based care) and the expectations of physicians in both groups should be that the intervention will help patients, over and above basic standard practice. This in turn helps accomplish part of the intent of physician blinding.

The primary outcome of the trial will be the change in patients' clinical symptoms scores (MADRS by blinded rater, QIDS-SR16) and quantification of adverse events and serious adverse events (including emergency visits and hospitalizations) in order to ascertain the safety, and effectiveness of the CDA, and to assess whether clinical outcomes and adverse event rates are not worse when using the CDA. Raters will be research staff identified by the Principal Investigator and/or could be provided by Aifred Health and trained in the use of the MADRS and MINI interview by phone, and will be blinded to patient participant group. The secondary outcome will be physician and patient use patterns of the clinical decision aid, including frequency of use of the CDA, use of CDA predictions, and physician- and patient-rating of the utility and ease-of-use of the CDA, in order to assess the CDA's naturalistic effectiveness and ease of use. Other secondary outcomes will include emergency room (ER) visits and hospitalizations/rehospitalization, amongst others (see full list of objectives). We will also record the length of time of each clinician-patient interaction in order to compare between both groups.

The trial will include two population groups: 1) physicians, including family doctors/primary care physicians and psychiatrists, and 2) patients of the physicians diagnosed with major depressive disorder. Physicians will be randomized to 1 of 2 arms: 1) the CDA group, who will have the choice to use the clinical decision aid in the treatment of their patients with MDD or 2) the active control group, who will receive guideline training at study start and the responses to patient's weekly questionnaires, but who will otherwise provide care according to their clinical judgment. Researchers will be unblinded to the trial arms as they will be required to provide the CDA physician group with both the CDA and use pattern questionnaires. However, raters administering the MADRS and MINI will be blinded to study arm.

The trial will be conducted at hospitals, local community services centres (i.e. CLSCs) or community health centres, private clinics, and family medicine clinics in Canada and in the USA. The total duration of the trial is 9 months, with an enrollment period of up to 6 months. Physicians will assess and treat individual patients for a minimum period of 3 months, with appointments scheduled at week 2, week 4-6, week 8 and week 12 at a minimum.

The intervention under investigation is access to the clinical decision aid by the CDA physician group. However, physicians will have full autonomy over whether or not they use the CDA (beyond opening the software) and whether or not they implement the CDA's predictions into patient treatment plans. The physician CDA group will be provided with the CDA upon initiation of the trial, during which they will also receive an orientation session and online guide with instructions on how to use the CDA. Continual user-support will be provided to physicians throughout the course of the trial should they have any questions or concerns regarding the CDA. Physicians will be completing a post-appointment questionnaire after each visit. Trained blinded raters will administer questionnaires to patients at the scheduled appointment time points in order to track patient symptoms. Self-report questionnaires will be filled out by the patient prior to each interaction with the clinician during the trial phase.

Given the context of COVID-19, where and when necessary visits will be conducted by site-approved telemedicine or by phone. When possible, telemedicine will be restricted to solutions where video chat and screen-sharing is possible in order to preserve the ability of clinicians in the CDA group to show the CDA to participants. Phone meetings may be utilized when video chat/screen sharing solutions fail or if the site cannot support video for part of the required visits. Study coordinators and/or research assistants may contact patients by phone in order to complete questionnaires, and the consent process may occur using electronic consent forms and phone conversations when in-person visits are impossible or pose a risk to the patient or research staff.

Further detail on electronic consent:

TrialStat Solutions eConsent platform will be used to document consent when informed consent is obtained from study participants remotely or in-person. eConsent will be controlled from within the eConsent module and the eConsent data will reside within the eConsent system. The patient ID will be generated after consent is obtained.

eConsent can be done with study staff in person, or eConsent forms can be completed at home at a participant's own pace. Should a participant choose to complete the forms at home, a member of the study staff will call them via telephone (after setting an appointment) to discuss the study, the consent form, and to answer any questions they may have. Patients will never be required to complete a consent form, on paper or electronically, right away and will have time to think about participating and to schedule subsequent visits or phone calls to ask more questions if needed.

Patient Consent

The site coordinator or designated person will generate an eConsent request. The eConsent module will either email the patient a link to submit eConsent in any browser or open a new window for the patient to complete the eConsent on site. The patient will then create an eConsent account with the required information. The patient will complete the consent module and confirm their agreement by entering their unique user ID and password. The consent will be encrypted and stored in a secure location. After the patient has completed the consent module, he/she will be registered and receive a unique ID number.

Patients will be able to access the signed consent form through their eConsent account. They can also withdraw consent or reconsent through their account.

Physician Consent

Physician eConsent will follow a similar path. The site coordinator or designated person will create the eConsent request and either a link to submit eConsent will be emailed, or a new window will open. The physician will then create an eConsent account with the required information. They will complete the consent module and confirm their agreement by entering their unique user ID and password. The consent will be encrypted and stored in a secure location.

Physicians will be able to access the signed consent form through their eConsent account. Physicians can also withdraw consent or reconsent through their account.

The consent forms used in the eConsent process and in the paper process will be identical.

#### 5. SELECTION AND ENROLLMENT OF PARTICIPANTS

This study will deal with two populations: physicians and patients. The physicians will be family doctors/primary care physicians and psychiatrists who treat depression as part of their regular practice. The patients will be any patients treated for depression by the physicians enrolled in the study.

##### 5.1 Inclusion Criteria

###### 5.1.1 Inclusion Criteria for Physicians

1. Any family doctor/primary care physician or psychiatrist accredited in Canada or the USA who treats patients with depression on at least a monthly basis, as well as residents from these specialties supervised by a participating physician.
2. Able to give informed consent to participate in the study.
3. Physicians must be comfortable prescribing the range of potential treatments which could have probabilities provided for them by the CDA

###### 5.1.2 Inclusion Criteria for Patients

1. All patients of the physicians in the study $\geq$ 18 years of age of either sex and diagnosed with major depressive disorder by a physician using the Diagnostic and Statistical Manual of Mental Disorders, Fifth Edition (DSM-5) criteria and by a blinded rater via the MINI diagnostic interview. The depression must also be of at least moderate severity on the MADRS as scored by a blinded rater.
2. All participants must be able to provide informed consent.
3. Contraception will be used as per established clinical guidelines and usual clinical practice for medications known to cause birth defects. The medications prescribed and the use of and type of contraception will be determined by the physicians in the study in consultation with their patients as would usually occur in clinical practice.
4. Patients must confirm that they are comfortable being treated for depression by their physician, who may propose a range of treatment options, such as medications or psychotherapies, consistent with best practice guidelines for depression which are included in the application. Physicians will be required, as in usual practice, to explain treatments to patients and patients will be able to give and withdraw consent for treatment in general or for specific treatments as in usual practice.

##### 5.2 Exclusion Criteria

###### 5.2.1 Exclusion Criteria for Physicians

There will be no exclusion criteria for physicians, provided they meet the inclusion criteria and are able to give informed consent.

###### 5.2.2 Exclusion Criteria for Patients

There will be the following exclusion criteria for patients:

1. Aged under age 18. We are excluding patients under age 18 because the data on which the model is trained is data from adults and we cannot be certain that it generalizes to patients under the age of 18.
2. Bipolar disorder of any type, as the data we have used to train the model does not allow for generalization to bipolar disorder (either pre-existing or as diagnosed according to DSM-5 criteria or during the MINI interview).
3. Inability or unwillingness of the individual to give informed consent.
4. Inability to manage patient in an outpatient setting (i.e. imminent suicidality).
5. An active major depression is not the main condition being treated (i.e. the patient has depressive symptoms in the context of severe substance abuse or a psychotic disorder, but a primary diagnosis of major depressive episode (MDE) cannot be made or would result in inappropriate care).
6. Inability to use the tool (i.e. patient cannot interface with a mobile phone or computer due to delirium, or another medical condition)*.

*Note that for patients who do not have access to mobile or desktop devices but are able to use them or to be trained to use them, these will be provided to them at no cost.

**Consent and intellectual disability:** Ability to provide informed consent will be determined by the physician proposing the treatment based on their ongoing communication with the patient. During their study training, clinicians will be given training on this process. Consent includes the following elements: is voluntary, requires adequate disclosure of information, demands proper material representation, should be appropriately communicated, must be specific to the treatment, can only be provided by mentally capable and (or) legally competent persons [21] and will be determined by the physician based on these criteria. The physician proposing treatment or other healthcare professional treating the patient will determine whether a potential participant has an intellectual disability. Intellectual disability will be determined based on DSM-5 criteria for an intellectual disability:

*A. Deficits in intellectual functions, such as reasoning, problem-solving, planning, abstract thinking, judgment, academic learning and learning from experience, and practical understanding confirmed by both clinical assessment and individualized, standardized intelligence testing.*

*B. Deficits in adaptive functioning that result in failure to meet developmental and sociocultural standards for personal independence and social responsibility. Without ongoing support, the adaptive deficits limit functioning in one or more activities of daily life, such as communication, social participation, and independent living, and across multiple environments, such as home, school work and school, work, and recreation.*

*C. Onset of intellectual and adaptive deficits during the developmental period.*

##### 5.3 Study Enrollment Procedures

###### 5.3.1 Physician Enrollment

Emails will be sent through the sites participating in the study, and study staff and local investigators will approach eligible clinicians at participating sites. If physicians express interest, they will be invited to an introductory session where the study and the artificial intelligence model will be described. Physicians opting to participate in the study will then have the opportunity to review the consent form, meet with a study representative for further information as needed, and then sign the consent form. As noted above, physicians will not be informed about specific study endpoints (aside from generally being informed that these are effectiveness and safety) and will not be informed about the expected effect sizes of either the CDA or the control group interventions. This could either be done at the information session, or at a later date, so that all physicians who desire will have adequate time to think about whether or not they wish to participate. Physicians may withdraw from the study at any time or speak to a study representative at any time for more information. As physicians will be randomized according to certain factors listed above, efforts will be made to recruit widely to ensure that meaningful randomization can take place.

Once physicians opting to participate in the study have provided their written informed consent, they will be randomized according to a block randomized and stratified randomization procedure [22]. Stratification will be done based on whether the clinician is a family doctor/primary care physician or psychiatrist, as this will have the greatest effect on the patient population being seen. Block randomization will be used to ensure appropriate randomization as physicians enter the study.

A log will be kept to track all cases where a physician was not eligible to participate and was excluded.

###### 5.3.2 Patient recruitment protocol

Eligible patients of the physicians included in the study will be offered induction into the study by the clinician. The patient presenting symptoms of depression or seeking consultation for possible depression will first meet with his/her physician. Upon diagnosis with depression, physicians will invite the patient to participate in the study, describing the study to the patient in lay terms (see "Physician to patient introduction to the trial"). If the patient is interested in participating, he/she would then be referred to a member of the research team to go through the consent process. If the patient is interested in participating, he/she will then be given the opportunity to speak with a member of the study team to learn more about the study and to address any questions they may have. For the patients of physicians randomized to the group receiving access to the model, patients will be asked if they consent to their physicians using the model to help them make a treatment decision and to share anonymized treatment outcomes and baseline treatment scores with the study authors. For the patients of physicians randomized to the treatment-as-usual group, they will be asked if they consent to their physicians sharing anonymized weekly and baseline questionnaire scores with the study authors and sponsor. All patients will sign an informed consent form. All physicians in the study will keep logs of the number of patients included in the study and the number of patients excluded, and the reasons for exclusion. These logs will be kept via a secure computer application or on paper and will be available in anonymized form to the study authors and sponsor. Once they provide consent, the patients will be asked to complete the screening questionnaires on a portable tablet computer or other device. Finally, the patient will return to meet with their physician to determine a treatment plan. No study procedures or data collection will occur prior to obtaining informed consent.

#### 6. STUDY INTERVENTIONS

##### 6.1 Interventions, Administration, and Duration

The intervention under study is physician access to a clinical decision aid software. Physicians will have full control over when they use the software (other than being required to log in), and whether or not they use the software's recommendations. They will be asked to keep a log of their interactions with the software. The log will be kept via a software provided by the study authors which will conform to standard data security protocols, or on paper where necessary.

The use of the software and the administration of questionnaires will take place during outpatient visits by study patients to study physicians. From the initial visit, at which point the patient will be selected into the study, the patients will be followed for twelve weeks, with follow-up appointments scheduled at 2 weeks, 4 to 6 weeks, 8 weeks, and 12 weeks at minimum. The study will last for a total of 9 months, with recruitment ending at 6 months so that all patients recruited by the end of the sixth month will have the necessary 12 weeks of follow-up. At the end of the trial, all clinicians will obtain access to the CDA if this has been shown to be safe as part of a separate continuation study; the CDA will continue collecting information on patient outcomes which will be analyzed up to one year after the end of the trial as part of this separate study.

##### 6.2 Concomitant Interventions

There will be no required or prohibited interventions. Physicians will have full control over whether or not to use the study software, if they have been randomized to the software group; full control over whether or not to follow the software's recommendations; and full control over which medication or other intervention to prescribe.

##### 6.3 Adherence Assessment

There will be two adherence measures in the intervention group: (1) of clinician use of the CDA to make predictions and (2) of adherence to CDA treatment suggestions. Adherence in the intervention group will be measured via monitoring clinician use of the software when seeing eligible patients enrolled in the study; this information will be automatically collected by the software.

DocuSign Envelope ID: 3DB15B31-6E89-4135-81C3-F2DBD9C216BB

The second adherence measure will be of adherence to software treatment suggestions. This will be assessed by the physician's study log and comparing to the treatments indicated as being chosen by the physician in the log with the corresponding recommendation recorded by the software. Importantly, physicians will never be told to use the recommendations or penalized for not doing so.

We will also measure adherence of patients to the treatments their physician prescribes as an outcome measure, via a standard questionnaire (BARS).

### 7. STUDY PROCEDURES

#### 7.1 Schedule of Patient Evaluations

| Assessment | Screening | Consent Baseline (0 to 14 days from Visit 1) | Treatment Visit 1 (Day 0) | Treatment Visit 2 (W2) ± 4 days | Treatment Visit 3 (W4-6) ± 4 days | Treatment Visit 4 (W 8) ± 4 days | Treatment Visit 5 (W12) ± 7 days |
|---|---|---|---|---|---|---|---|
| **Assessed by the clinician** | | | | | | | |
| Inclusion/ exclusion criteria | x | | | | | | |
| Enrollment | | | x | | | | |
| Intake assessment: Medical history, including previous psychiatric history and assessment of possible bipolarity | | | x | | | | |
| General physical examination | | | x | | | | x |
| Height | | | x | | | | |
| Weight | | | x | x | x | x | x |
| Blood pressure | | | x | x | x | x | x |
| Heart rate | | | x | x | x | x | x |
| Current medications | | | x | x | x | x | x |
| UKU Side Effect Rating Scale | | | | x | x | x | x |

| Assessment | Screening | Consent Baseline (0 to 14 days from Visit 1) | Treatment Visit 1 (Day 0) | Treatment Visit 2 (W2) ± 4 days | Treatment Visit 3 (W4-6) ± 4 days | Treatment Visit 4 (W 8) ± 4 days | Treatment Visit 5 (W12) ± 7 days |
|---|---|---|---|---|---|---|---|
| Post-Appointment Questionnaire | | | x | x | x | x | x |
| Scale to Assess Therapeutic Relationship (Clinician Version, STAR-C) | | | | | | | x |
| Clinician End Questionnaire | | | | | | | x |
| **Administered by the research assistant (RA)** | | | | | | | |
| Informed consent form | | x | | | | | |
| Brief Adherence Rating Scale (BARS) | | | x | x | x | x | x |
| Patient End Interview | | | | | | | x |
| Clinician End Interview | | | | | | | x |
| **Administered by Blinded Rater** | | | | | | | |
| MADRS (Montgomery-Asberg Depression Rating Scale) | | x | | | x | x | x |
| MINI (Mini International Neuropsychiatric Interview) | | x | | | | | |

| Assessment | Screening | Consent Baseline (0 to 14 days from Visit 1) | Treatment Visit 1 (Day 0) | Treatment Visit 2 (W2) ± 4 days | Treatment Visit 3 (W4-6) ± 4 days | Treatment Visit 4 (W 8) ± 4 days | Treatment Visit 5 (W12) ± 7 days |
|---|---|---|---|---|---|---|---|
| **Patient self-report questionnaires** | | | | | | | |
| Demographics questionnaire | | x | | | | | |
| Adverse Childhood Experiences (ACE) | | x | | | | | |
| Life Events Checklist for DSM-5 (LEC-5) | | x | | | | | |
| Patient Health Questionnaire (PHQ-9) | | | x (every week as of this time) | x (every week) | x (every week) | x (every week) | x (every week) |
| Generalized Anxiety Disorder Questionnaire (GAD-7) | | | x (every week as of this time) | x (every week) | x (every week) | x (every week) | x (every week) |
| Alcohol Use Disorders Identification Test (AUDIT) | | | x | | | | |
| Drug Abuse Screening Test (DAST-10) | | | x | | | | |
| Standardized Assessment of Personality (SAPAS) | | | x | | | | |
| World Health Organization | | | x | | | | x |

| Assessment | Screening | Consent Baseline (0 to 14 days from Visit 1) | Treatment Visit 1 (Day 0) | Treatment Visit 2 (W2) ± 4 days | Treatment Visit 3 (W4-6) ± 4 days | Treatment Visit 4 (W 8) ± 4 days | Treatment Visit 5 (W12) ± 7 days |
|---|---|---|---|---|---|---|---|
| Disability Assessment Schedule (WHODAS) | | | | | | | |
| Quick Inventory of Depressive Symptomatology (QIDS-SR16) | | x (in the 14 days prior to this appointment) | x | x (every 2 weeks) | x (every 2 weeks) | x (every 2 weeks) | x (every 2 weeks) |
| Custom Questionnaire | | x (patient part, in the 14 days prior to this appointment) | x (clinician part) | | | | |
| Patient Rated Inventory of Side Effects (PRISE-20) | | | x (one week after this appointment and then every two weeks) | x (every 2 weeks) | x (every 2 weeks) | x (every 2 weeks) | x (every 2 weeks) |
| Frequency, Intensity and Burden of Side Effects Rating (FIBSER) | | | x (one week after this appointment and then every two weeks) | x (every 2 weeks) | x (every 2 weeks) | x (every 2 weeks) | x (every 2 weeks) |
| Scale to Assess | | | | | | | x |

| Assessment | Screening | Consent Baseline (0 to 14 days from Visit 1) | Treatment Visit 1 (Day 0) | Treatment Visit 2 (W2) ± 4 days | Treatment Visit 3 (W4-6) ± 4 days | Treatment Visit 4 (W 8) ± 4 days | Treatment Visit 5 (W12) ± 7 days |
|---|---|---|---|---|---|---|---|
| Therapeutic Relationship (Patient Version, STAR-P) | | | | | | | |
| Patient End Questionnaire | | | | | | | x |

#### 7.2 Description of Evaluations

##### 7.2.1 Measurements and study instruments

In this section, we provide a brief description of each of the questionnaires used and other assessments performed, as defined in the "Assessment" column heading of the study procedures table described in Section 7.1 Schedule of Patient Evaluations**.**

*Assessed by the clinician*

**Medical history, including previous psychiatric history and assessment of possible bipolarity**: review of medical records and questionnaire regarding prior medical history. This includes previous psychiatric history and assessment of possible presence of bipolar affective disorder. This will also include a history of past and concomitant medications, as well as previous adverse medication reactions, which will be entered into the tool; this data entry can be undertaken by the clinical coordinator or the research assistant. This data will also be entered into the TrialStat electronic data capture platform.
**General physical examination (as clinically indicated)**: see attached form.
**Height (as clinically indicated)**: measurement of height for BMI calculation.
**Weight (as clinically indicated)**: measurement of weight for BMI calculation.
**Blood pressure (as clinically indicated)**: measurement of blood pressure.
**Heart rate (as clinically indicated)**: measurement of heart rate.
**Current medications**: determined from patient interview or medical records.
**UKU Side Effect Rating Scale**: a standard list of adverse events.
**Post-Appointment Questionnaire**: questionnaire assessing the physicians' experience with using the CDA or study software (administered after every appointment with a participating patient). This questionnaire will also assess whether or not the physicians in the treatment group considered the CDA's predictions in their choice of treatment and if they selected one of the three treatments with the highest predicted remission probability.
**Scale to Assess Therapeutic Relationship (Clinician Version, STAR-C)**: assessment of the therapeutic alliance from the clinician's perspective.
**Clinician End Questionnaire**: the clinician will be asked about their experience as a participant in the study.

Data about side effects and adverse events, as well as changes in medications after baseline, will be recorded by physicians in physician notes (as required by GCP guidelines on clinical trials) and will also be entered in the electronic data capture platform from TrialStat.

*Administered by the research assistant (RA - Aifred or Site-based)*

**Informed consent form**: see separate document.
**Brief Adherence Rating Scale (BARS)**: assessment of adherence to psychiatric medications.
**Patient End Interview**: the patient will be asked about their experience as a participant in the study.
**Clinician End Interview**: the clinician will be asked about their experience as a participant in the study.

*Administered by blinded rater*

**MADRS (Montgomery-Asberg Depression Rating Scale):** Rating scale for depression symptoms and severity.

DocuSign Envelope ID: 3DB15B31-6E89-4135-81C3-F2DBD9C216BB

**MINI (Mini International Neuropsychiatric Interview):** Diagnostic interview to confirm diagnoses and assess exclusion criteria.

***Patient self-report questionnaires***

**Demographics questionnaire**: we will record demographic information (such as age, gender, ethnicity, place of birth, languages spoken, marital status) via a questionnaire.
**Adverse Childhood Experiences (ACE)**: assessment of abuse, neglect and other adverse events in childhood.
**Life Events Checklist for DSM-5 (LEC-5)**: assessment of stressful life events.
**Patient Health Questionnaire (PHQ-9)**: screening, monitoring and measuring the presence and severity of depression over time [23].
**Generalized Anxiety Disorder Questionnaire (GAD-7)**: screening and measuring the severity of generalized anxiety disorder over time [24].
**Alcohol Use Disorders Identification Test (AUDIT)**: questionnaire developed by the World Health Organization (WHO) to assess alcohol consumption, drinking behaviors, and alcohol-related problems [25].
**Drug Abuse Screening Test (DAST-10)**: assessment of drug use (not including alcohol or tobacco use) in the past 12 months [26].
**Standardized Assessment of Personality (SAPAS)**: screening questionnaire for personality disorders.
**World Health Organization Disability Assessment Schedule (WHODAS)**: assessment of the effect of the patient's medical or mental health conditions on his or her life or ability to function.
**Quick Inventory of Depressive Symptomatology (QIDS-SR16)**: assessment of the severity of depression before, during, and after the study [27].
**Custom Questionnaire**: a set of demographics and symptom questions which are required for the machine learning algorithm. In addition to the self report questions completed by the patient no more than two weeks prior to Visit 1, there is also a Clinician Part of the Custom Questionnaire, comprising 7 questions from the HAM-D [36] and HAM-A [37] scales, which are completed by the treating physician using their application account at Visit 1 (in the Active Group), or completed on paper and then transmitted via the site coordinator to a research assistant to be entered into the application after Visit 1 (Active Control Group). Treating physicians will be provided interview guides for the HAM-D [38] and HAM-A [39] to assist them in asking these questions.
**Patient Rated Inventory of Side Effects (PRISE-20)**: assessment of the side effects of antidepressants.
**Frequency, Intensity and Burden of Side Effects Rating (FIBSER)**: assessment of the frequency, intensity and burden of the side effects experienced by the patient.
**Scale to Assess Therapeutic Relationship (Patient Version, STAR-P)**: assessment of the therapeutic alliance from the patient's perspective.
**Patient End Questionnaire**: the patient will be asked about their experience as a participant in the study.

##### 7.2.2 Enrollment, Screening, Baseline, and Randomization

***Clinician Recruitment***

DocuSign Envelope ID: 3DB15B31-6E89-4135-81C3-F2DBD9C216BB

Physicians will be recruited from approved sites and screened using a simple interview to ensure that they meet the inclusion criteria and do not meet the exclusion criteria. They will sign an informed consent form and have a chance to discuss with study personnel and ask questions about the CDA and other study technologies. Physician randomization to the intervention group or to the control group will proceed as described above, at least one month prior to the study start date. Physician recruitment will comply with 21 CFR 812.43 [28].

***Patient Recruitment***

Patients thought to be eligible by study clinicians will be asked by the clinician if they are interested in participating in the study and speaking to study personnel. If so, the patient can either speak to a research assistant (RA) over the phone or come early to their next appointment to meet with an RA, at which point they will be given more information about the study as well as a consent form. Prospective patients will be assured that participation is voluntary, does not affect their access to care, and that their consent to participate can be withdrawn at any time. Prospective patients will have two weeks upon receipt of the consent form to elect to participate, in which case they will sign the consent form.

Physicians in the control group will be asked to explain that patients are given access to a novel software which might help in the management of their treatment, and that they, as clinicians, have received training on best guideline-informed practice. Physicians in the active group will further explain the CDA without mentioning the control group.

***Baseline (0 to 14 days from Visit 1)***

After signature of the consent form and within 14 days prior to Visit 1 (Day 0), patients will have to complete the custom questionnaire. This questionnaire includes a subset of demographics and symptom questions as well as the QIDS-SR16, which are required for the machine learning algorithm at baseline.

***Visit 1 (Day 0)***

Patients should expect their appointment to last up to 1.5 hours longer than usual (this will include time with their physician, but also time to train on the study software or answer questionnaires). Prior to the appointment, the patients will meet with an RA virtually (up to 45 minutes). First, the RA or study coordinator will provide the patient with the demographics questionnaire, the ACE and the LEC-5. These self-report questionnaires should be completed as soon as possible after consent has been obtained. Then, the RA will ensure that the patient and doctor accounts are linked so that the patient can receive all the study questionnaires. The RA will ask the patient to complete the following questionnaires on the app as soon as possible: PHQ-9, GAD-7, AUDIT, DAST-10, SAPAS, and WHODAS. The RA will also inform the patient that every week, they will receive an email asking them to complete the PHQ-9 and GAD-7 questionnaires on the app. The RA will ask the patient to fill out the PRISE-20, FIBSER, and QIDS-SR16 every 2 weeks on their app. During the appointment, clinicians will complete the following assessments (up to 45 minutes):

- Medical history, including present history, previous psychiatric history and assessment of possible bipolarity
- Verify previous trials of treatment for depression
- Initial patient interview
- General physical examination (as clinically indicated)
- Height (as clinically indicated)
- Weight (as clinically indicated)
- Blood pressure (as clinically indicated)
- Heart rate (as clinically indicated)
- Current medications

In the control group, the patient and clinician will discuss treatment options and need for treatment and select a treatment; this treatment will be recorded in the study software. In the intervention group, the patient and clinician will discuss the need for treatment and then, if treatment is deemed to be needed, the clinician will use the CDA to generate a list of remission probabilities for different treatments. The patient and clinician will then decide whether to accept or reject these treatments and a treatment will be selected which will be recorded in the study software.

After the appointment, the study coordinator or RA will administer the BARS and the clinician will fill out the UKU.

After the appointment, the clinician will fill out the Post-Appointment Questionnaire. Clinicians in the intervention group will be asked to rate their experience with the CDA, while clinicians in the control group will be asked to rate the study software. This questionnaire will also assess whether or not the physicians in the treatment group considered the CDA's predictions in their choice of treatment and if they selected one of the three treatments with the highest predicted remission probability.

***Visit at Week 2, 4-6, and 8***

At the follow-up visits at Week 2 and Week 4-6, and Week 8 the patient will arrive early to complete the following patient-rated questionnaires, or will complete them at home (up to 20 mins per week): PHQ-9, QIDS-SR16, GAD-7, PRISE-20 and FIBSER. The clinician will complete: weight (as clinically indicated), blood pressure (as clinically indicated), heart rate (as clinically indicated), current medications and, after the appointment, the UKU and the Post-Appointment Questionnaire. Patients can expect these appointments to last up to 1 hour longer than usual.

In the intervention group, the clinician may elect to use the CDA to make a new prediction if the treatment previously selected is not effective or if the side effect burden is too high for the patient. In the control group, treatment may be changed if the treatment previously selected is not effective or if the side effect burden is too high for the patient. After the appointment, the study coordinator or RA will administer the BARS.

###### 7.2.3 Final Evaluation – Visit 5 (Week 12)

At the final 12-week visit (Visit 5), the patient will arrive early to complete the following patient-rated questionnaires, or will complete them at home: PHQ-9, GAD-7, PRISE-20 and FIBSER. The clinician will complete: weight (as clinically indicated), blood pressure (as clinically indicated), heart rate (as clinically indicated), current medications and, after the appointment, the UKU and the Post-Appointment Questionnaire. After the appointment, the RA will administer the BARS.

As soon as possible after this appointment, the RA will meet with patients who accept to participate in the End Interview, in which they will be asked about their experience as a participant in the study. The RA will ask the patient to fill out the QIDS-SR16, WHODAS, and STAR-P. The patient will also complete the self-report End Questionnaire containing questions about their experience in the study. At study end, the RA will meet with the clinician for the End Interview, in which they will be asked about their experience as a participant in the study. The RA will ask the clinician to fill out the STAR-C. The clinician will also have a self-report End Questionnaire containing questions about their experience in the study.

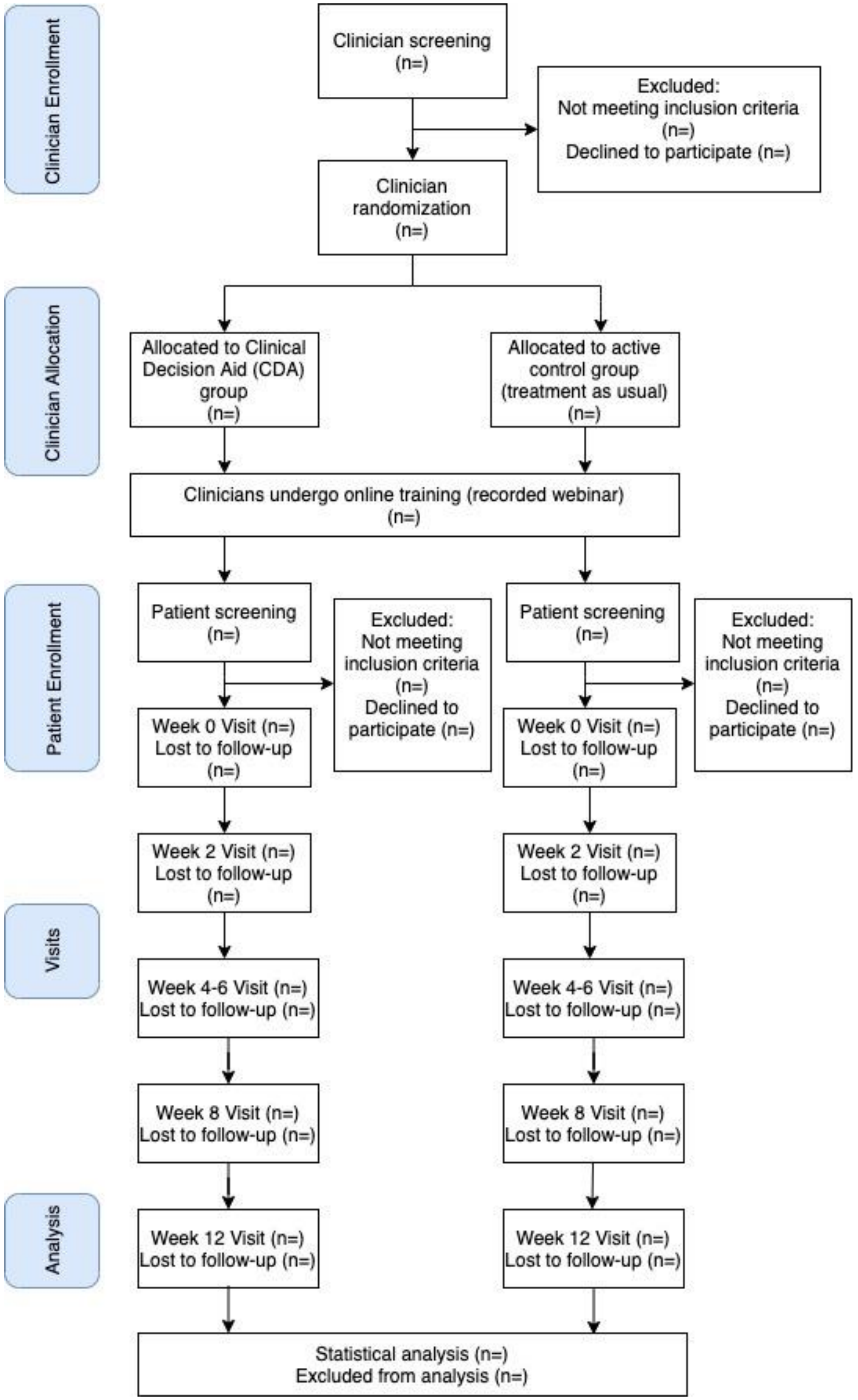

**Figure 1.** CONSORT diagram we will use to report study participant numbers.

#### 8. SAFETY ASSESSMENTS

##### 8.1 Specification of Safety Parameters

Safety parameters in this study will relate to the safety of recommendations made by the software. As all medications and other interventions will be decided upon by study physicians, adverse events related to treatment will not be a safety parameter per se, but will be tracked and compared between physician groups in the data analysis. Adverse events will be logged by clinicians in the post-appointment questionnaire and in their clinical notes with reference to the UKU questionnaire. These will then be entered into the case report form.

Specific safety parameters will be as follows:

1. Model suggests a medication that has a dangerous interaction with the patient's current medications in cases where the software was provided with this information.
2. Model suggests a combination of medications that could have dangerous interactions with each other.
3. The model makes a treatment suggestion which, in the opinion of the treating physician, diverges dangerously from usual practice.

##### 8.2 Methods and Timing for Assessing, Recording, and Analyzing Safety Parameters

Safety parameters will be recorded in two ways. Each time that a physician uses the software, they will be prompted by the software to report any violation of the above safety parameters. In addition, all treatment recommendations will be analyzed monthly by the study authors in order to identify any violations of the above parameters.

##### 8.3 Adverse Events and Serious Adverse Events

Adverse events (AE), defined as any unfavorable and unintended diagnosis, symptom, sign (including an abnormal laboratory finding), syndrome or disease which either occurs during the study, having been absent at baseline, or if present at baseline, appears to worsen, will be monitored and recorded in physician logs via entries into the software. Adverse events are to be recorded regardless of their relationship to the study intervention. Adverse events will be any worsening in symptomatology of depression or the appearance or worsening of other medical or physical symptoms or diagnoses which are not classified as serious adverse events. In addition, laboratory, imaging or EEG values which worsen from baseline will be counted as adverse events, though these measures will only be collected as the physicians involved see fit.

Serious adverse events (SAE), defined as any untoward medical occurrence that results in death, is life-threatening, requires inpatient hospitalization or prolongation of existing hospitalization, results in persistent or significant disability/incapacity, or is a congenital anomaly, will be monitored and recorded in physician logs via entries into the software.

Side effects to medication or other therapies, suicidal ideations and attempt, and worsening of symptoms of depression or other psychiatric comorbidities will be collected as solicited adverse events. Adverse events will be logged by clinicians in the post-appointment questionnaire and in their clinical notes with reference to the UKU questionnaire. These will be reported, as required, to relevant regulatory authorities.

The study committee will meet monthly over the course of the study to review these logs and will meet more frequently should there be an unexpectedly high number of AEs/SAEs.
In addition to patient AEs and SAEs, model safety parameters will be collected and reviewed as discussed above.

##### 8.4 Reporting Procedures

If the model makes any unsafe predictions, clinicians must alert the study committee immediately. The committee will then meet to review the incident and determine its cause and its likelihood of recurring. If the AE are significantly higher in the intervention group as opposed to the control group when these numbers are reviewed at monthly meetings of the study committee, then a stop to the study will be considered; a stop will happen automatically if there is a much higher rate of SAEs.

##### 8.5 Follow-up for Adverse Events

Physicians in the trial will have the responsibility to follow up on any AEs or SAEs as part of their usual clinical practice within 30 days of their occurrence, with an indefinite period of follow-up the duration of which will relate to the clinical situation. Of course, should transfers to other care or hospitalization be necessary, this is allowed within the study parameters and will follow the clinical need as determined by the physician in question.

##### 8.6 Providing Support to Patients Experiencing Suicidality or Breaches in Continuity of Care

During training, patients will be informed that if they are concerned for their safety they should present themselves to the ER or crisis services; all sites will also be required to furnish a list of local support or crisis services which will be provided to the patient at study start. If patients have suicidal thoughts, but are not concerned for their immediate safety, then they will be advised to contact their clinician or clinical service, who will be able to provide the appropriate care or provide the patient with their local list of services. However, if they cannot reach their clinical services, we will provide a phone number for the PI of their site or their delegate (who will be a clinician); they will be able to provide the patient with direction to appropriate care and with a pre-prepared list of local services and care options specific to their site. The medical monitor, Dr. Benrimoh, will also be able to assist the site PI should any issues arise that are specific to the application, and to log any of these events for reporting. In addition, should patients experience a sudden rupture in their care during the study (for example, their physician becomes unavailable,

DocuSign Envelope ID: 3DB15B31-6E89-4135-81C3-F2DBD9C216BB

or is hard to reach and is not seeing the patient at the frequency required by the study) the patient will be able to contact the site PI who will work to ensure that the patient has adequate follow-up. As part of the informed consent process, it will be explained to all participants that the clinical decision aid is not intended to be used for the identification, follow-up, prediction and/or monitoring of suicidal risk or for use in diagnosing or monitoring patient status for any particular disease or condition.

##### 8.7 Notes on Ensuring Safety in the Context of COVID-19 when providing patients with tablets:

Study coordinators or RAs will a) sanitize tablets between uses using sanitary wipes rated for antiviral activity by both Canadian and CDC guidelines (see links below); b) they will handle these using gloves rated for the use of the chosen disinfectant, masks (both of which will be changed between patients), and appropriate social distancing of at least 6 feet/2 meters and c) no direct contact between patients and study staff will be made; tablets will be put by study staff on a surface first cleaned with soap and water and then wiped-down with the antiviral cleaning product; study staff then will back up, and then patients will pick up the tablet. All cleaning activities will use products and procedures as per Canadian guidelines here: https://www.canada.ca/en/public-health/services/publications/diseases-conditions/cleaning-disinfecting-public-spaces.html and CDC guidance here: https://www.cdc.gov/coronavirus/2019-ncov/community/reopen-guidance.html . These guidelines will determine which products we use for disinfection, which gloves are used, and the social distancing and cleaning procedures.

#### 9. INTERVENTION DISCONTINUATION

Individual physicians will have the right to decide when to use or not use the model or its suggestions and as such have discretionary power to discontinue the intervention. However, the study protocol will be to discontinue use of the model for a given patient if it violates any of the safety parameters described above:

1. Model suggests a medication that has a dangerous interaction with the patient’s current medications in cases where the software was provided with this information.
2. Model suggests a combination of medications that could have dangerous interactions with each other.
3. The model makes a treatment suggestion which, in the opinion of the treating physician, diverges dangerously from usual practice.

Should the rate of these parameter violations or AE’s exceed what is to be expected in usual clinical practice in the opinion of the study committee, or should the intervention group have significantly more SAE’s than the control group, the study will be discontinued.

DocuSign Envelope ID: 3DB15B31-6E89-4135-81C3-F2DBD9C216BB

#### 10. STATISTICAL CONSIDERATIONS

##### 10.1 General Design Issues

This is a patient-and-rater blinded, partially physician blinded, randomized control trial. We are including a control group in order to be able to compare our intervention group to a control group with the same cultural background and resources with the ability to use current best-practice, which is guideline-driven measurement based care. It is cast primarily as a safety, non-inferiority and effectiveness trial to establish the safety and utility of the study software, and to justify further studies of more specific elements of the tool (for example, studies where patients are assigned to a treatment based on the AI alone).

The study will be analyzed as a cluster randomized trial, where clinicians are the cluster. Clinicians are randomized to avoid contamination effects that could result if patients were randomized and clinicians were to see some patients with and without the CDA.

Study objectives were noted above (see section 3. Study Objectives).

##### 10.2 Sample Size and Randomization

Given that this is the first clinical trial of a deep-learning powered clinical decision aid, estimating an effect size in order to calculate a sample size is difficult. In order to do so, we have employed two different techniques. Firstly, we examined a previous decision support trial [29], and based on this, expect detecting statistically significant differences in outcomes to require roughly 150 patients and 15-20 clinicians. With this as a foundation we then conducted a more rigorous sample size calculation for a specific and meaningful primary endpoint- binary remission as rated using the MADRS scale at study exit. We proceeded with the effect size calculation for a cluster randomized trial [32]. We set the intracluster correlation coefficient at 0.05, based on previous cluster randomized studies in depression (e.g. [33]). We set the P1, or baseline remission value, at 35%, based on the datasets we have analyzed to produce the model (and according with estimates from STAR*D and CO-MED, where similar remission rates were seen using measurement-based care). The cluster size was estimated at 7, as 7 patients per physician seemed to be a reasonable estimate for a study recruiting patients with a common condition over 6 months. Finally, the effect size was estimated at 20% difference in remission rate- 10% based on the AI treatment allocation (see [1], and as we have demonstrated in recent unpublished testing work) and 10% based on the fact that physicians in the active group will have access to a tool that provides measurement-based care and algorithm guided treatment (hence the hybrid nature of the tool reflected in the title); this 10% is based on previous studies of algorithm guided treatment (for example 34 and 35). In these previous studies, fairly large differences between algorithm guided treatment and treatment as usual were found- roughly 30-40% differences in remission rate. We estimated a reduced effect size of 10% because our active control includes guideline training and access to measurement-based care assessments, though it does not structure or force physician decisions based on this. At

90% power, these parameters generated a requirement for 47 physicians and 325 patients. We will therefore aim to recruit 350 patients and up to 50 clinicians in order to ensure an adequate sample size, with a maximum recruitment of 75 physicians and 500 patients (this is based on the maximum potential capacity of the sites identified thus far, though we are unlikely to recruit this many and so our target remains 50 clinicians and 350 patients). Note that this power calculation is only for the primary endpoint, and all secondary and exploratory endpoints are included as analyses of interest but without the expectation that the study is powered to find differences in all of them after correction for multiple comparisons. The aim of roughly 7-10 patients provided per clinician is also being chosen in order to ensure minimal disruption to the regular practices of participating clinicians. The results of this trial will allow for appropriate effect size calculation and the sample size needed for a future randomized control trial.

***Treatment Assignment Procedures***

Once all physicians opting to participate in the study have provided their written informed consent, they will be randomized according to a stratified randomization procedure [21]. Stratification will be done with the following covariates: number of years in practice, specialty (psychiatry or family medicine/primary care physician), and practice setting (community vs. tertiary care).

##### 10.3 Outcomes

We will test hypotheses of the CDA safety and effectiveness by assessing changes in primary and secondary clinical patient outcome measures, as well as outcome measures for the clinical decision aid, over patient treatment and use of the CDA.

###### 10.3.1 Primary Outcome

The primary outcome measure will be the severity of a patient's major depression and response to treatment, as measured by the remission rate using the MADRS at study exit using an intent-to-treat design. Remission status will be measured at study exit or at 3 months (study completion), whichever occurs first. A pre-post treatment design will be used with baseline measurements of the primary and secondary patient outcome variables taken before commencement of treatment (first visit with physician) and repeated at each visit (see schedule above for deviations). Patient outcome variables will also be measured between the pre- and post-treatment assessments (week 2, 4-6, 8 and 12 visits) to capture possible early therapeutic effects of treatment.

###### 10.3.2 Secondary Outcomes

**Patient Outcomes**

1. Remission as measured by the QIDS-SR16.
2. Treatment response (50% improvement from baseline on the MADRS and QIDS-SR16).
3. Time to remission and time to response.
4. General and social function as measured by the WHODAS.

5. Body weight and body mass index as a measure of weight gain or loss, a side-effect of certain ADs.
6. Medication and dosage used.
7. Number of completed outpatient appointments.
8. Possible side-effects and potential co-morbidities will be recorded.
9. Adverse events (side effects, hospitalization/rehospitalization, ER visit, suicide attempt).
10. Number of times needed to switch treatments.
11. The BARS will be used to assess adherence to treatment. Secondary patient outcome measures will be assessed at the same visits as primary patient outcomes.

**CDA Outcomes**

1. Frequency of use (of the CDA itself) by both patients and clinicians.
2. Frequency of use of CDA predictions by clinicians.
3. Likeability / user-interface rating (did the doctors or patients like using it, was it easy to use).

CDA outcomes will be assessed periodically (week 2, 4-6, 8, 12) to attain ongoing feedback of the tool's ease of use and utility.

##### 10.4 Data Analyses

**Quantitative:** Simple descriptive statistics will be used to characterize the physicians in the intervention and control groups; t-tests at an alpha of 0.05 will be used to determine if there are significant differences between these two groups with respect to the randomization criteria. Similarly, descriptive statistics will be used to characterize patients from both groups based on the severity of their depression, comorbidities, rates of adverse events, number of treatment switches, and demographic variables. Simple t-tests will be used to determine if the patient groups differed significantly at baseline or at the 2, 4-6 or 12-week timepoints on any of these measures.

The primary outcome in terms of remission rate will be assessed as a mean remission rate between-group difference tested at an alpha of 0.05. Outcomes in terms of adverse events and improvement in depressive symptoms and function would be evaluated at 2, 4-6, 8 and 12 weeks and compared between groups. In the intervention group, one important exploratory subgroup analysis would be the comparing outcomes between patients whose clinicians chose to use or not use the CDA's treatment recommendations (again measured as mean outcomes and tested via t-test at alpha = 0.05).

Physician use of the CDA, adherence to its recommendations, and experience in using it would be reported via descriptive statistics and synthesis of any narrative comments. Linear or logistic regression would be used to determine any factors that predicted physician use of the CDA or adherence to its recommendations.

Exploratory analyses will be conducted as applicable and could include for example, investigation of reduction in outpatient visits, reduction in in-patient stays, reduction in ER visits, and reduction in hospital readmission rates.

**Qualitative:** We will conduct verbal interviews in a subset of both populations (physicians and patients) to explore individual's needs and reactions towards the CDA. Via random selection, we will invite physicians (n=10) and patients (n=35) to partake in a short (20-30 min) semi-structured interview to explore their perspectives and needs on the use of clinical decisions aids in medical practice, especially in relation to the individual's experience as a participant in the study. Interviews will be transcribed, and the transcripts will be analyzed through a thematic step-by-step analyzing method [30, 31]. This is a form of inductive (data-driven) thematic analysis to identify, analyze, and report patterns (themes) within data. At least 2 researchers from the team will independently code a subset of the interviews to develop a thematic coding frameworks for categorizing themes. Each interview will be analyzed alongside the participant's questionnaire results from the qualitative sources reported above and will be considered in theme development. Coding discrepancies will be resolved through discussion leading to improvements in accuracy and homogeneity of the coding framework. Themes will be discussed and refined by the data analysis team. The remaining interviews and questionnaire results will be analyzed with the developed framework to confirm concordance of emerging themes. We will look at keywords and key themes in participant narratives and identify if any factors influence the frequency of these themes. We will also use the qualitative data to identify solutions for any issues encountered by participants during testing, and to identify ways in which Aifred can be improved.

#### 11. DATA COLLECTION AND QUALITY ASSURANCE

##### 11.1 Data Collection Forms

As noted above, this study will employ patient and clinician-rated measures. Patient measures will be completed digitally via a patient version of the study software. Clinician rated measures will be completed by the clinician enrolling patients in the study. As clinically indicated, height and weight for patients may be completed by qualified personnel. Physical examination may be completed by a resident as well as the enrolling clinician, if clinically indicated. The enrolling clinician may delegate the medical history to a resident.

##### 11.2 Data Management

Data management will be primarily done through study software developed by the investigators and sponsor which is compliant with relevant Canadian and U.S privacy regulations. All forms and data used in the study data analysis will be collected through this software, except for eConsent forms and data from physician notes which will be stored via the TrialStat Solutions eConsent and electronic data capture platform (see section 12.2 for more information). All the data will be stored on a secure server. Data, while not in use, will remain encrypted with a key available to only a select few individuals working regularly with the data. Any interaction with the data will be logged

DocuSign Envelope ID: 3DB15B31-6E89-4135-81C3-F2DBD9C216BB

and regular audits of who accessed the data will be made so as to ensure that only authorized personnel have worked with the data. Backups of the data will also remain encrypted in case of corrupted or altered files. Extended periods of time of inactive service will log the user out of the server so as to prevent unwanted access to the data. In case of an emergency, data can be quickly accessed by the administrator and if there is a break-in attempt (which would be logged in the log files), data will be migrated over to a different server and be destroyed from the original server.

Clinical sites will maintain adequate patient records which act as source documents of all study activities for each patient, including patient informed consent forms, which could be paper copies stored in locked cabinets at the study sites prior to being collected by the study team and then deposited in a locked cabinet at the Allan Memorial Institute, MUHC for sites located in Montreal, or at the location of the Principal Investigator for sites outside of the province of Quebec. Clinician consent forms will be in paper as well and stored in a locked cabinet at the Allan Memorial Institute, MUHC for sites in Montreal or at the location of the Principal Investigator for sites outside of the province of Quebec. Note that consent forms may also be completed electronically, using software that has been verified as being compliant for use in clinical research.

Data collection forms have already been detailed and will include measures of depressive and other psychiatric symptomatology and of adverse events and treatment adherence. These forms will all be in digital format, completed through the study software or the electronic data capture platform. Some forms (see above) will be completed by the study physician; others will be completed by the patient via a patient version of the study software or the electronic data capture platform. All forms will be identified using the patient code but not patient name.

All entries into the electronic data capture platform should be consistent with source documents. Data entry must be completed and made available as soon as possible after each patient’s visit in order that the monitor may verify the validity of the patient information. If discrepancies exist in the electronic data capture platform, the monitor will require the investigator or his delegate to make the appropriate corrections. Once all data management activities are complete, the database will be locked to ensure there is no manipulation of study data during the final analysis. After the data lock, the investigator will receive a copy of the patient data for archiving at the investigational site. The investigator must not destroy any records associated with the study without receiving prior written approval from the sponsor.

#### 11.3 Quality Assurance

##### 11.3.1 Training

Study staff will attend a day-long orientation and training session prior to the study start. They will be trained on study protocol, study evaluations, and the technical features of the study software and CDA and on the use of the electronic data capture platform and eConsent process. Clinicians recruited into the study will be required to complete a webinar that explains study protocols and assessments and will be contacted and met by study staff to review this information and ensure that the study software is properly installed on their computer, as applicable.

In more detail, study physician will need to complete webinars in which they will be taught to use the study's questionnaires by a qualified instructor; to use the study software (i.e. a tutorial on the interface and how to use it); and a refresher video on the CANMAT depression guidelines covering first, second, and third line treatments and the algorithm suggested by CANMAT.

In addition, study personnel will meet each study physician prior to study start in order to ensure the successful installation of the study software on the clinicians' computer, and to show the clinician the features of the software (i.e. how to log in, how to answer questions, how to navigate to the predictive model, how to understand the model's outputs, etc.).

Study personnel will be available throughout the study to provide technical support.

###### 11.3.2 Protocol Deviations

Protocol deviations include appropriate forms not being filled out, appointments missed, physician non-use of study software or the CDA. All this will be monitored via the study software, where the study committee will be able to see rates of missing or uncompleted assessments.

###### 11.3.3 Monitoring

Adverse events will be logged by clinicians in the post-appointment questionnaire and in their clinical notes with reference to the UKU questionnaire and will be accessible for review through the electronic data capture form. Monitoring of outcomes and adverse events will be done via the study software. The study authors will also be able to receive concerns of study clinicians or patients by email. Review of all outcomes and adverse events and clinician or patient concerns will occur at monthly study committee meetings.

#### 12. CIRCLE OF CARE FEATURE

The circle of care function allows clinicians, if they choose, to exchange messages with patients. This feature only allows clinicians to message a single patient at a time; they are not able to exchange information with other clinicians or other persons aside from the patient. These messages are text only messages and do not generate notifications. Clinicians must turn on this feature on a patient-by-patient basis (it is turned off by default) and are not required to use this feature. This is only an option in the Active group, where physicians and patients are connected on the application. In the Control group, where this option is not available as physicians are not connected to patient accounts, patients will be able to communicate with their physician as they normally would (i.e. by phone, email, or whatever other method the physician and patient jointly choose to use as part of normal care). This circle of care feature is not a key part of the intervention, is optional, will likely not be used by most clinicians based on experience in feasibility studies, and can be easily replaced by outside-of-application communication. This difference between groups is not expected to bias results, and is included as an option in the Active group in order to utilize all the features of the CDA as it is intended to be released in the future (with a view to ensuring that no issues or safety concerns arise as a result of the feature). With respect to safety, patients in the Active group are expressly trained not to rely on the application for urgent communication and are not to use the circle of care feature to communicate safety concerns. Patients in both groups will be provided with the same safety information (the list of local resources) and patients in both groups will be able to contact their clinical teams as they would during usual practice outside of the study.

#### 13. PARTICIPANT RIGHTS AND CONFIDENTIALITY

##### 13.1 Informed Consent Forms

A signed consent form will be obtained from each participant. The consent form will describe the purpose of the study, the procedures to be followed, and the risks and benefits of participation. A copy will be given to each participant or legal guardian and this fact will be documented in the participant's record. Forms will be available in French and in English.

##### 13.2 Participant Confidentiality

Study patients will have their confidentiality protected as a core aim of the study. No participant names will be directly associated with study data; all study data will be associated with a participant code. This code will be assigned to the participant at the time of enrollment, and their name will be linked to their code only on the paper informed consent form. These codes will be generated by the study authors and then distributed to study clinicians and staff, who will then assign them as patients are recruited. The study clinicians or staff will record the code chosen on the informed consent form, which will be stored in a locked cabinet at their practice until the end of the study, when these forms will be collected by study staff in order to be centrally stored in a locked cabinet at the Allan Memorial Institute, MUHC, unless the local site prefers the consent form be kept on site by the local PI in a similar secure locked cabinet, or a software approved for this purpose is used to collect consent forms. All information entered into the study software or the CDA will therefore be de-identified. Data collected by the study will be stored on a secure server for twenty-five (25) years after the study's end date. As is described in section 11.2, with respect to technical and software safeguards to protecting the data stored on the server, all patient data will remain encrypted when not in use and regular audits of those who access the data will be made. Any member of the research team will be required to go through an authorized individual to see the data and will be accompanied during their use. The informed consent forms of both clinicians and patients will be destroyed after twenty-five (25) years. Access to this data will be limited to the study authors and investigators designated as collaborators by the study authors.

Paper forms to be digitized will be digitized and kept on an isolated (non-internet connected) computer for analysis.

Any forms (other than the eConsent forms described above) completed outside the study software will be completed using a Microsoft Azure server located in Canada (Quebec City) and will be identified only by the patient and/or physician code and will include no identifying or personal information, only information regarding experience with the tool or safety information completed by physicians.

Data assessed by physicians and recorded in physician notes will be collected via an e-CRF and stored in a secured database in a dedicated hosting environment owned by TrialStat, a Canadian company. TrialStat's primary data center is in Somerset, New Jersey and is provided through Rackspace. TrialStat operates entirely on the Microsoft Stack and uses Microsoft Azure as a hot backup data center. The dedicated hosting environment is 21 CRF Part 11, HIPAA and GDPR compliant, SOC1, SOC II and PCI compliant. All data is encrypted and protected via a managed firewall and the TrialStat Network is monitored for intrusion detection.

##### 13.3 Ethical Considerations

The study will be conducted according to ethical principles stated in the Tri-Council Policy Statement 2nd Edition (TCPS2) and ethics approval will be obtained before initiating study. Good Clinical Practice guidelines, which inform the conduct of clinical trials, will also be followed. Informed consent will be obtained from all participants and consent forms will clearly detail study aims, participant rights, and measures to ensure confidentiality; it will also ensure that patients' rights to autonomy and dignity are respected.

#### 14. COMMITTEES

We will have a **study steering committee** that will meet monthly (or at a frequency defined by the committee) to monitor outcomes, providing support to study clinicians, data analysis and publication. The study committee will be comprised of the study investigators, invited researchers, site coordinators. The committee will also advise study authors on study management and trial design issues, as well as on exploratory analyses beyond those defined in the protocol. The committee will be otherwise comprised of psychiatrists or family doctors/primary care physicians, a statistician or epidemiologist, and one person with lived experience of mental health treatment, whose names, titles, and affiliations are the following:

| | | |
|---|---|---|
| Jordan Karp, MD (Committee chair) | Psychiatrist | University of Arizona |
| Sagar Parikh, MD, CM, FRCPC | Psychiatrist | University of Michigan |
| Soham Rej, MD, MSc, FRCPC | Psychiatrist | McGill University |
| Howard Magolese, MD, CM, MSc, FRCPC | Psychiatrist | McGill University |
| Shirin Golchi, PhD | Statistician | McGill University |
| Erica Moodie, PhD | Statistician | McGill University |
| Fanny Hersson-Edery, MD | Family doctor | McGill University |

| | | |
|---|---|---|
| Martin Enault | Person with lived experience | Centech Mtl |

A Data Safety Monitoring Board (DSMB) will receive reports from the study committee and ensure adherence to protocol and ethical requirements. This committee will meet three times: prior to study start, at three months, and at the end of the study. It will also meet as needed to review any complaints or concerns raised by participants, the study committee, the IRB, or others. This committee will be comprised of two study investigators (H. Margolese and D. Benrimoh), who will be non-voting members present in order to report to the committee. Serge Beaulieu (MD, PhD, FRCPC, McGill University), Warren Steiner (MD, FRCPC, FAPA, McGill University), and Mark Speechley (PhD, Western University) will be the committee's voting members. This committee may hold closed meeting without the non-voting members if they so choose will have the power to order a halt to the study or to trigger a review by the IRB should they feel this is necessary.

#### 15. PARTICIPANT COMPENSATION

In order to ensure that all physicians and patients can participate in the study without financial considerations being a limiting factor, participants will be fairly compensated for their time, as evaluated and approved by each IRB/REB. We recommend the following compensation:

- Patients will be compensated $20 per visit for the 5 key study visits, and 20$ for the optional end interview, up to $120 total
- Physicians will be compensated $200 for the training session, $100 for the optional end interview, and $100 for the initial assessment of each of the first seven patients, for a total of up to $1000 per physician.

In choosing these sums we tried to stay close to what we have used in IUSMD 19-08. There, physicians were paid $400 ($300 for a training session and $100 for an interview). The training sessions, now that we have more experience, should be more efficient hence the change in price. We have added the compensation for initial patient assessment as this requires time and attention from the physician and these costs are not covered by insurance plans. The patient compensation is equivalent to that in 19-08 ($20 per visit).

#### 16. PUBLICATION OF RESEARCH FINDINGS

We intend to publish results in a peer-reviewed journal in accordance with accepted scholarly standards.

DocuSign Envelope ID: 3DB15B31-6E89-4135-81C3-F2DBD9C216BB

Depression: A Deep Learning Analysis of STAR*D and CO-MED Data. BioRxiv. doi: 10.1101/679779

2. Mehltretter, J., Rollins, C., Benrimoh, D., Fratila, R., Perlman, K., Israel, S., … Turecki, G. (2020). Analysis of Features Selected by a Deep Learning Model for Differential Treatment Selection in Depression. Frontiers in Artificial Intelligence, 2. doi: 10.3389/frai.2019.00031
3. World Health Organization. (2017). Depression and other common mental disorders: global health estimates (No. WHO/MSD/MER/2017.2). World Health Organization.
4. Lam, R. W., McIntosh, D., Wang, J., Enns, M. W., Kolivakis, T., Michalak, E. E., ... & Milev, R. V. (2016). Canadian Network for Mood and Anxiety Treatments (CANMAT) 2016 clinical guidelines for the management of adults with major depressive disorder: section 1. Disease burden and principles of care. The Canadian Journal of Psychiatry, 61(9), 510-523.
5. American Psychiatric Association. (2013). Diagnostic and statistical manual of mental disorders (5th ed.). Washington (DC): The Association.
6. Greenberg, P. E., Fournier, A. A., Sisitsky, T., Pike, C. T., & Kessler, R. C. (2015). The economic burden of adults with major depressive disorder in the United States (2005 and 2010). The Journal of clinical psychiatry.
7. Sutherland, G. (2016). Healthy brains at work: estimating the impact of workplace mental health benefits and programs. Conference Board of Canada.
8. Leuchter, A. F., Cook, I. A., Hunter, A. M., & Korb, A. S. (2009). A new paradigm for the prediction of antidepressant treatment response. Dialogues in clinical neuroscience, 11(4), 435.
9. Warden, D., Rush, A. J., Trivedi, M. H., Fava, M., & Wisniewski, S. R. (2007). The STAR* D Project results: a comprehensive review of findings. Current psychiatry reports, 9(6), 449-459.
10. Lépine, J.-P., & Briley, M. (2011). The increasing burden of depression. Neuropsychiatric Disease and Treatment, 7(Suppl 1), 3–7. http://doi.org/10.2147/NDT.S19617
11. Parikh, S. V., Quilty, L. C., Ravitz, P., Rosenbluth, M., Pavlova, B., Grigoriadis, S., … the CANMAT Depression Work Group. (2016). Canadian Network for Mood and Anxiety Treatments (CANMAT) 2016 Clinical Guidelines for the Management of Adults with Major Depressive Disorder: Section 2. Psychological Treatments. Canadian Journal of Psychiatry. Revue Canadienne de Psychiatrie, 61(9), 524–539. http://doi.org/10.1177/0706743716659418
12. Kemp, A. H., Gordon, E., Rush, A. J., & Williams, L. M. (2008). Improving the prediction of treatment response in depression: integration of clinical, cognitive, psychophysiological, neuroimaging, and genetic measures. CNS spectrums, 13(12), 1066-1086.
13. Phillips, M. L., Chase, H. W., Sheline, Y. I., Etkin, A., Almeida, J. R., Deckersbach, T., & Trivedi, M. H. (2015). Identifying predictors, moderators, and mediators of antidepressant

DocuSign Envelope ID: 3DB15B31-6E89-4135-81C3-F2DBD9C216BB

response in major depressive disorder: neuroimaging approaches. American Journal of Psychiatry, 172(2), 124-138.
14. Strawbridge, R., Young, A. H., & Cleare, A. J. (2017). Biomarkers for depression: recent insights, current challenges and future prospects. Neuropsychiatric disease and treatment.
15. Simon, G. (2011). What little we know about tailoring depression treatment for individual patients. Depression and Anxiety, 28(6), 435-438.
16. Weng, S. F., Reps, J., Kai, J., Garibaldi, J. M., & Qureshi, N. (2017). Can machine-learning improve cardiovascular risk prediction using routine clinical data?. PloS one, 12(4), e0174944.
17. Ramasubbu, R., Brown, M. R., Cortese, F., Gaxiola, I., Goodyear, B., Greenshaw, A. J., ... & Greiner, R. (2016). Accuracy of automated classification of major depressive disorder as a function of symptom severity. NeuroImage: Clinical, 12, 320-331.
18. Passos, I. C., Mwangi, B., Cao, B., Hamilton, J. E., Wu, M. J., Zhang, X. Y., ... & Soares, J. C. (2016). Identifying a clinical signature of suicidality among patients with mood disorders: A pilot study using a machine learning approach. Journal of affective disorders, 193, 109-116.
19. Benrimoh, D., Tanguay-Sela, M., Perlman, K., Israel, S., Mehltretter, J., Armstrong, C., ... & Vahia, I. V. (2020). Using a Simulation Centre to Evaluate the Effect of an Artificial Intelligence-Powered Clinical Decision Support System for Depression Treatment on the Physician-Patient Interaction. medRxiv.
20. Page, S. J., & Persch, A. C. (2013). Recruitment, Retention, and Blinding in Clinical Trials. American Journal of Occupational Therapy, 67(2), 154-161. doi:10.5014/ajot.2013.006197
21. Neilson, G., & Chaimowitz, G. (2015). Informed Consent to Treatment in Psychiatry. Canadian Journal of Psychiatry. Revue Canadienne de Psychiatrie, 60(4), 1–11.
22. Suresh, K. (2011). An overview of randomization techniques: An unbiased assessment of outcome in clinical research. Journal of Human Reproductive Sciences, 4(1), 8–11. http://doi.org/10.4103/0974-1208.82352
23. Kroenke, K., Spitzer, R. L., & Williams, J. B. (2001). The PHQ- 9: validity of a brief depression severity measure. Journal of general internal medicine, 16(9), 606-613.
24. Spitzer, R. L., Kroenke, K., Williams, J. B., & Löwe, B. (2006). A brief measure for assessing generalized anxiety disorder: the GAD-7. Archives of internal medicine, 166(10), 1092-1097.
25. Saunders, J. B., Aasland, O. G., Babor, T. F., De la Fuente, J. R., & Grant, M. (1993). Development of the alcohol use disorders identification test (AUDIT): WHO collaborative project on early detection of persons with harmful alcohol consumption- II. Addiction, 88(6), 791-804.
26. Skinner, H. A. (1982). The drug abuse screening test. Addictive behaviors, 7(4), 363-371.
27. Rush, A. J., Trivedi, M. H., Ibrahim, H. M., Carmody, T. J., Arnow, B., Klein, D. N., ... & Thase, M. E. (2003). The 16-Item Quick Inventory of Depressive Symptomatology

DocuSign Envelope ID: 3DB15B31-6E89-4135-81C3-F2DBD9C216BB

(QIDS), clinician rating (QIDS-C), and self-report (QIDS-SR): a psychometric evaluation in patients with chronic major depression. Biological psychiatry, 54(5), 573-583.

28. US Food and Drug Administration. (2017). CFR-code of federal regulations title 21.
29. Elliott, L. S., Henderson, J. C., Neradilek, M. B., Moyer, N. A., Ashcraft, K. C., & Thirumaran, R. K. (2017). Clinical impact of pharmacogenetic profiling with a clinical decision support tool in polypharmacy home health patients: A prospective pilot randomized controlled trial. PLOS ONE, 12(2), e0170905. https://doi.org/10.1371/journal.pone.0170905
30. Braun, V., & Clarke, V. (2006). Using thematic analysis in psychology. Qualitative research in psychology, 3(2), 77-101.
31. Braun V, Clarke V. (2012). Thematic analysis. In Cooper, H. E., Camic, P. M., Long, D. L., Panter, A. T., Rindskopf, D. E., & Sher, K. J. APA handbook of research methods in psychology, Vol 2: Research designs: Quantitative, qualitative, neuropsychological, and biological.
32. Rutterford, C., Copas, A., & Eldridge, S. (2015). Methods for sample size determination in cluster randomized trials. *International Journal of Epidemiology*, *44*(3), 1051–1067. https://doi.org/10.1093/ije/dyv113
33. Underwood, M., Lamb, S. E., Eldridge, S., Sheehan, B., Slowther, A.-M., Spencer, A., Thorogood, M., Atherton, N., Bremner, S. A., Devine, A., Diaz-Ordaz, K., Ellard, D. R., Potter, R., Spanjers, K., & Taylor, S. J. C. (2013). Exercise for depression in elderly residents of care homes: a cluster-randomised controlled trial. *The Lancet*, *382*(9886), 41–49. https://doi.org/10.1016/s0140-6736(13)60649-2
34. Guo, T., Xiang, Y.-T., Xiao, L., Hu, C.-Q., Chiu, H. F. K., Ungvari, G. S., Correll, C. U., Lai, K. Y. C., Feng, L., Geng, Y., Feng, Y., & Wang, G. (2015). Measurement-Based Care Versus Standard Care for Major Depression: A Randomized Controlled Trial With Blind Raters. *American Journal of Psychiatry*, *172*(10), 1004–1013. https://doi.org/10.1176/appi.ajp.2015.14050652
35. Adli, M., Wiethoff, K., Baghai, T. C., Fisher, R., Seemüller, F., Laakmann, G., Brieger, P., Cordes, J., Malevani, J., Laux, G., Hauth, I., Möller, H.-J., Kronmüller, K.-T., Smolka, M. N., Schlattmann, P., Berger, M., Ricken, R., Stamm, T. J., Heinz, A., & Bauer, M. (2017). How Effective Is Algorithm-Guided Treatment for Depressed Inpatients? Results from the Randomized Controlled Multicenter German Algorithm Project 3 Trial. International Journal of Neuropsychopharmacology, 20(9), 721–730. https://doi.org/10.1093/ijnp/pyx043
36. Hamilton M. A rating scale for depression. J Neurol Neurosurg Psychiatry 1960;23:56–62
37. Hamilton M . The assessment of anxiety states by rating. Br J Med Psychol 1959;32:50–55.
38. Williams J. SIGH-D 1988, inline: (Williams, 1988)
39. G.S. Bruss, Ph.D., A.M. Gruenberg, M.D., R.D. Goldstein, Ph.D., and J.P. Barber, Ph.D. Dave Garroway Laboratory for the Study of Depression, Institute of Pennsylvania Hospital 111 N. 49th St., Philadelphia, PA 19139. 1994

DocuSign Envelope ID: 3DB15B31-6E89-4135-81C3-F2DBD9C216BB
