## Supplementary material for "Artificial Intelligence in Depression – Medication Enhancement (AID-ME): A Cluster Randomized Trial of a Deep Learning Enabled Clinical Decision Support System for Personalized Depression Treatment Selection and Management": Final SAP

DocuSign Envelope ID: 29EC5C33-90E5-4076-9DE0-2CC10407A41A

### Statistical Analysis Plan

| | |
|---|---|
| **Protocol #:** | IUSMD 18-04 |
| **Protocol Title:** | AID-ME: Artificial Intelligence in Depression – Medication Enhancement: A Randomized, Patient and Rater Blinded, Active-Controlled Trial of a Hybrid-Classic/Machine-Learning Enabled Clinical Decision Aid for Personalized and Individualized Pharmacological Depression Treatment Selection |
| **Project Code:** | AH01DEK |
| **Trial Design:** | Physician-Randomized, Patient-and-Rater Blinded, Partially Physician-Blinded, Active-Controlled Intervention Trial |
| **Study Treatment:** | Clinical decision aid (CDA) software intervention and standard of care |
| **Participants:** | 30-50 physicians (maximum 75); 350 patients (maximum 500) |
| **Patient Treatment Period:** | 12 weeks |
| **Sponsor Contact and final analysis contact:** | David Benrimoh<br>Aifred Health Inc.<br>304-1600 rue Notre-Dame O, Montreal, QC, H3J 1M1, Canada<br>514-463-7813<br> |
| **Analysis Contact (old):** | Blue Neustifter<br>McDougall Scientific<br>789 Don Mills Rd, Suite 802<br>Toronto, Ontario, M3C 1T5<br>416-673-9478<br> |
| **Date:** | February 6, 2024<br>New date for v 1.1: Jan 30, 2024<br>Version 2.0 Feb 6 2024 |
| **Status:** | Version 2.0 |

#### Document Change Control

This section tracks the versions of this Statistical Analysis Plan (SAP). The last entry is the final agreed to version.

Approval for the final version is given by the contacts listed in this SAP. We note that the SAP was finalized after database locking due to the early end of the study and the fact that database lock and initial analyses on the complete dataset based on DSMB data pulls proceeded concurrently due to the imminent end of funding. All analyses are repeated as pre-specified on the final locked data. In addition, the SAP was adapted after unblinded data review. This was done because there was a lack of the remitters in the control group, which necessitated a change in statistical approach. The loss of access to the blinded statistician for analysis meant that only unblinded researchers were left to review the data directly.

| Version | Date of Issue | Author | Brief Description of Change |
|---|---|---|---|
| 1.0 | Aug 23 2022 | Blue Neustifter | Original version |
| 1.1 | Jan 30 2024 | David Benrimoh | Updated version- comments incorporated from original and deviations from protocol and original noted throughout |
| 2.0 | Feb 6 2024 | David Benrimoh | Updated after meeting with Shirin Golchi and Erica Moodie, statisticians. Clarifications made regarding statistical analyses, specifically adding certain sensitivity and secondary analyses and clarifying primary endpoint analyses given the lack of remitters in the control group. |

Note: due to loss of funding and early end of the study, the SAP was not completed by MacDougall but rather by Aifred Health, specifically medical monitor David Benrimoh; changes were limited to pre-noted comments (available in commented draft), clarification that “study exit

DocuSign Envelope ID: 29EC5C33-90E5-4076-9DE0-2CC10407A41A

visit” means any post-baseline visit with a MADRS for the MADRS analysis, and specifying that Fisher’s exact test will be used for MADRS group comparison as no remitters were available in the control group and also specifying that we will not correct for clinician type as all clinicians were specialty care. Any further deviations will be noted.

**Impact of Changes**

1. Version 1.0 - Original version.
2. Version 1.1 – Working version used for analyses. Comments on original integrated and deviations specifically related to the lack of MADRS remitters in the control group and the lack of primary care clinicians integrated.
3. Version 2.0 - Updated after meeting with Shirin Golchi and Erica Moodie, statisticians. Clarifications made regarding statistical analyses, specifically adding certain sensitivity and secondary analyses and clarifying primary endpoint analyses given the lack of remitters in the control group.

#### Signature Approval Page

Date of Final Protocol: 19-January-2022, Version 4.0

Amendments: N/A

Date of Final Plan: 06-FEB-2024

**I have reviewed the Statistical Analysis Plan. My signature below confirms my agreement with the contents and intent of this document.**

**Reviewed by:**

DocuSigned by:
4F4C30388CB141B...

2024/02/12 | 08:25:55 EST

Dr. David Benrimoh
Chief Science Officer
Aifred Health Inc.

Date:

DocuSigned by:
Maud Richard
6358B5239CC140C...

2024/02/12 | 08:31:57 EST

Maud Richard
Regulatory Officer
QA&RA Director, Clinical Trial Manager
Aifred Health Inc.

Date:

DocuSigned by:
Dr. Howard Margolese
E211BF01A4DE463...

2024/02/12 | 06:23:47 PST

Dr. Howard C. Margolese, MD.,CM., MSc, FRCPC
Global Principal Investigator for IUSMD 18-04 study protocol
Allan Memorial Institute, McGill University Health Centre (MUHC)

Date:

We would like to sincerely thank Dr. Shirin Golchi and Dr. Erica Moodie for their advice and guidance while preparing versions 1.1 and 2.0 of this SAP.

DocuSign Envelope ID: 29EC5C33-90E5-4076-9DE0-2CC10407A41A

### 1. List of Abbreviations Definition of Terms

| Abbreviation or Term | Definition |
|---|---|
| ACE | Adverse Childhood Experiences |
| AE | Adverse Event |
| APR | Analysis Programming Requirements - detailed programming specifications required to convert the CRF data into analysis/presentation data sets |
| BARS | Brief Adherence Rating Scale |
| BMI | Body Mass Index |
| CDA | Clinical Decision Aid |
| CI | Confidence Interval |
| CRF | Case Report Form |
| DMP | Data Management Plan |
| EDC | Electronic Data Capture (i.e. eCRF) |
| ePRO | Electronic Patient-Reported Outcome |
| ER | Emergency Room |
| GAD-7 | Generalized Anxiety Disorder Questionnaire |
| ICC | Intra-Cluster Correlation |
| MADRS | Montgomery-Asberg Depression Rating Scale |
| MDD | Major Depressive Disorder |
| MedDRA | Medical Dictionary for Regulatory Activities (coding for AEs) |
| MINI | Mini International Neuropsychiatric Interview |
| PHQ-9 | Patient Health Questionnaire |
| QIDS-SR16 | Quick Inventory of Depressive Symptomatology Self-Report |
| SAE | Serious Adverse Event |
| SAP | Statistical Analysis Plan |
| SDLC | Systems Development Lifecycle |
| SOC | System Organ Class (from MedDRA coding dictionary) |
| SOP | Standard Operating Procedure |
| STAR-C | Scale To Assess Therapeutic Relationship – Clinician |
| STAR-P | Scale To Assess Therapeutic Relationship – Patient |
| TEAE | Treatment Emergent Adverse Event |
| UKU-SERS | UKU Side Effect Rating Scale |
| WHODAS | World Health Organization Disability Assessment Schedule |
| WHODD | World Health Organization Drug Coding Dictionary managed by the Uppsala Monitoring Centre |

#### 2. Background

IUSMD 18-04 is a patient-and-rater blinded and partially physician-blinded, physician-randomized, prospective controlled trial to test the effectiveness and safety of the use of a clinical decision aid (CDA) by physicians treating adults with major depressive disorder (MDD). Physicians will be randomized to have access to the CDA tool in addition to the usual guideline-informed practice or to simply have access to the usual guideline-informed practice in treatment of patients diagnosed with MDD. Both groups of physicians will have access to results from assessments of patient symptoms (e.g. study questionnaires) to assist in guiding treatment.

The CDA is a predictive model which takes input individual patient characteristics which are inputted by the clinician or by patient self-report and outputs a list of all possible treatments, with each treatment associated with a predicted efficacy (likelihood to achieve remission, expressed as a percentage). The treatments (which include 8 first line and 2 combinations of first line medications individually approved for the treatment of depression) will be ordered by efficacy and presented to the clinician. In this study, the application will only provide predictions for a set of first line and combinations of first-line pharmacological treatments for depression, but the software will still include the ability to select other treatment (such as psychotherapy or neurostimulation) as the clinician sees fit.

Patients will not be randomized; rather, they will receive treatment under their physician as usual, with the physician's assignment (CDA group vs. active control group) determining the tools the physician has access to for treatment. Physicians in the active arm will not be required to accept treatments suggested by the CDA and will have opportunities to record feedback about CDA treatment suggestions if they are felt to be ineffective or unsafe. Physicians in the active arm will, however, be required to open the tool once per session, whether or not they use the information and results provided by it. Patients will be followed for 12 weeks from their initial visit in order to monitor their safety and any response/remission of depression. Patients in both groups answer questionnaires using the CDA and as they may have their accounts created in advance of their first study appointment (though after being consented and enrolled) the patient CDA accounts may collect somewhat more than 12 weeks of data.

#### 3. Objectives

##### 3.1. Primary Objectives

To evaluate remission rates between patients in the two groups: those under a physician with access to a CDA in addition to usual guideline-informed practice and those under a physician with only usual guideline-informed practice.

##### 3.2. Secondary Objectives

1) To evaluate time to remission between groups.

2) To evaluate response rate between groups.
3) To evaluate time to response between groups.
4) To evaluate between-group differences in patient functional outcomes.
5) To evaluate physician engagement with the CDA.
6) To evaluate physician adherence to CDA suggestions.
7) To evaluate how physician behaviour in the CDA group affects outcomes.
8) To evaluate patient engagement in providing data to the CDA.
9) To evaluate between-group differences in hospital service utilization.
10) To evaluate between-group differences in medication adherence rates.
11) To evaluate between-group differences in outpatient appointment numbers.

##### 3.3. Safety Objectives

To evaluate safety and clinical outcomes between the two groups.

#### 4. Study Design

Physicians at participating sites will be screened and enrolled into the study, then randomized to either have access to study software that contains the CDA or to receive only guideline training and records and summaries from patient outcomes surveys.

After a baseline measurement of psychiatric symptomatology, physicians who diagnose a patient with depression and who have been randomized to the CDA group will have the choice to use the CDA. Physicians not randomized to the CDA group will proceed according to usual guideline-informed practice with the benefit of patient symptom questionnaires provided to them by paper or secure, site-approved electronic means. Physicians with access to the CDA will be required to open the CDA at each patient visit but will not be required to follow CDA suggestions or predictions. Subjects will have outpatient visits scheduled within 2 weeks of the first visit, at 4-6 weeks, 8 weeks, and 12 weeks.

##### 4.1. Primary Outcomes

Proportion of remission (overall score of less than 11 using the Montgomery-Asberg Depression Rating Scale (MADRS) at study exit or at the Week 12 visit, whichever occurs first.

##### 4.2. Secondary Outcome(s)

- MADRS remission proportion at Week 4-6, and Week 8 visits (Note the previous SAP version noted MADRS at Week 2, which was not collected).

- QIDS-SR16 remission proportion (QIDS-SR16 overall score of 0-5) at Week 2, Week 4-6, Week 8, and Week 12 visits
- Treatment response at each scheduled visit as measured by a 50% decrease from baseline in MADRS.
- Treatment response at each scheduled visit as measured by a 50% decrease from baseline in QIDS-SR16
- Time to MADRS remission
- Time to QIDS-SR16 remission
- Time to MADRS response
- Time to QIDS-SR16 response
- Change in general and social function from Day 0 to end of study/Week 12 as measured by the World Health Organization Disability Assessment Schedule (WHODAS)
- Percent change in body weight at each scheduled visit
- Percent change in body mass index (BMI) at each scheduled visit
- Summarization of medication and dosage used as physician-prescribed treatment.
- Number of completed outpatient appointments.
- Number of treatment switches during study
- Adherence to treatment, as captured by the Brief Adherence Rating Scale (BARS)
- Clinician quantitative ratings of CDA usefulness and ease of use at each visit and at study end
- Proportion of active group clinicians adhering to one of the top three medications predicted by the CDA.
- Proportion of remission and response in the subgroup of patients whose clinicians prescribed CDA adherent treatment (for the top 3 treatments) who did not require medication switch and who achieved response and remission (considered separately) compared to patients who did not get CDA-optimal treatment.
- Time to response and remission of patients whose clinicians prescribed CDA adherent treatment (top 3 treatments) who did not require medication switch and who achieved response and remission (considered separately) compared to patients who did not get CDA-optimal treatment (top 3 treatments)

- Proportion of remission and response of all patients (including control group) who got CDA-optimal treatment (top 3) compared to those who did not.
- Proportion of patients who got vs. did not get CDA optimal treatment who required a medication switch, and proportion who required an adjuvant/augmentation medication, compared to those who did not get CDA-optimal treatment.

##### 4.3. Safety Outcomes

- Incidence of Adverse Events (AEs)
- Incidence of Serious Adverse Events (SAEs)
- Incidence of hospitalization/rehospitalization
- Incidence of Emergency Room (ER) visits
- Incidence of suicide attempts

#### 5. Data Management

##### 5.1. Data Management

Data will be collected at the sites via an electronic data capture (EDC) system, via paper forms that are later entered into the EDC, or via an electronic patient-reported outcome (ePRO) application. The development and management of the EDC database will be conducted by McDougall Scientific. The study-specific applications will be developed based on the protocol requirements and following the full Systems Development Lifecycle (SDLC). All data activities are performed on validated computer systems as per 21CRF11 and kept under the control of the McDougall Standard Operating Procedures (SOPs). All programming will be performed using SAS version 9.4, which resides on a Windows Server 2012R2 at McDougall Scientific in Toronto, Canada.

The application design will, where appropriate, provide choice fields in the form of checkboxes, buttons and lists to aid in ensuring high quality standardized data collection. In addition, Data Logic Checks (or data Edit Checks) will be built into the application based on variable attributes (e.g. value ranges), system logic (e.g. sequential visit dates) and variable logic (e.g. onset date must be before cessation date). Visual review and data responses will be overseen by a trained data manager.

The database will be locked when all the expected data has been entered into the application, all query responses have been received and validated, the designated data have been noted as monitored in the system.

The data management processes are outlined in the project specific Data Management Plan (DMP); this and all related documentation are on file at McDougall Scientific and are identified by the project code AH01DEK.

#### 5.2. Coding

The adverse experiences are coded in MedDRA. All concomitant medications are coded using WHO Drug and reviewed and signed off prior to data base lock.

#### 5.3. Missing Data

The original SAP proscribed no imputation for any missing covariates and last observation carried forward for missing outcomes. The current plan still maintains no imputation for missing covariates, and further states no imputation for missing outcomes.

The primary endpoint (MADRS remission at Week 12 or study exit) will account for subjects who withdraw from the study early by using the last available MADRS score prior to withdrawing; subjects who do not have a study exit visit (e.g., those lost to follow-up) will be treated as truly missing. Clarification note study exit is defined as any visit with a MADRS *after* screening. To further clarify in SAP 2.0, this endpoint (week 12 or study exit) has not changed from the original SAP; the only update is that there is no analysis carrying forward earlier MADRS scores, but rather study exit MADRS is taken as the endpoint.

We have added several analyses which will serve to better contextualize this primary outcome and assess the robustness of findings:

- As a sensitivity test, we will conduct both best case (assuming all dropouts are remitters) and worst case (assuming all dropouts are treatment failures) analyses. The latter analysis was already pre-specified, see below.
- We will conduct an imputation using MICE as an exploratory analysis, imputing missing MADRS scores for a) the patients who were screened and eligible and b) the patients who had a least a visit 1.
- We will also examine the completer sample (those patients who completed visit 5)

### 6. Change to Analysis as Outlined in the Protocol

**Due to no remitters on MADRS in the control group, we will need to use Fisher's exact rather than regression tests. In addition, we cannot control for clinician type as only specialized care clinicians were recruited. Further sensitivity analyses were added in order to better understand the data given these changes and are detailed below.**

### 7. Statistical Methods

#### 7.1. Study Populations

The patient population for this study is comprised of adults (age >/= 18) of both sexes and any gender with a diagnosis of major depressive disorder of at least moderate severity (20 or more points on the MADRS). Patients may have psychiatric comorbidities, except for bipolar disorder of any type, but must have in addition to psychiatric comorbidity an active major depressive episode which can be clearly distinguished from other psychopathologies. In addition, patients must be in an active phase of treatment which means they must require a change in or initiation of treatment. Patients must be safe, at baseline, to be treated as outpatients.

##### 7.1.1. Physician Set

The Physician Set consists of all physicians who were randomized and enrolled at least one patient during the study. The Physician Set will be used for any summaries of physician-level information, such as physician post-interview questionnaires and physician end of study questionnaires.

##### 7.1.2. Safety Patient Population (SAF)

The SAF population consists of all subjects who provided informed consent and attended Treatment Visit 1 (Day 0). This population will be used for all other summaries and analyses. Patients will be summarized according to the treatment arm of their physician.

#### 7.2. Calculated Outcomes

The following are key endpoints derived from data captured on the CRF. Complete documentation of the calculations and data manipulation required to go from the CRF database to the analysis database are contained in the companion document - the study Analysis Programming Requirements (APR).

**Baseline Value:** Baseline values for any measurement are defined as the last non-missing measurement up to (and including) the date of Treatment Visit 1 (Day 0).

**Change from Baseline:** Value at time point – Baseline value.

**Percent Change from Baseline:** 100* [Change from Baseline]/ [Baseline value].

**Time in Trial (Days) [Physician]:** [Date of study completion or withdrawal] – [Date of randomization] + 1.

**Time in Trial (Days) [Patient]:** [Date of last visit or study withdrawal] – [Date of informed consent] + 1.

**Time on Treatment (Days):** [Date of last visit or study withdrawal] – [Treatment Visit 1/Day 0 date] + 1.

**Treatment Emergent Adverse Events (TEAEs):** An adverse event report where the time of onset is on or after the date/time of Treatment Visit 1 (Day 0).

**MADRS Remission**: Remission on the MADRS scale is defined as a total score of 0 to 10. Clarification note: calculated based on the latest MADRS available after screening.

**QIDS-SR16 Remission:** Remission on the QIDS-SR16 scale is defined as a total score of 0 to 5.

DocuSign Envelope ID: 29EC5C33-90E5-4076-9DE0-2CC10407A41A

**Treatment response:** For both the MADRS and the QIDS-SR16, treatment response is defined as a score reduction of at least 50% from baseline.

**Time to Remission:** [Date of first MADRS/QIDS-SR16 assessment showing remission] – [Treatment Visit 1/Day 0 date] + 1. If remission is not achieved, the time-to-event data are considered censored for this patient and the time to censoring is identical to Time on Treatment.

**Time to Response:** [Date of first MADRS/QIDS-SR16 assessment showing response] – [Treatment Visit 1/Day 0 date] + 1. If response is not achieved, the time-to-event data are considered censored for this patient and the time to censoring is identical to Time on Treatment.

**Number of Treatment Switches During Study:** This is defined as the number of visits after Treatment Visit 1 for a specific patient where the physician selects a new depression treatment.

**Number of Completed Outpatient Visits:** This is defined as the number of visits on or after Treatment Visit 1 for a specific patient, including unexpected visits.

##### 7.3. Interim Analysis

There are no interim analyses planned for this study.

##### 7.4. Analysis Methods

All programming will be performed using SPSS, R and Python. Continuous data summaries will consist at minimum of N, mean, standard deviation, median, and range. Categorical data summaries will be presented as counts and percentages or proportions.

Primary analyses will be focused on individual-level data, with Intra-Cluster Correlation (ICCs) calculated to examine cluster (physician) effects.

SAP V2.0 – removed section on normality assessment.

#### 8. Results

##### 8.1. Study Subjects

All data collected will be at a minimum listed. To clarify in SAP 2.0: all data here refers to all data captured in the EDC. Data from the Aifred Application pertaining to questionnaire responses will also be listed, but there is some data, for example access logs, which would not be easily understandable as a listing, but which may be used for some secondary analyses; this data will be described when used for analyses but will not necessarily be listed.

Summarization of continuous data will include at minimum N, mean, median, standard deviation, minimum, and maximum. Summarization of categorical variables will consist of the count and percentage or proportion of subjects or physicians in that category.

##### 8.1.1. Physician Disposition

All randomized physicians will be accounted for.

Physicians' time in trial and completion status (completed study or early withdrawal) will be summarized by treatment group; early discontinuations will be summarized by primary reason for discontinuation.

##### 8.1.2. Patient Disposition

All patients who provided informed consent will be accounted for.

Patient's time in trial, time on treatment, completion status, and primary reason for early discontinuation will be summarized by treatment group. The number of patients who did not pass post-screening eligibility and the number who provided informed consent but did not attend Treatment Visit 1 will also be summarized.

##### 8.1.3. Patient Characteristics

###### 8.1.3.1. Baseline Characteristics

Select demographics will be summarized for the safety patient population: age; sex; gender; living area; ethnicity; race; education level; employment; marital/relationship status; social network supportiveness rating; loneliness rating; smoking status and number of packs per day, and family history of psychiatric/mental health problems or suicide (yes/no). Baseline vital signs (height, weight, BMI, blood pressure, heart rate, systolic and diastolic blood pressure) as well as baseline scores on the MADRS, QIDS-SR16, BARS, Patient Health Questionnaire (PHQ-9), Generalized Anxiety Disorder Questionnaire (GAD-7), and Adverse Childhood Experiences Questionnaire (ACE) will also be summarized. Information regarding the current diagnosis of depression (whether it is a new diagnosis, patient/physician relationship, and stage of depression treatment) will also be summarized. These summaries will be by treatment group as well as overall. We will summarize the MINI summary as well as the demographics: Age, Sex, Education, Income, Ethnicity, whether this is a new onset or recurrent depression, Baseline MADRS severity, Depression duration (current episode). Total number of prescribed medications at baseline (total medications, not just depression), Total depression treatment trials for this episode prior to baseline

###### 8.1.3.2. Medical History

Medical history, surgical history, trauma/abuse history, psychiatric history, hospitalization history, suicide attempt history, and lifestyle questionnaire responses will each be separately listed by physician and patient. Number of comorbidities as per the MINI, Number of each type of comorbidity as per the MINI (focus on anxiety), Number of medical comorbidities (based on medical history)

###### 8.1.4. Mini International Neuropsychiatric Interview (MINI)

MINI responses will be listed by physician and patient. Each section of the MINI questionnaire will be provided in a separate listing in order to allow easier reference.

#### 8.2. Efficacy Outcomes

##### 8.2.1. Primary Outcome - MADRS Score and Remission at Week 12

MADRS remission at Week 12, defined as an overall MADRS score of 0-10 at the Week 12 visit or study exit (whichever occurs first), will be summarized by treatment group. NOTE RE: CHANGE IN VERSION 1.1 AND LATER: Due to no remitters in the control group, the regression analysis originally planned has been changed to Fisher's exact test.

As a further supplementary analysis, the primary analysis will be repeated, with subjects for whom there was not a study exit visit (e.g., those who were lost to follow-up) will be treated as not having remission rather than missing. The analysis will be run otherwise identically with these subjects counted as non-missing and not reaching remission at study exit. ADDED IN VERSION 2.0: we will also perform a best-case analysis, where those lost to follow-up are assumed to be remitters.

UPDATED IN VERSION 2.0: Box-and-whisker plots of MADRS score at Week 12 by physician, organized by treatment group will no longer be performed given the expected low patient count per physician.

##### 8.2.2. Secondary Outcomes

###### 8.2.2.1. MADRS Scores, Remission, and Response

MADRS overall scores, change from baseline, and percent change from baseline at each scheduled visit will be summarized by treatment group. Additionally, number of subjects who achieved MADRS remission (overall score 0-10) and response (at least 50% reduction from baseline) at that scheduled visit will be summarized by treatment group.

CHANGED IN VERSION 2.0: Fisher's exact test will be used to examine treatment group differences in MADRS remission at Week 4-6, and Week 8.

CHANGED IN VERSION 2.0: Cox modelling for MADRS removed, given no remitters in the control group.

Plots of mean overall MADRS score over time by treatment group will be provided for the Safety population.

CHANGED IN VERSION 2.0: Linear mixed effect models will be used for MADRS total score to assess for effect of group. Baseline MADRS will be included as a covariate (fixed) and individual physician will be included as a random effect. A random intercept will be used.

DocuSign Envelope ID: 29EC5C33-90E5-4076-9DE0-2CC10407A41A

CHANGED IN V2.0: Differences in change slopes will be carried out using the MADRS change in score divided by number of weeks in the study and compared between groups.

###### 8.2.2.2. QIDS-SR16 Scores, Remission, and Response

QIDS-SR16 overall scores, change from baseline, and percent change from baseline at each scheduled visit will be summarized by treatment group. Additionally, number of subjects who achieved QIDS-SR16 remission (overall score 0-5) at that scheduled visit will be summarized by treatment group. ICCs and 95% confidence intervals will be calculated for QIDS-SR16 score and remission at each scheduled visit.

MOVED HERE AND MODIFIED IN VERSION 2.0: A simple logistic regression will be used to measure effect of group on QIDS remission if there are at least 5 remitters per group. A mixed effect logistic regression will be used to model QIDS remission if there are at least 8 remitters in each group, with treatment group as the fixed effect variable of interest, with baseline QIDS score as a fixed effect. (In previous versions, we included clinician type, but this is no longer relevant as there was only psychiatry type clinicians in the study), and a random intercept for physician. Odds ratios of remission by treatment group will be presented with 95% confidence intervals and *p*-value. Odds ratios, 95% confidence intervals, and *p*-values will be presented for baseline QIDS score. ICC and 95% confidence interval will be calculated for QIDS score at Week 12 and QIDS remission (reported as proportion by physician) at Week 12.

MODIFIED IN VERSION 2.0: Individual-level analyses have been shown to underestimate p-values if there are fewer than 40 clusters in a cluster randomized trial (Leyrat, Morgan, Leurent, & Kahan, 2017). Data permitting (e.g. if there is not a significant proportion of no remitters for most clinicians) when the data are final, if there are 20 or fewer physicians in each treatment arm, a two-stage cluster-analysis model will be used instead. This two-stage analysis will be performed by first calculating the proportion of remissions by physician, then analyzing proportions by physician using a generalized linear model with a logit link; covariates for this model will include treatment group, weighted by number of subjects under the physician. In the case that there are more than 20 physicians in each treatment arm and the mixed-effect logistic model is used as the primary analysis, this two-stage cluster-analysis model will be performed as a supplementary analysis.

The same logistic regression model as defined above will be used to examine treatment group differences in QIDS-SR16 remission at Week 2, Week 4-6, Week 8, and Week 12/study exit. This model will also be used for QIDS-SR16 response at all scheduled visits.

Data permitting, estimates from a Cox proportional hazard model with a shared frailty term to account for physician clustering will be used to obtain treatment effect and 95% confidence intervals for time to QIDS-SR16 remission and time to QIDS-SR16 response. Kaplan-Meier plots for QIDS-SR16 remission and QIDS-SR16 response will also be provided. Kaplan-Meier plots for QIDS-SR16 remission and QIDS-SR16 response will be provided.

CHANGED IN VERSION 2.0: Linear mixed effect models will be used for QIDS total score to assess for effect of group. Baseline QIDS will be included as a covariate (fixed) and individual physician will be included as a random effect (specifically, a random intercept).

CHANGED IN VERSION 2.0: Differences in change slopes will be studied in a linear mixed effect model (physician as a random intercept); this outcome is computed using the QIDS change in score divided by number of weeks in the study and compared between groups.

###### 8.2.2.3. WHODAS Scores

Overall WHODAS scores and scores for each WHODAS domain will be presented for baseline and at Week 12/study exit. Change from baseline and percentage change from baseline will be summarized for overall and domain scores at Week 12/study exit. ICCs and 95% confidence intervals will be calculated for WHODAS score at each scheduled visit. Groups will be compared.

###### 8.2.2.4. Body weight and BMI

Body weight and BMI will be summarized at every scheduled visit.

###### 8.2.2.5. Medication and Dosage

The new treatments prescribed by study physicians—as recorded in the EDC—during the study will be listed.

###### 8.2.2.6. Completed Outpatient Appointments

The number and percent of completed outpatient appointments (as defined in Section 7.2) will be summarized overall by treatment group.

###### 8.2.2.7. Treatment Switches

The number of treatment switches (as defined in Section 7.2) will be summarized by treatment group as the number and percentage of subjects who experienced a given number of switches. The number and percentage of subjects experiencing a treatment switch at each post-baseline visit will also be presented by treatment group.

###### 8.2.2.8. Treatment Adherence

Each of the questions of the BARS will be summarized and compared by treatment group and by visit.

###### 8.2.2.9. CDA Evaluation

The responses to the questions from Sections 2 and 3 of the Post-Appointment Questionnaire (except the ones with a free-text answer) will be summarized by visit for physicians in the CDA treatment group.

###### 8.2.2.10. Therapeutic Relationship

The results from each question and the overall score for the Scale to Assess Therapeutic Relationship for the Patient (STAR-P) and the Clinician (STAR-C) will be summarized and compared by treatment group. Additionally, the individual questions from the Patient End of Study Questionnaire will also be summarized by treatment group.

###### 8.2.2.11. Further exploratory analyses

- Completion rate of questionnaires (PHQ9, GAD7, PRISE, FIBSER, QIDS) Overall and week by week (combined population and also between group)

- Doctor and patient use of different pages in app (flow/Sankey, duration spent on each page per visit, number of times visited app per week); proportion of active doctors who a) accessed the CDA b) accessed algorithm and c) saw AI results at each visit.

- Exploratory analyses of PHQ-9, GAD7, PRISE, FIBSER, totals over time

- Summarization of baseline questionnaires from CDA (AUDIT, DAST-10, SAPAS)

#### 8.3. Safety Outcomes

The safety profile will primarily be analysed by means of descriptive statistics and qualitative analysis. All summaries and listings in the section are based on the Safety population.

##### 8.3.1. Adverse Experiences

The adverse experiences/events reported will be summarized using the counts of patients and also the number of reports, for each System Organ Class (SOC) and Preferred term. The presentations will separate out and highlight any Serious Adverse Experiences (including deaths) and adverse events leading to discontinuation from the study.

The summaries will at a minimum be: 1) the number of patients reporting for all events; 2) the number of reports for all events; 3) the number of patients reporting treatment-related events (possible, probable, and/or definite relationship); 4) the number of reports of treatment (drug and device)-related events.

A by-patient listing of all adverse events will also be provided.

##### 8.3.2. Hospitalizations, ER Visits, and Suicide Attempts

The number of hospitalizations, ER visits, and suicide attempts will be summarized by treatment group. Hospitalizations and ER visits will also be summarized by reason (Psychiatric condition, medical condition, or other). Suicide attempts will be further summarized by whether they resulted in hospital or intensive care unit admission, and whether they were related to a psychiatric condition.

##### 8.3.3. Concomitant Medication

The concomitant medications, which are those with a start time/date on or after the first dose of study medication, will be listed by patient; the reason for the medication as well as the start and stop date/time will be presented.  The study times will be time from first dose of study medication.

##### 8.3.4. Vital Signs

Vital signs (heart rate, systolic blood pressure, and diastolic blood pressure) will be listed.
