## Supplementary for "Artificial Intelligence in Depression – Medication Enhancement (AID-ME): A Cluster Randomized Trial of a Deep Learning Enabled Clinical Decision Support System for Personalized Depression Treatment Selection and Management"

Supplementary Methods Required by CONSORT checklist:

This study is reported as per the CONSORT-AI checklist [32] and was conducted in accordance with all relevant ethical regulations including the Declaration of Helsinki and the Tri-Council Policy Statement. The research ethics board of the Douglas Research Center gave ethical approval for this work and it was subsequently approved by central and local ethics review boards for each site. The study was conducted in accordance with Good Clinical Practice. Written informed consent was obtained from all study participants.

*Design*

This was a two-arm, cluster-randomized trial, with clinicians serving as the cluster. Clinicians were allowed to recruit a maximum of 10 patients in order to reduce the impact of within physician intra-class correlation on inference. The expected cluster size was 7. Clinicians rather than patients were randomized as they were the ones receiving the decision support intervention, and to avoid contamination [33]. Patients entered the intervention arm of their treating clinician. No changes were made to study design after study initiation other than 1) to allow clinicians to participate longer than the originally planned 9 months in order to reach recruitment targets and 2) once the need for early study termination was determined, all participants currently enrolled were invited to complete exit interviews rather than being randomly selected.

*Participants - Clinicians*

Clinicians were recruited by the site primary investigator and treated as research participants for the purpose of the study, signed consent forms, and were overseen by site primary investigators. Clinicians could include primary care doctors, psychiatrists, residents overseen within their primary residency program by a participating clinician, nurse practitioners (with or without specialized mental health training), or nurse practitioner students overseen by a participating nurse practitioner. They needed to see at least one patient with depression per month, on average, before study start.

*Participants - Patients*

Patient recruitment criteria were intended to be broad in order to replicate a naturalistic outpatient depression population with moderate to severe depression. This was done as the CDSS is intended to be helpful at scale. Patients were recruited from the practices or hospital-based clinics of the participating clinicians. Inclusion criteria were as follows: 1) age 18 and over 2) diagnosed by their treating clinician with MDD using DSM-5 criteria [34] 3) MDD diagnosis confirmed via a blinded rater who completed the Mini Neuropsychiatric Interview (MINI) [35] and 4) at least moderate severity, as assessed by a blinded rater completing the Montgomery Asberg Depression Rating Scale (using a cutoff of 20) [36]. Patients must have been able to 5) provide their own informed consent and 6) needed to agree to be treated by their clinician for depression, understanding that they might use a range of approved treatments which might be presented in the CDSS, and understanding they were able to provide or withhold consent for any particular treatment. Contraception was used as per usual clinical practice. Exclusion criteria were as follows: 1) age under age 18; 2) presence of bipolar disorder of any type (e.g.

by clinician diagnosis or identified on the MINI) or in the patient history (except in cases where the history of bipolar disorder in the medical record was vague and was not confirmed by repeat clinical assessment or the MINI); 3) inability or unwillingness to give informed consent; 4) inability to manage patient safely as an outpatient (importantly, patients with suicidal ideation who were deemed safe to be managed as outpatients by the treating clinicians were eligible); 5) an active major depressive disorder was not the main condition being treated; and 6) an inability to use the tool (e.g. because of severe cognitive impairment). An active major depression meant that the depression, in the judgment of the treating clinician, required an initiation or a change in treatment. In addition, psychiatric comorbidities (aside from bipolar disorder) were permitted. There were no inclusion or exclusion criteria related to input data to the AI.

*Settings*

Eligible settings included any public or private outpatient setting in the United States or Canada which provided outpatient care for patients presenting with MDD, with the exception that highly specialized settings where treatments were generally pre-determined (such as ketamine clinics) were not approached for participation. Both primary care and psychiatric services were invited to participate. Given these broad requirements, a diverse array of sites joined the study. These included public sector psychiatric clinics in Canada and university-affiliated and Veteran's Affairs mental health services in the U.S. Participating sites included: the Douglas Mental Health University Institute, the McGill University Health Centre, the Jewish General Hospital, the Centre for Addiction and Mental Health, Michigan University, Emory University, the Salem VA Health Care System and VA Connecticut Health Care.

*Intervention - Aifred CDSS*

The Aifred CDSS platform consists of the following elements. The *patient portal*, accessible by web browser or mobile phone application, allows patients to complete questionnaires, receive email reminders to complete questionnaires, visualize questionnaire scores and interpretations, and track treatments which they or their clinicians enter. The *clinician portal*, accessible by web browser, allows clinicians to see all the information patients enter, while at the same time giving them access to the *clinical algorithm* module. This module is a rule-based decision tree based on the CANMAT 2016 guidelines for depression treatment [15]. This module presents the clinician with patient-specific, guideline derived information about treatment options based on the patient's depression severity, change in depression severity over time (measured using the Patient Health Questionnaire (PHQ-9) [37], and current treatments. For example, if a patient has been on a treatment that has not resulted in early improvement (25% decrease in baseline PHQ-9 score) after four weeks, the algorithm will remind clinicians that the guidelines recommend treatment switch (or augmentation if, for example, this is not the first treatment trial). The algorithm provides new information at each patient visit, based on patient progress. The clinician portal and its clinical algorithm are mainly focused on solving the treatment management problem and do not utilize AI. The AI component is focused on assisting with treatment *selection* by generating remission probabilities for 8 commonly used first line antidepressants (Citalopram, Paroxetine, Duloxetine, Venlafaxine, Fluoxetine, Bupropion, Sertraline and Escitalopram) and two commonly used combinations of first line antidepressants (Venlafaxine-XR plus Mirtazapine, and Escitalopram plus Bupropion). At the point where the clinical algorithm presents a page for the clinician to select treatment, these probabilities are displayed and, in accordance with clinician feedback during development, the treatments are ranked in order of their probability of remission. All other treatments present in the guidelines which do not have treatment probabilities associated with them are also presented on the page; these were treatments for which no or insufficient training data was available. The AI is a deep learning model trained on 9042 patients from depression treatment trials, with remission as the training objective. It takes symptom and demographic questionnaire-based data as inputs, with both the clinician and the patient each completing a short, dedicated questionnaire at the beginning of treatment to provide this data (the AI-related or “custom” questionnaires). As the AI makes predictions only for the initial treatment chosen during the study, it takes in only data at baseline. When responding to these questionnaires, full responses are required, meaning that there was no missing data when AI predictions were made; all questions were previously validated standardized questions, a decision investigators made in order to improve data reliability. The input data is based on feature selection performed during model training ([38], Benrimoh et al., 2024). Patients responding to the patient version of the custom questionnaire did not require any expertise, as they were responding to questions previously validated for patient self-report. Clinicians responding to the clinician version of the custom questionnaire (which consisted of 6 questions from the HAM-D scale and one from the HAM-A scale [39] were provided with interview guides and written prompts to help them ask and respond to these questions, but in keeping with the naturalistic design of the trial were not provided separate training on how to administer these questionnaires. The model predictions are intended to be used to inform the first treatment initiation or change in the study; the model does not update predictions after treatment failure. In order to improve interpretability, a list of the 5 most important item responses for each treatment's prediction are displayed. In order to help the clinician understand the remission probability in context, they are provided with the baseline probability of remission in the training data; the patient's mean probability of remission across predicted treatments (i.e. the average of the 10 predicted probabilities), which provides a sense of initial overall treatment success likelihood for the patient; and the relative increase or decrease relative to the patient's mean remission probability for each treatment (which helps clinicians get a sense of the ranking of each treatment relative to the others). The choice to present remission *probability*, rather than a class prediction (e.g. remit or non-remit) was made in order to provide clinicians with more nuanced information and avoid an overly-prescriptive approach to AI that would infringe on clinician autonomy[40]. The version of the AI model used in this study is extensively described in these publications ([38], Benrimoh et al., 2024).

*Intervention - Patients*

All patients received access to the same patient portal of the CDSS, where they were able to respond to questionnaires, track their responses over time using graphs, and enter and track their current and past treatments. Patients were trained to use the platform by study staff Patients did not have access to the AI or the clinical algorithm. Patient experiences differed only in their interaction with their clinicians, who had different information available based on their group assignment. Patients remained in the study for 12 weeks from their first treatment visit, and were required to see their clinician, in person or via telemedicine, at week 2, weeks 4-6, week 8 and week 12.

*Intervention - Clinicians*

There were two intervention groups: an Active group and an Active-Control group (hereinafter referred to as the Active-Control group)[33,34]. The Active-Control group was provided with all the tools required to perform best-practice measurement-based and guideline-informed care[15]. Clinicians in the Active-Control were provided with the results of questionnaires patients completed as well as training on the guidelines[15]. Guideline training involved a powerpoint presentation by DB on the guideline document as well as provision of the clinician with a copy of the CANMAT guidelines. Active-Control group clinicians were not required to use the information they were provided in any specific manner, in keeping with the naturalistic objectives of the study.

Active group clinicians received guideline training, and were provided with full access to the clinician portal of the Aifred CDSS. They were provided with training on the CDSS and on how to interpret AI results (i.e. probabilities of remission). They were instructed to consider their clinical judgment and the limitations of the AI (for example, that it only considered the data provided in the AI-related questionnaires; that it did not adapt after treatment failure; that the training data had limited demographic features available, meaning that a thorough assessment of social determinants of health was warranted for every patient; and that the model, consistent with the training data, would usually rank escitalopram as the most effective treatment, while providing more variable rankings for the other treatments (see[30]) when making treatment decisions. Active clinicians were also not required to use the information provided to them or to adhere to the AI's predictions or to the guideline information provided by the clinical algorithm. They were required to at least log in to the CDSS at each visit. While clinicians were not removed from the study if they failed to log in at each visit, they were reminded to do so.

As all raw data provided to clinicians was the same in the Active and Active-Control groups, the only group differences consisted of the provision of the data processed by the clinical algorithm and AI model to the Active group. In addition, the Active-Control group was given the tools to approximate the clinical algorithm as they were trained on the guidelines and provided with regular questionnaire data.

*Measures*

At baseline, patients were asked to complete a demographics questionnaire as well as several clinical questionnaires. These included the Mini International Neuropsychiatric Interview (MINI) and Montgomery–Åsberg Depression Rating Scale (MADRS), assessed by the blinded rater; Patient Health Questionnaire (PHQ-9), self-report Quick Inventory of Depressive Symptomatology (QIDS-SR-16, depression), General Anxiety Disorder (GAD-7, anxiety), Alcohol Use Disorders Identification Test (AUDIT, alcohol use disorder), Drug Abuse Screen Test (DAST-10, drug use), Self-Administered Standardized Assessment of Personality – Abbreviated Scale (SAPAS-SA, personality disorder screening), Adverse Childhood Experiences (ACE ), Life Events Checklist for DSM-5 (LEC-5) (both assess trauma history), and the World Health Organization Disability Assessment Schedule (WHODAS V 2.0, disability assessment) all via self-report. They were asked to complete the patient AI-related custom questionnaire no more than 2 weeks prior to their first visit, in order to ensure the AI results were reflective of their current condition. They were also asked to complete a PHQ-9 and GAD-7 weekly once they had accounts on the CDSS.

The MADRS was administered by the blinded rater at screening (no more than 2 weeks prior to the first treatment visit), visit 3 (weeks 4-6 of treatment), visit 4 (week 8) and visit 5 (week 12). Trained study staff also administered the Brief Adherence Rating Scale (BARS) after every visit to assess treatment adherence [35].

Clinicians were asked to complete the clinician version of the AI "custom questionnaire" at visit 1 (on paper in the Active-Control, and in the CDSS in the Active group). All clinicians were asked to complete a post appointment questionnaire within 48 hours of each patient visit detailing patient safety information, their assessment of patient status, any treatment changes made, and, for Active clinicians, their impression of the CDSS.

*Outcomes*

The pre-specified primary outcome of the study was remission of depressive symptoms, defined as a score of <11 [36,37] on the MADRS at study exit for those patients with at least two MADRS scores.Remission was chosen as the main outcome as it is the outcome which guidelines recommend [15]. Safety outcomes included an examination of the nature and number of adverse and serious adverse events in each group. Secondary outcomes included response (50% reduction in symptoms) on the MADRS, rate of change of the MADRS score, and medication adherence using the BARS score. Subgroup analyses aimed at examining whether patients receiving AI-prediction consistent treatments had improved outcomes and other secondary and exploratory analyses are available in the Supplementary.

*Sample Size*

We proceeded with the effect size calculation for a cluster randomized trial[38]. The intracluster correlation coefficient was set at 0.05[39]. The baseline remission value was set at 35%, based on studies in similar populations[3,40]. Cluster size was set at 7, and minimum effect size to detect was set at a 20% difference in remission rate. This was intended to be a conservative estimate based on machine learning results (see[6,20–22] and previous studies using measurement based care and algorithm-guided treatment which found larger differences in remission rate[41,42]. At 90% power, these parameters generated a requirement for 47 clinicians and 325 patients. Investigators aimed to recruit 350 patients and up to 50 clinicians. No interim analyses were planned.

*Randomization, Sequence Generation, Allocation Concealment, and Implementation*

Randomization of clinicians proceeded at 1:1 to the Active and Active-Control groups. Randomization was stratified by clinician type: primary care clinicians and non-specialized nurse practitioners were coded as being "primary care" and psychiatrists and nurse practitioners with mental health specialization were coded as being "specialized care". Block randomization with a block size of 4 was used. Sequence generation proceeded using cluster randomization where clinicians were randomized and any patients allocated to those clinicians were assigned to the same arm as the clinician. This randomization was programmed and performed in SAS. Allocation was concealed until the participating clinician completed enrollment procedures by using an interactive web response system. The sequence was generated by Hong Chen at Alimentiv Inc. and then retrieved by local site coordinators after clinician enrollment who then informed clinicians of their intervention group.

*Blinding*

*Patients*

Patients were fully blinded to group assignment. They were told that they were entering a study where they would be using a new digital technology and that there were two groups. It was explained that while their experience of the platform in each group would be the same, and that they would provide the same information, the clinicians in each group would use the information in different ways. They were told that their clinician would explain how they were using the information. Clinicians were instructed not to reveal group allocations or to tell patients what the clinicians in the other group were provided with.

*Clinicians*

Clinicians were aware of their group assignments as they were the ones receiving the AI predictions and investigators judged that, at this early stage in clinical AI research, providing clinicians with fake predictions would have been ethically questionable. Clinicians were instead partially blinded in the following manner to reduce expectation bias[43]: they were not told the study endpoints, and they were not informed of the expected effect sizes of the interventions.

*Raters*

Raters who collected the primary outcome (MADRS) and conducted the MINI were blind to group allocation.

*Study Staff*

Study staff were not blind to group allocation as they needed to provide technical support to the clinicians in the study, conduct study interviews, and provide clinicians in the Active-Control with the weekly questionnaires. Researchers conducting the analysis and those supporting the sites were likewise not blinded to group allocation.

*Statistical Analysis*

Outcome data were analyzed, as prespecified in the Statistical Analysis Plan, on an intent-to-treat basis for patients who had at least two ratings of the MADRS (the Analysis set). Safety data were analyzed for the Safety population, pre-specified as all patients who attended at least the first treatment visit. Missing data were not imputed. Analyses were carried out using SPSS (IBM) version 29.0.1.1, Microsoft Excel, and R studio. Data were collected using the Aifred CDSS (Aifred Health) and TrialStat (TrialStat). Those conducting analysis were not blinded.

Demographic and baseline clinical data were summarized and are presented in Table 1. Baseline MADRS was compared between groups using one-way ANOVA. The primary outcome (MADRS remission) was assessed using a Fisher's exact test due to the lack of remitters in the Active-Control group. As pre-specified, sensitivity analyses were carried out (see Supplementary Material). Secondary outcomes were compared using one-way ANOVAs, and proportions were compared using two-sided $X^2$ or two-sided Fisher's exact tests, as appropriate. Cox models were initially to be used to assess time to remission; as this was not possible because of the lack of remitters in the control group, this analysis was replaced with an analysis of slope of MADRS change (change in score over time in study). While the analysis plan called for adjustment by clinician type, this was not relevant or necessary as only specialist clinicians were recruited. To assess patient engagement, investigators calculated the percentage of the self-report questionnaires completed by patients during the 12 treatment weeks, accounting for study dropout. Physician engagement was determined by examining clinician access logs for the platform at each visit. Secondary and exploratory analyses were not corrected for multple comparisons. Further pre-specified Supplementary analyses are detailed in the Supplementary Material.

*Early Study Termination*

Unfortunately, due to lack of funding caused by delays related to the COVID-19 pandemic, the study was terminated early.

Extended results requires for CONSORT:

*Sites*

Ten sites were recruited and were cleared to recruit patients; of these 1 site was closed early because of lack of capacity to complete the trial. 8 sites recruited patients into the study. Sites were located in Canada (5) and the United States (4) and included U.S.Veterans Affairs hospitals and mood disorders programs in university-affiliated psychiatric departments.

*Recruitment - Clinicians*

50 clinicians were recruited, consistent with the recruitment target. 26 were randomized in the Active group and 24 in the Active-Control group. 39 of these clinicians were psychiatrists; 2 were nurse practitioners specialized in psychiatry, and 9 were psychiatry residents. Of the 47 clinicians recruited who were cleared to recruit patients prior to early study termination, 25 were randomized to the Active group and 22 to the Active-Control group. 27 clinicians recruited at least one patient (57%); 16 in the Active group (64%) and 11 in the Active-Control group (50%). Active and Active-Control clinicians spent essentially the same mean number of months in the study (Active = 9.9 months; Active-Control = 10.1 months). Further details are available in the Supplementary Material.

Further supplementary data:

Further secondary outcomes are presented in the Supplementary Material. These include total score differences, remission and response on the QIDS-SR-16; functional outcomes, as measured by between group differences in final WHODAS scores; clinician engagement with the CDSS as assessed by clinician responses to questions on the end and post-appointment questionnaires and clinician behavior in the application; patient engagement as assessed by patient completion rates of questionnaires in the CDSS; to evaluate service utilization by comparing number of visits and ER/hospitalization use between groups.

Any analyses specified in the SAP which were not conducted were not included for brevity, or not conducted because of lower than expected sample size or because a previous analysis did not show significance and the analysis not conducted was as a result not expected to be significant. For example, treatment change rates between groups were very similar, and treatment changes after visit 1 were very small in number, and as such it was not feasible to examine treatment change rates in AI consistent vs. AI inconsistent subgroups.

Section 1: Further results from MADRS analysis:

1.1 MADRS remission and remission sensitivity analysis

In exploratory analysis, between group differences in remission were not observed at V3 (4-6 weeks) (n = 60 (19 Active-Control), 17.1% active, 5.3% Active-Control, p = 0.42, Fisher's exact test), but became apparent at V4 (8 weeks) (n = 53 (19 Active-Control), 20.6% active, 0% Active-Control, p = 0.04, Fisher's exact test) and at V5 (12 weeks) (study completers) (n = 50 (18 Active-Control), 25% active, 0% Active-Control, p = 0.04, Fisher's exact test).

As pre-specified, sensitivity analyses were carried out assuming best and worst case outcomes for those patients who were not followed to week 5 (i.e., assuming patients who did not reach visit 5 were remitters or non-remitters, respectively).

In the worst case sensitivity analysis, assuming all patients who did not complete visit 5 did not remit, 0% of the Active-Control and n = 8 (13.1%) of the Active group remitted, which was marginally significantly different (p = 0.05, Fisher's exact). In the best case scenario, assuming all dropouts did remit, there was one remitter in the Active-Control group (5%) and 19 in the Active group (45%), which was significantly different ($X^2 = 9.5$, $p = 0.002$). As such, sensitivity testing showed consistent between group differences in remission.

1.2 MADRS scores for each visit

No significant differences were noted in the final MADRS score (active: mean = 21, SD = 12.8; Active-Control = 25.3, SD = 9.3; F = 1.8, p = 0.19, ANOVA). Significant differences in MADRS total score were similarly not observed at visit 3, 4, or 5.

| MADRS Score | Mean (SD) | F | p-value |
|---|---|---|---|
| Baseline | 32.10 (6.91) | 2.097 | 0.15 |
| Active-Control (n = 19) | 30.21 (5.75) | | |
| Active (n = 42) | 32.95 (7.28) | | |
| V3 | 25.00 (10.62) | 0.358 | 0.55 |
| Active-Control (n = 19) | 26.21 (7.92) | | |
| Active (n = 41) | 24.44 (11.7) | | |
| V4 | 23.89 (10.79) | 1.197 | 0.28 |
| Active-Control (n = 19) | 26.05 (8.28) | | |
| Active (n = 34) | 22.68 (11.91) | | |
| V5 | 22.76 (11.49) | 0.726 | 0.40 |
| Active-Control (n = 18) | 24.61 (9.06) | | |
| Active (n = 32) | 21.72 (12.67) | | |
| Last MADRS | 22.31 (11.96) | | |
| Active-Control (n = 19) | 25.32 (9.33) | 1.77 | 0.19 |

| | |
|---|---|
| Active (n = 42) | 20.95 (12.84) |

1.3 Linear mixed model predicting MADRS total score at study end

Using a linear mixed model in R per the statistical analysis plan, MADRS at study end was predicted using treatment group as a predictor, baseline MADRS as a covariate, and accounting for clinician level clustering (intraclass correlation coefficient of 0.166). In this analysis group was not a significant predictor (Bootstrap CI = -12.7, 0.52; Model 95% CI -13.6, 1.4). However, there was a -6.1 difference in score. While this was not significant, the final sample size did not reach the original target, and so the comparison may be underpowered.

1.4 MADRS scores

Figure 1.4.1 MADRS total at each time point for each participant (full sample)

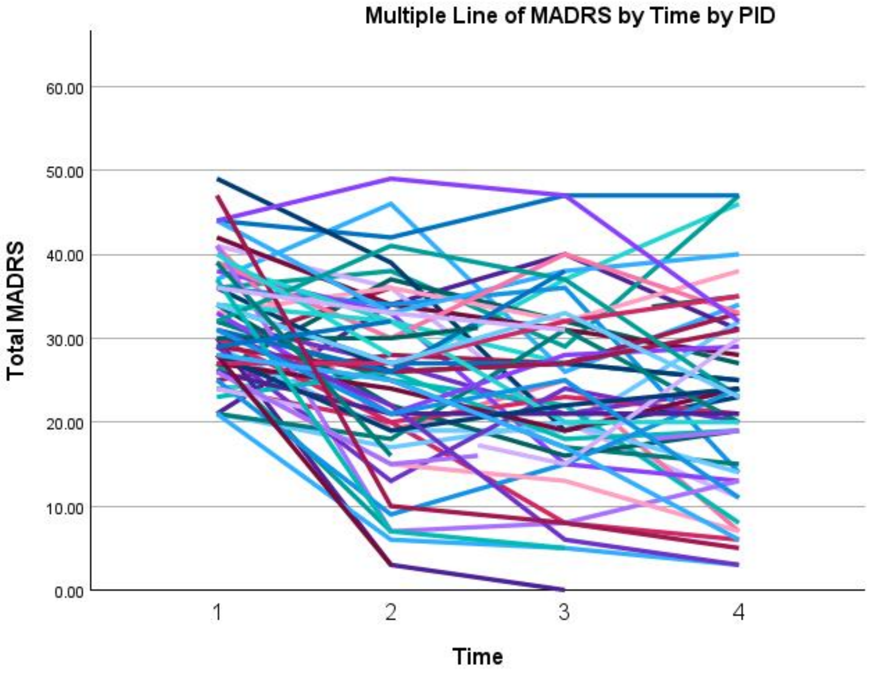

Fig 1.4.1 the total MADRS score at each timepoint for each participant (total sample). PID = Patient ID

Fig. 1.4.2 Total MADRS at each timepoint for each participant, active group

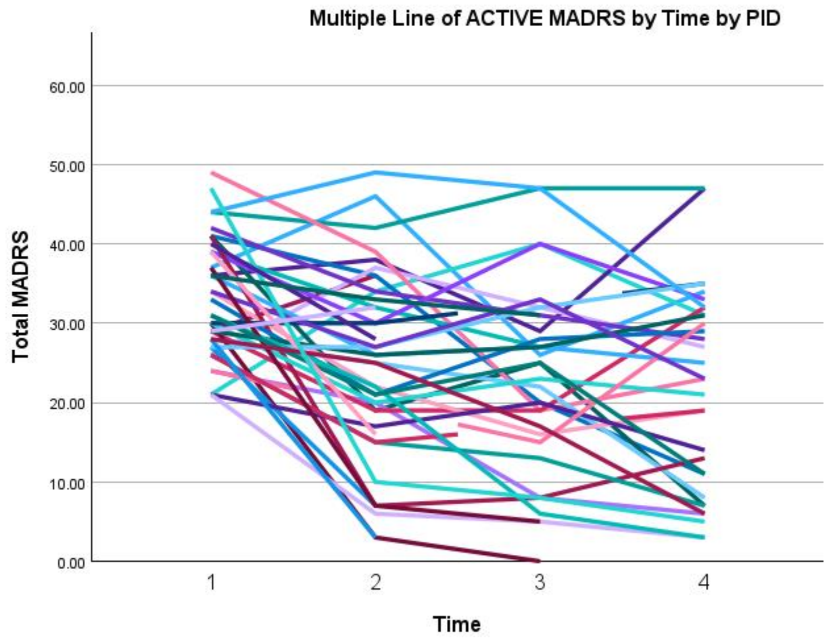

Fig 1.4.2 total MADRS score at each timepoint for each participant in the Active group. PID = Patient ID

Fig. 1.4.3 Total MADRS at each timepoint for each participant, control group

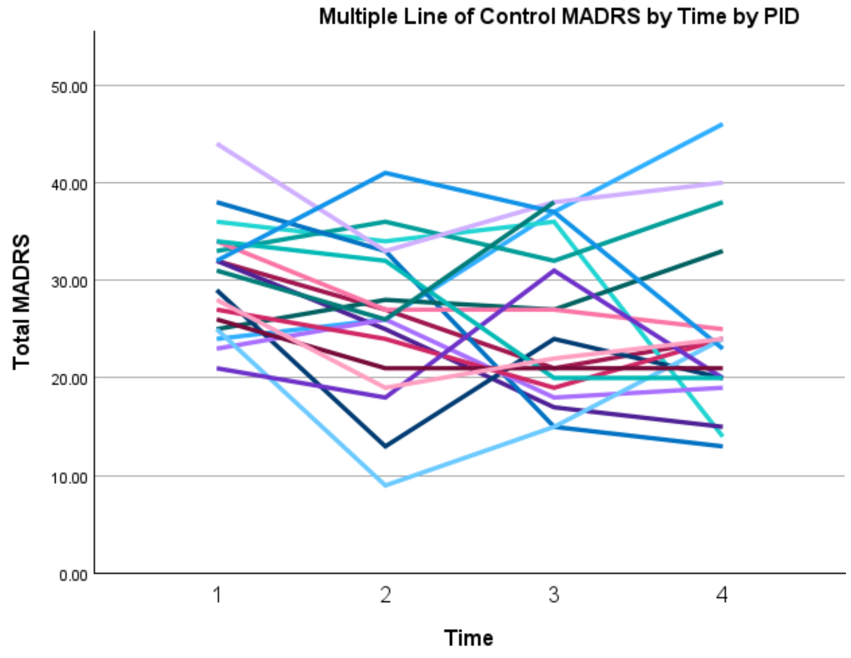

Fig 1.4.3 total MADRS score at each timepoint for each participant in the Control group. PID = Patient ID

Fig 1.4.4 Mean MADRS is plotted by group for all participants with at least one MADRS.

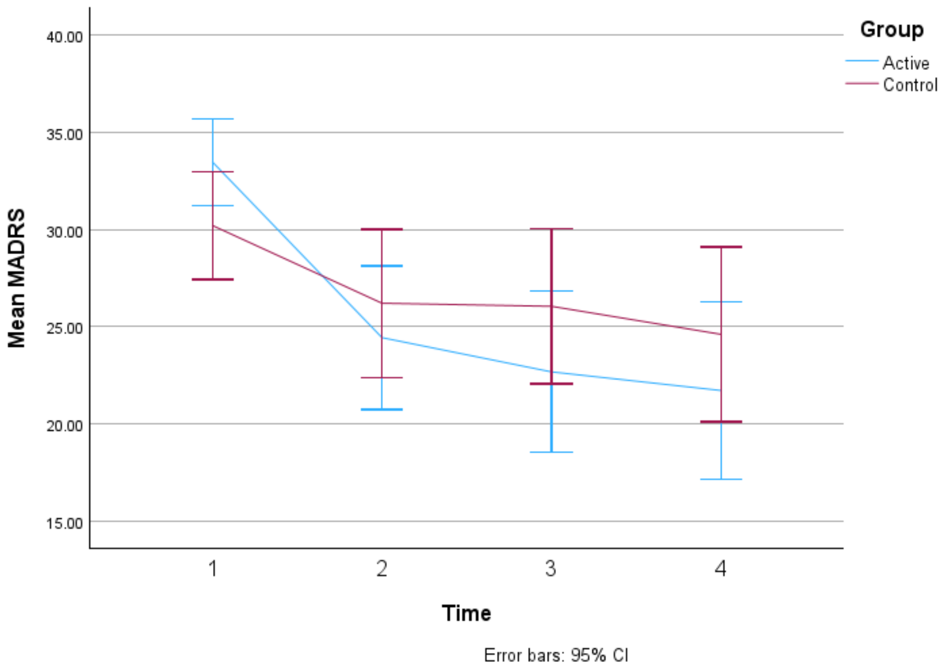

Fig 1.4.4 Mean MADRS is plotted by group for all participants with at least two MADRS (the analysis set).

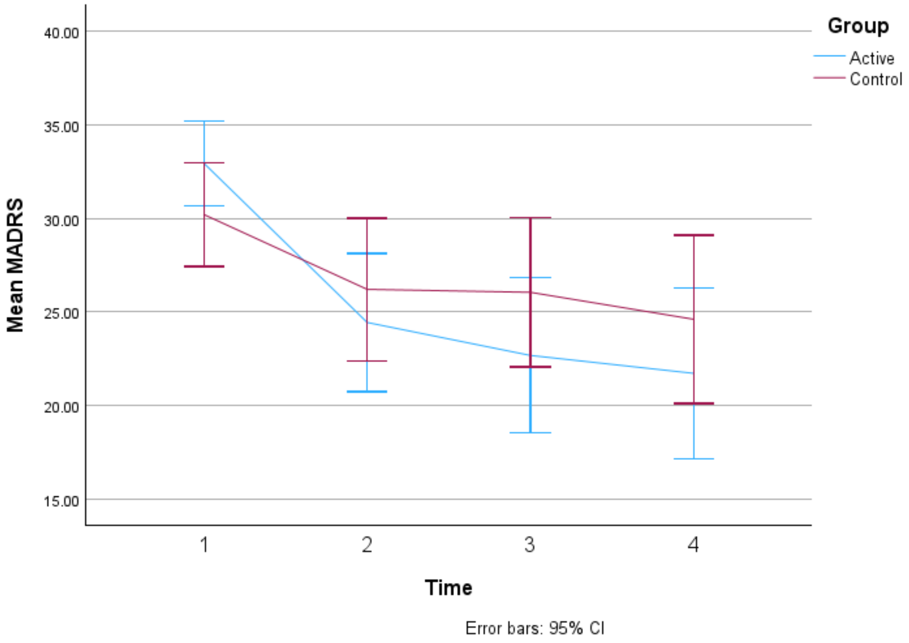

1.5 Baseline MADRS score is not different between remitters and non-remitters

To ensure that remitters in the Active group did not have lower baseline MADRS severity than other patients due to chance imbalance, investigators examined this difference with an ANOVA; no significant difference was found. Remitters (n = 12) had a baseline MADRS of 30.67 (SD = 7.44); non-Remitters had a baseline MADRS of 32.45 (SD = 6.81); this was not significantly different using ANOVA: (F(1) 0.64, p = 0.43).

1.6 Number of weeks when between first and last MADRS, by group

There was no significant difference in the number of weeks between the first and last MADRS was completed between groups (F(1) = 3.40, p = 0.07, ANOVA). Mean number of weeks in control was 12.31 (SD = 1.64) and in the active was 10.92 (SD = 3.10).

Analysis based on the visit number for patients making it to visit 4 or 5 and treatment weeks (between visit 1 and the last visit) for patients making it to visit 4 (Weeks 4-6) was also non-significant (F(1) = 3.46, p = 0.07, ANOVA). Mean number of weeks in control was 11.79 (SD = 0.92) and in the active was 10.61 (SD = 2.69).

Section 2: Data from the QIDS-SR-16

Data from the QIDS are presented as a self-report counterpart to the MADRS score. As patients were completing these questionnaires in the study application, the analysis was conducted with scores aligned to visits. Below is the analysis for the analysis set.

Baseline (first) QIDS-SR-16 scores in the Active group were (mean = 13.0238, SD = 4.4310) and in the Active-Control group were (mean = 14.0526, SD = 4.1025); this was not significantly different (t = -0.8844, p = 0.38, t-test).

2.1 For total QIDS score, there is a significant between-group difference at visit 5 only, showing reduced total QIDS scores in the Active compared to the Active-Control group

| **61 patients with at least 2 visits – analysis based on last visit and on each visit** | | | | | |
|---|---|---|---|---|---|
| **For last visit** | | | | | |
| **Data** | **N** | **Active-Control Mean** | **Active Mean** | **Active-Control SD** | **Active SD** |
| ***Last visit score*** | 61 | 14.8421 (n = 19) | 12.5 (n = 42) | 5.3257 | 5.7689 |
| **ANOVA** | **DF** | **Sum sq** | **Mean Sq** | **F value** | **p-value** |
| ***Group*** | 1 | 71.8 | 71.76 | 2.258 | 0.14 |
| ***Residuals*** | 59 | 1875.0 | 31.78 | | |
| **For each visit** | | | | | |
| **Data** | **N** | **Active-Control Mean** | **Active Mean** | **Active-Control SD** | **Active SD** |
| ***Screening*** | 61 | 14.0526 | 13.0238 | 4.1025 | 4.4310 |
| **ANOVA** | **DF** | **Sum sq** | **Mean Sq** | **F value** | **p-value** |
| ***Group*** | 1 | 13.8 | 13.85 | 0.737 | 0.39 |
| ***Residuals*** | 59 | 1107.9 | 18.78 | | |
| **Data** | **N** | **Active-Control Mean** | **Active Mean** | **Active-Control SD** | **Active SD** |
| ***Visit 1*** | 61 | 14 | 13.4524 | 5.3955 | 4.7996 |

| ANOVA | DF | Sum sq | Mean Sq | F value | p-value |
|---|---|---|---|---|---|
| ***Group*** | 1 | 3.9 | 3.923 | 0.158 | 0.69 |
| ***Residuals*** | 59 | 1468.4 | 24.888 | | |
| **Data** | **N** | **Active-Control Mean** | **Active Mean** | **Active-Control SD** | **Active SD** |
| ***Visit 2*** | 60 | 13.8945 | 13.8293 | 3.1780 | 4.9593 |
| **ANOVA** | **DF** | **Sum sq** | **Mean Sq** | **F value** | **p-value** |
| ***Group*** | 1 | 0.1 | 0.056 | 0.003 | 0.96 |
| ***Residuals*** | 58 | 1165.6 | 20.096 | | |
| **Data** | **N** | **Active-Control Mean** | **Active Mean** | **Active-Control SD** | **Active SD** |
| ***Visit 3*** | 61 | 14.9474 | 12.7381 | 6.3550 | 4.6121 |
| **ANOVA** | **DF** | **Sum sq** | **Mean Sq** | **F value** | **p-value** |
| ***Group*** | 1 | 63.9 | 63.85 | 2.356 | 0.13 |
| ***Residuals*** | 59 | 1599.1 | 27.10 | | |
| **Data** | **N** | **Active-Control Mean** | **Active Mean** | **Active-Control SD** | **Active SD** |
| ***Visit 4*** | | 15.0526 | 15.1842 | 6.3637 | 4.8483 |
| **ANOVA** | **DF** | **Sum sq** | **Mean Sq** | **F value** | **p-value** |
| ***Group*** | 1 | 0.2 | 0.219 | 0.008 | 0.93 |
| ***Residuals*** | 55 | 1598.7 | 29.067 | | |
| **Data** | **N** | **Active-Control Mean** | **Active Mean** | **Active-Control SD** | **Active SD** |
| ***Visit 5*** | 56 | 14.8421 | 11.6487 | 5.3257 | 5.1598 |
| **ANOVA** | **DF** | **Sum sq** | **Mean Sq** | **F value** | **p-value** |
| ***Group*** | 1 | 128 | 128.0 | 4.706 | 0.03 * |
| ***Residuals*** | 54 | 1469 | 27.2 | | |

2.2 *Remitters and responders on the QIDS-SR-16 for last score in the analysis set*

4 patients (9.524%) went into remission based on the QIDS-SR-16 (Remission = last score < 6) in the Active group, and 0 patients in the Active-Control group (0%). This was not statistically significant:

| **Remitters (n = 61)** | | |
|---|---|---|
| | Remission | Not Remission |

| | | |
|---|---|---|
| Active | 4 | 38 |
| Active-Control | 0 | 19 |
| **Fisher's exact test** | | |
| p = 0.31<br>95 percent confidence interval: (0.2721912, Inf) | | |

Furthermore, 4 patients (9.524%) responded based on the QIDS-SR-16 (Responders have a (first score - last score)/first score > = 0.5) in the Active group, and 0 patients in the Active-Control group (0%). This was not statistically significant:

| **Responders** | |
|---|---|
| Active | 4 |
| Active-Control | 0 |
| **Fisher's exact test** | |
| p = 0.31<br>95 percent confidence interval: (0.2721912, Inf) | |

Finally, change from baseline to last score in QIDS-SR-16 were (mean = 0.5238, SD = 7.8902) in the Active group and (mean = -0.7895, SD = 5.7887) in the Active-Control group. This was not statistically significant (p = 0.47, t-test), but indicates a numerical reduction in scores in the Active group, and an increase (negative reduction) of scores in the Active-Control group. Lastly, the difference in rate of change (mean = -0.0401, SD = 0.8367) in the Active group and (mean = -0.0702, SD = 0.4837) in the Active-Control group over time was not statistically significant (p = 0.86, t-test).

| **t-tests by group with at least 2 visits** | | | | | |
|---|---|---|---|---|---|
| **Score change from baseline by group (baseline - last score)** | | | | | |
| T stat | p-value | Mean score change from baseline in Active (n = 42) | SD of score change from baseline in Active (n = 42) | Mean score change from baseline in Active-Control (n = 19) | SD of score change from baseline in Active-Control (n = 19) |
| 0.7289 | 0.47 | 0.5238 | 7.8902 | -0.7895 | 5.7887 |
| **Rate of change from baseline by group (change from baseline / # of weeks from first-last visit)** | | | | | |

| T stat | p-value | Mean rate of change in Active (n = 42) | SD of rate change in Active (n = 42) | Mean rate of change in Active-Control (n = 19) | SD of rate of change percent in Active (n = 19) |
|---|---|---|---|---|---|
| 0.1768 | 0.86 | -0.0401 | 0.8367 | -0.0702 | 0.4837 |

Note: given a lack of significant differences in QIDS-SR-16 scores between groups on most measures and the smaller than expected sample sizes, mixed modeling and further analyses were not carried out using the QIDS-SR-16 as originally intended per the SAP given they were not a main outcome and power would have been lower than the analysis conducted above.

3. Baseline, outcomes, and changes in the WHODAS

The WHODAS served as a measure of disability in the study. Investigators examined the baseline, last scores, and the change between them in terms of both the total score and the 6 domain scores of the WHODAS. Note that while not statistically significant, the Active group experienced a small reduction in disability, and the Active-Control group experienced a small increase.

| | Active Mean (SD) (n = 28) | Active-Control Mean (SD) (n = 15) | Test Statistic | P-value |
|---|---|---|---|---|
| WHODAS Baseline total score (n = 43) | 88.6428 (26.4255) | 84.8666 (22.0417) | ANOVA | 0.64 |
| WHODAS last score (n = 43) | 80.3928 (24.7615) | 87.3333 (25.3593) | ANOVA | 0.39 |
| WHODAS change in total score (n = 43) | 8.25 (22.9356) | -2.4666 (19.8884) | ANOVA | 0.13 |
| Domain 1, last score (n = 43) | 13.7857 (4.4833) | 15.4 (4.9396) | ANOVA | 0.28 |
| Domain 2, last score (n = 43) | 10 (4.3461) | 9.6666 (3.9940) | ANOVA | 0.81 |
| Domain 3, last score (n = 43) | 6.4285 (2.3637) | 7.3333 (3.8483) | ANOVA | 0.34 |
| Domain 4, last score (n = 43) | 11.8571 (4.2226) | 12.3333 (5.5506) | ANOVA | 0.75 |
| Domain 5, last score (n = 43) | 17.6428 (8.3409) | 19.7333 (8.1105) | ANOVA | 0.43 |
| Domain 6, last | 20.6785 | 22.8666 | ANOVA | 0.31 |

| | | | | |
|---|---|---|---|---|
| score (n = 43) | (6.7057) | (6.4016) | | |
| Domain 1, change in score (n = 43) | 1.8571 (4.3266) | -0.6666 (3.8668) | ANOVA | 0.07 |
| Domain 2, change in score (n = 43) | 0.6785 (2.9944) | -1.4 (3.7186) | ANOVA | 0.05 |
| Domain 3, change in score (n = 43) | 0.5 (3.4156) | -0.4666 (2.4455) | ANOVA | 0.34 |
| Domain 4, change in score (n = 43) | 1.75 (4.1510) | -0.4666 (2.7996) | ANOVA | 0.07 |
| Domain 5, change in score (n = 43) | 2.3214 (9.2738) | 1.7333 (9.0511) | ANOVA | 0.84 |
| Domain 6, change in score (n = 43) | 1.1428 (5.4277) | -1.2 (3.7834) | ANOVA | 0.15 |

4. Clinician perception of application safety

4.1 Qualitative comments:

After *each* appointment, clinicians were asked several questions to assess their perception of the safety of the CDSS and its recommendations.

When asked: “Did the application produce any recommendations you felt were unsafe, or diverged dangerously from standard practice?” 2 clinicians responded yes; however, their responses clarify that their concerns were not related to patient safety. The first clinician wrote “The 1st choice recommended by the application not reimbursed, selected a parent compound”; likely related to the fact that escitalopram is not a covered medication in the Canadian province of Quebec. The second clinician wrote “The algorithm recommended medications she has tried during this depressive episode. Not helpful”; clinicians were trained to expect that the AI does not take previous medications into account due to lack of this information in the training data; this is indeed a limitation of the AI, but not, investigators argue, a clear safety concern. The rule-based algorithm, however, would have flagged previously tried medications if they were entered, in order to help offset this limitation.

When asked: “Did you feel any of the treatments in the treatment list were inappropriate?” 5 clinicians answered yes. One clinician did not provide further information. Two clinicians noted that they believed the application was recommending adding escitalopram to venlafaxine (which the patient was already taking). Another clinician said that they had asked for augmentation strategies but were provided with medications for switching. It is likely these clinicians were presented with a treatment selection page in the context of their patient not yet being in remission, where they were invited to review the treatment and consider augmentation; the algorithm presents the treatments for augmentation lower down the list than the first line treatments and they must be scrolled to to be accessed. The first line treatments are presented in case the clinician decides the switch. The application is not programmed to ever recommend specifically adding escitalopram to venlafaxine. In addition, if this combination had been selected, the application would have provided a warning about the risks of combining an SSRI and SNRI. As such this seems to have been an issue with the understanding of these clinicians of how the treatment selection page was laid out; future versions of the design of this page will be taken into account. Finally, one last clinician (the same clinician who responded similarly to the above question) noted: “The top 3 recommendations were medications she has tried during the depressive episode. Not helpful.” As such, while these comments demonstrate that improvements could be made to the layout of the treatment selection page, there were no clear recommendations that led to patient harm, and clinicians recognized that the recommendation would have been inappropriate, had the application actually made it, and did not implement it.

### 4.2 Quantitative Data

Investigators next examine clinician trust in the AI/algorithm predictions. After each visit, Active clinicians were asked “Did you trust the information provided by the clinical algorithm and AI model (i.e. treatment lists or remission probabilities)?”. They responded in a 1-5 likert scale (not at all to very much); investigators consider a response of 4 or 5 to be “positive” and 1-3 to be negative or neutral. Here is presented both the means of the responses per visit, as well as a summary of the responses split by positive or negative responses.

Mean and SD of responses (trust):

| | N | Minimum | Maximum | Mean | Std. Deviation |
|---|---|---|---|---|---|
| Visit 1 | 28 | 2.00 | 5.00 | 3.82 | 0.90 |
| Visit 2 | 23 | 3.00 | 5.00 | 3.74 | 0.69 |
| Visit 3 | 25 | 2.00 | 5.00 | 3.56 | 0.96 |
| Visit 4 | 23 | 2.00 | 5.00 | 3.70 | 0.82 |
| Visit 5 | 22 | 2.00 | 5.00 | 3.73 | 0.88 |

Binarized Responses (trust):

| | Visit 1 | Visit 2 | Visit 3 | Visit 4 | Visit 5 |
|---|---|---|---|---|---|
| 1,2,3 | 36% | 39% | 44% | 35% | 27% |
| 4,5 | 64% | 61% | 56% | 65% | 73% |
| Total | 100% | 100% | 100% | 100% | 100% |

By the end of the study, the numbers of clinicians indicating trust in the CDSS is consistent with the previous feasibility study, which found that 71% of clinicians expressed trust in the AI.

Investigators also asked clinicians after each appointment to indicate if they felt that “I feel that the clinical algorithm (AI model)’s results have been: 1 = very dangerous, 5 = very safe”. Using the same analysis approach as above:

Mean and SD of responses (perception of safety):

| | N | Minimum | Maximum | Mean | Std. Deviation |
|---|---|---|---|---|---|
| Visit 1 | 28 | 1.00 | 5.00 | 4.04 | 0.88 |
| Visit 2 | 23 | 3.00 | 5.00 | 4.04 | 0.77 |
| Visit 3 | 25 | 3.00 | 5.00 | 4.08 | 0.57 |
| Visit 4 | 23 | 3.00 | 5.00 | 4.13 | 0.46 |
| Visit 5 | 22 | 3.00 | 5.00 | 4.09 | 0.53 |

Binarized responses (perception of safety):

| | Visit 1 | Visit 2 | Visit 3 | Visit 4 | Visit 5 |
|---|---|---|---|---|---|
| 1,2,3 | 18% | 26% | 12% | 4% | 9% |
| 4,5 | 82% | 74% | 88% | 96% | 91% |
| Total | 100% | 100% | 100% | 100% | 100% |

As such, the vast majority of clinicians found that the CDSS produced safe results.

Finally, investigators inquired about clinician perception of appropriateness: “I feel that the clinical algorithm (AI model)’s results have been: 1 = very inappropriate, 5 = very appropriate”

Mean and SD of responses (perception of appropriateness):

| | N | Minimum | Maximum | Mean | Std. Deviation |
|---|---|---|---|---|---|
| Visit 1 | 28 | 2.00 | 5.00 | 3.79 | 0.79 |
| Visit 2 | 23 | 3.00 | 5.00 | 3.65 | 0.78 |
| Visit 3 | 25 | 2.00 | 5.00 | 3.68 | 0.85 |
| Visit 4 | 23 | 2.00 | 5.00 | 3.65 | 0.78 |
| Visit 5 | 22 | 2.00 | 5.00 | 3.77 | 0.75 |

Binarized responses (perception of appropriateness):

| | Visit 1 | Visit 2 | Visit 3 | Visit 4 | Visit 5 |
|---|---|---|---|---|---|
| 1,2,3 | 36% | 52% | 40% | 43% | 32% |
| 4,5 | 64% | 48% | 60% | 57% | 68% |
| Total | 100% | 100% | 100% | 100% | 100% |

Taken together, there were no indications in the qualitative or quantitative data of significant safety concerns related to the use of the CDSS. There were however some comments which will likely provide the substance for improving the interface in the future.

5.1 Completion rates for the PHQ-9 - first 14 weeks of application access

For the first 14 weeks of application access, the PHQ-9 completion rate was 77% (75% Active, 82% Active-Control). This difference reflects the fact that patients tend to respond to more questionnaires earlier in treatment, as can be seen below. 12 week data included for comparison

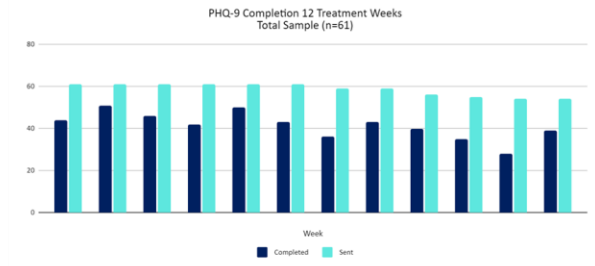

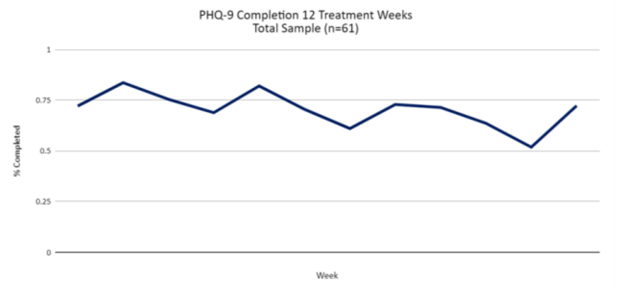

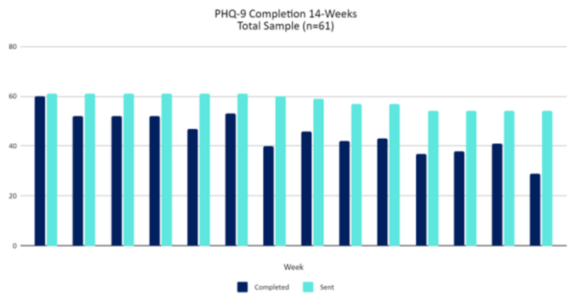

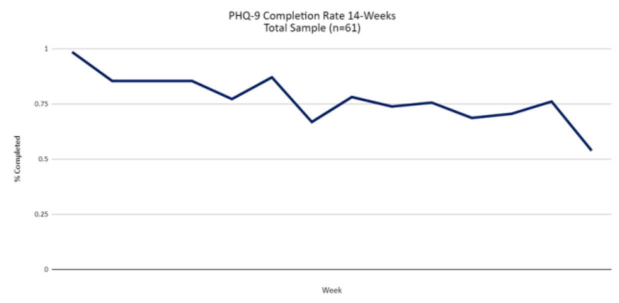

5.2 Questionnaire completion rates for other CDSS questionnaires completed by patients, expressed as a % completed of those sent

| Questionnaire (frequency) | Total -12 treatment weeks | Active - 12 treatment weeks | Active-Control - 12 treatment weeks | Total - first 14 weeks | Active - first 14 weeks | Active-Control - first 14 weeks |
|---|---|---|---|---|---|---|
| GAD-7 (weekly) | 68 | 64 | 77 | 76 | 73 | 83 |
| QIDS-SR-16 (biweekly) | 76 | 73 | 84 | 87 | 83 | 94 |
| FIBSER (biweekly) | 77 | 74 | 83 | 87 | 84 | 94 |
| PRISE20 (biweekly) | 76 | 74 | 82 | 87 | 84 | 93 |

It is interesting to note the higher completion rate for biweekly questionnaires, even though these are longer than the PHQ-9 and GAD-7. It is possible that sending more questionnaires simply means more chances for questionnaires to not be completed. Alternatively, the fact that these were sent less frequently may have made them feel more important to patients; or patients may have just felt overwhelmed with the idea of responding to weekly questionnaires. While the data available does not allow us to answer this question, and completion rates were high overall and in line with investigators' previous feasibility study, this would be an interesting avenue for future research.

### 6. Further details on clinician application use

For the 16 clinicians who had a patient at some point in the study, investigators generated a heat map which shows the number of app access per patient per week, for the weeks in which the doctor had a patient in the study; this shows that most clinicians opened the app once or twice per patient per week when they had a patient active.

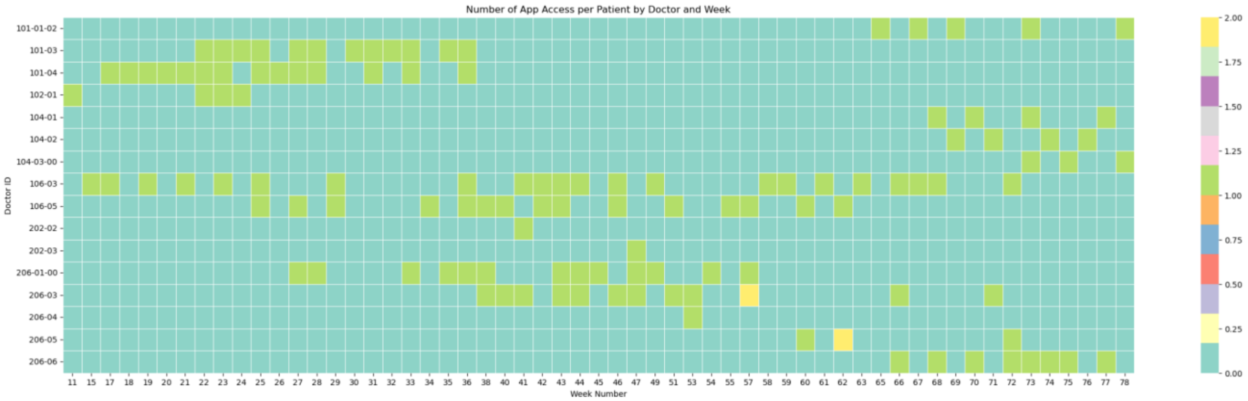

Investigators also produced a Sankey diagram which shows how clinicians used the app, demonstrating that, reasonably, most clinicians after accessing the app either reviewed patient treatment or entered the treatment algorithm as their next step; comparatively fewer accessed the detailed versions of the questionnaires, suggesting they would be focusing on the graphs of questionnaire total scores provided immediately when they accessed a patient's file (as the landing page once a patient is clicked on is the graphs page). Future research might focus on seeing if further benefit to patient care could be had by helping clinicians to understand and utilize more detailed questionnaire results. Note: the diagram represents the summed total accesses across all doctors.

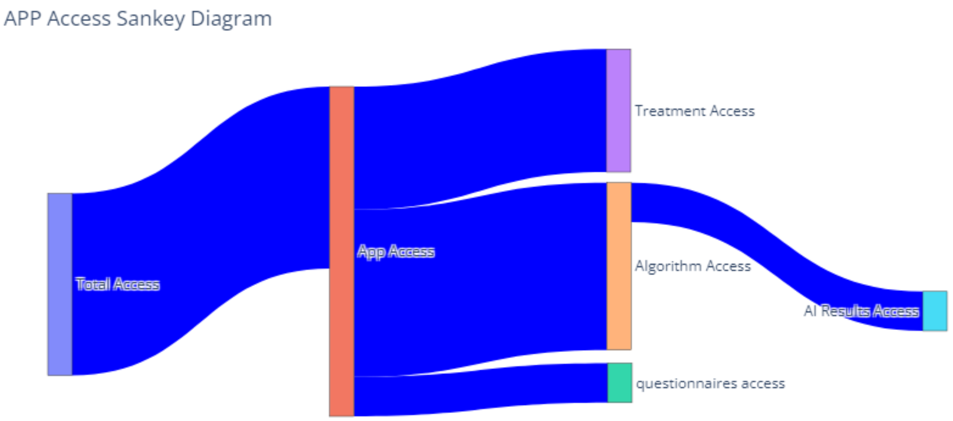

7. Treatment initiation, switches, and dose changes in the Active vs. the Active-Control group

This table lists, as totals and broken down per patient in each group, for psychiatric medications, the number of new medications started, the number of dose increases and decreases, the number of medications stopped, and the number of patients who had no change in their treatment across all treatment visits. As can be seen, the groups were comparable in terms of the numbers of changes made of each type. As such, one possibility is that the *quality* or *timing* of each decision was positively influenced by access to the CDSS, given improved outcomes in the Active group. Limitations in sample size, and patient complexity in this sample, limits investigators' ability to definitively assess this possibility.

| | Active patient N = 42 | Control patient N = 19 |
|---|---|---|
| Total count of NEW medication | 40 | 19 |
| NEW MEDICATION PER patient | 0,9523809524 | 1 |
| Total count - CHANGE OF DOSE - INCREASE | 56 | 30 |
| DOSE INCREASE per patient | 1,333333333 | 1,578947368 |
| Total count of CHANGE OF DOSE - DECREASE | 7 | 7 |
| DOSE DECREASE per patient | 0,1666666667 | 0,3684210526 |
| Total count of STOP A TREATMENT | 14 | 8 |
| STOP A TREATMENT per patient | 0,3333333333 | 0,4210526316 |
| NO NEW MED OR DOSE CHANGE | 4 | 1 |
| NO NEW MED OR DOSE CHANGE per patient | 0,09523809524 | 0,05263157895 |

Importantly, differences in treatment outcome between groups was not likely to be due to differences in psychotherapy access. As per the clinician post-appointment questionnaires, no patients were listed as being in psychotherapy before study start and only 2 Active and 4 Active-Control patients were listed as being prescribed or beginning psychotherapy during the 12 treatment weeks.

In an exploratory analysis, investigators examined the number of patients in each group who experienced a switch in their medication or the addition of an adjuvant medication after V1. 3 patients in each group had a switch in their medication (7.1% active, 15.8% Active-Control). In addition, both groups had similar numbers of adjuvant treatments added after V1 (n = 12, 28.6% active; n = 5, 26.3% Active-Control). While the numbers are too small to draw firm conclusions, there is some suggestion that initial treatment choices were more effective in the Active group; while there was no significant effect of selecting AI consistent treatment (See below), this suggests that perhaps access to the platform in general or the AI results in particular helped with treatment choice outside of providing a ranked list of treatments.

8. Aifred CDSS visuals

Here investigators demonstrate how the CDSS results are displayed to clinicians. First, patient questionnaire results are provided in a graphs page. Within the clinical algorithm, the interface provides the name of the drug, information about dosing, clinical pearls derived from the literature (mostly from the CANMAT guidelines), as well as the results of the AI. These AI results are provided in purple to differentiate them from other, literature-derived information. The treatments, per requests from clinicians in previous studies, are presented in an ordered list based on the probability of remission. Raw probabilities, rather than class labels, are provided in order to avoid being overly prescriptive and to encourage clinicians to think of the prediction as ‘one more piece of information’ rather than as a directive of what to do. The platform then calculates the mean remission rate over all 10 treatments predicted for this patient, and how they rank relative to each other. Furthermore, the platform provides the baseline remission rate of 43.15% in order to help clinicians get a sense of how the individual patient compares to the sample of depressed adults used to train the model. Finally, the model provides the interpretability report- the 5 features which were most predictive of the outcome for that particular treatment, alongside participant responses to the question representing the feature. In this example, a question related to gastrointestinal symptoms is ranked first, which is interesting given that escitalopram is known to have superior tolerability, and the patient is noting significant gastrointestinal symptoms at baseline- a situation in which an SSRI with a more favorable tolerability profile would be preferred. In the second example, for duloxetine, hypochondriasis is included, which may be consistent with duloxetine’s known benefit for somatic symptoms [15].

8.1 Graphs page, with example for PHQ-9

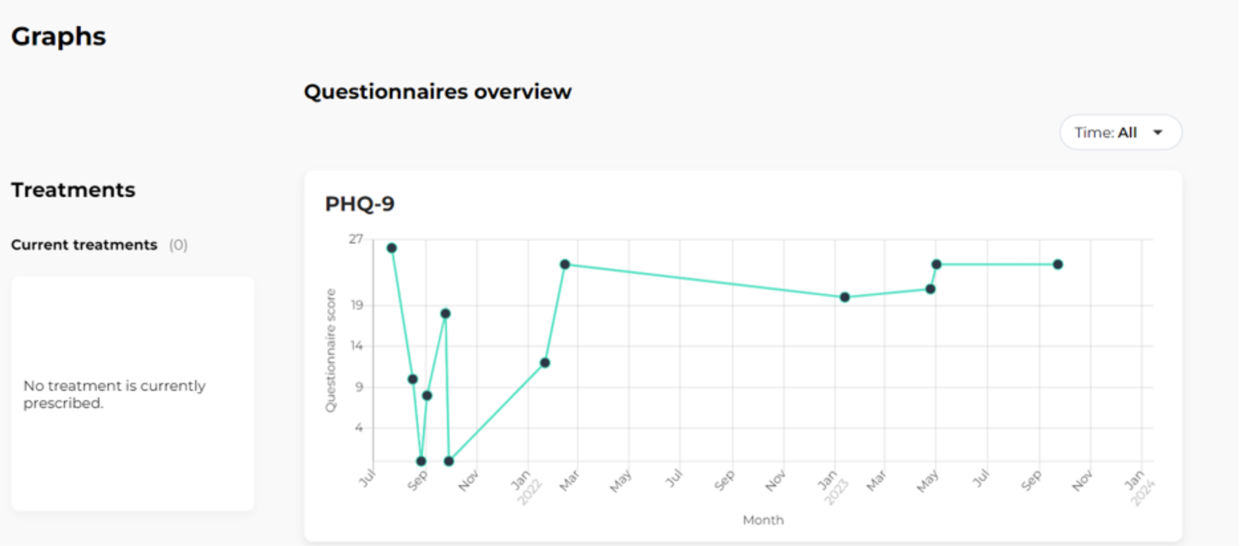

8.2- Example of the guideline-based information provided to clinicians

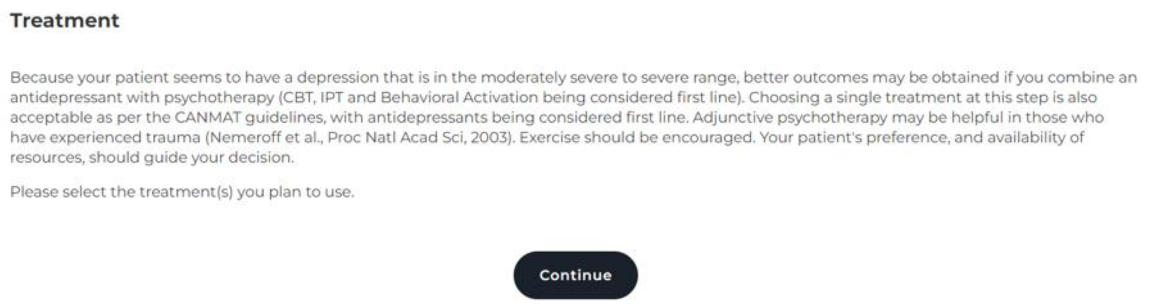

8.3 - Example result for A) escitalopram and B) duloxetine predictions. Information displayed includes drug class, typical dose, effective dose range, minimum/maximum dose range, dosing tips, clinical pearls, patient-specific remission probability for each drug, patient-specific remission probability across treatments, and population baseline.

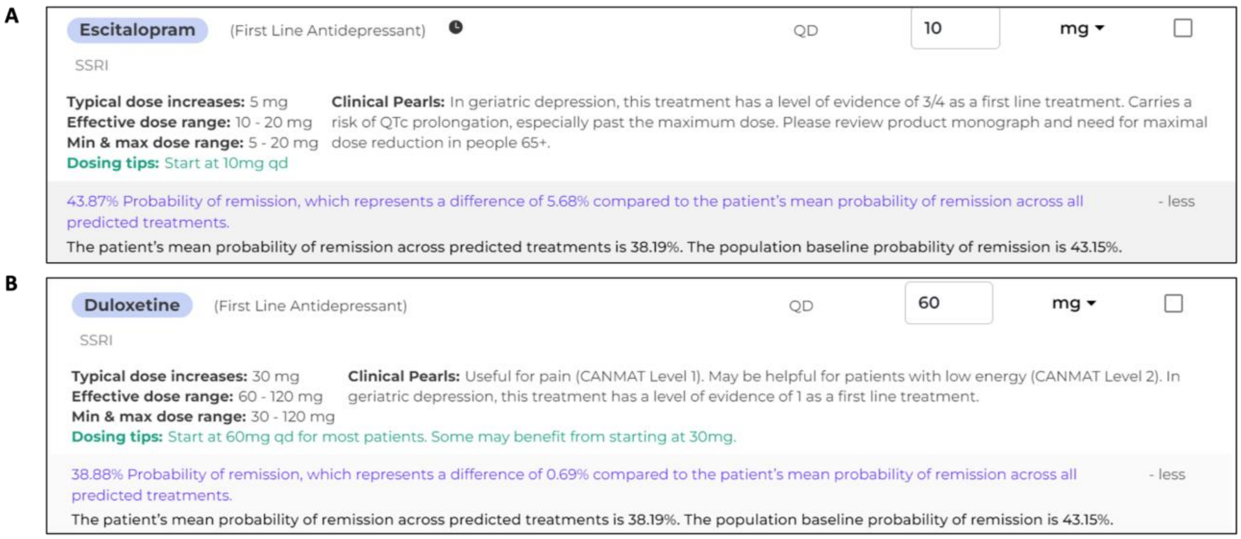

8.4- Example interpretability report for A) Escitalopram and B) Duloxetine that provides the top 5 most salient variables for each treatment prediction (note that Duloxetine is an SNRI; this was a typographical error in the application which was to be corrected in a later release).

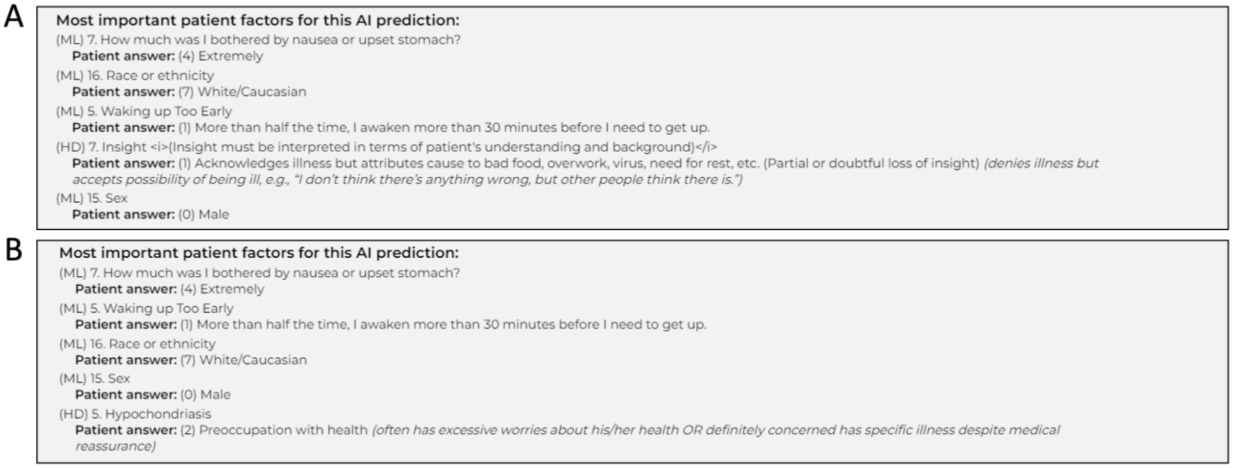

9- Total number of visits and unscheduled visits per group

In order to assess total resource usage, investigators provide an accounting of visits per patient, including unscheduled visits. There were more unscheduled visits in the Active-Control group numerically.

| Summary | Completing outpatient visits | Completing V1 | Completing V2 | Completing V3 | Completing V4 | Completing V5 | total missed visits | unscheduled visits |
|---|---|---|---|---|---|---|---|---|
| **42 active patients** | 204 | 42 | 41 | 42 | 38 | 37 | 10 | 4 |
| **19 control patients** | 103 | 19 | 19 | 19 | 19 | 19 | 0 | 8 |
| **Total** | 307 | 61 | 60 | 61 | 57 | 56 | 10 | 12 |
| **Active** | | | | | | | | |
| Total visits per patient | | | | | 4.86 | | | |
| Total unscheduled visits per patient | | | | | 0.095 | | | |
| **Control** | | | | | | | | |
| Total visits per patient | | | | | 5.42 | | | |
| Total unscheduled visits per patient | | | | | 0.63 | | | |

Exploratory analysis: PHQ-9 and GAD-7

Note that PHQ-9 and GAD-7 score analysis is limited as patients in both groups completed them at numerically different rates (see completion rate analysis), and patients completed questionnaires when they chose to.

PHQ-9

Baseline (first) PHQ-9 scores in the Active group were (mean = 11.7619, SD = 6.1676) and in the Active-Control group were (mean = 14.1053, SD = 4.6773); this was not significantly different (t = -1.6338, p = 0.11, t-test).
*Remitters and responders on the PHQ-9 for last score in the analysis set*

Last PHQ-9 scores in the Active group were (mean = 13.8333, SD = 5.6823) and in the Active-Control group were (mean = 11.8421, SD = 5.4392); this was not significantly different (t = 1.3057, p = 0.1999, t-test).

| **PHQ-9 t-tests by group - last visit** | | | | | |
|---|---|---|---|---|---|
| **Mean last score comparisons by group** | | | | | |
| T stat | p-value | Mean in Active | SD in Active | Mean in Control | SD in Control |
| 1.3057 | 0.1999 | 13.8333 | 5.6823 | 11.8421 | 5.4392 |

5 patients (11.90%) went into remission based on the PHQ-9 (Remission = last score < 5) in the Active group, and 3 patients in the Active-Control group (15.79%). This was not statistically significant:

| Remitters (n = 61) | | |
|---|---|---|
| | Remission | Not Remission |
| Active | 5 | 37 |
| Active-Control | 3 | 16 |
| **Fisher's exact test** | | |
| p-value = 0.70<br>95 percent confidence interval: (0.1230731 5.228974)<br>Odds ratio: 0.7248 | | |

Furthermore, 6 patients (14.29%) responded based on the PHQ-9 (Responders have a (first score - last score)/first score > = 0.5) in the Active group, and 5 patients in the Active-Control group (26.32%). This was not statistically significant:

| Responders | | |
|---|---|---|
| | Responder | Not Responder |
| Active | 6 | 36 |
| Active-Control | 5 | 14 |
| **Fisher's exact test** | | |
| p-value = 0.29<br>95 percent confidence interval: (0.1008697 2.2960105)<br>Odds ratio: 0.4731 | | |

Finally, change from baseline to last score in PHQ-9 were (mean = -2.0714, SD = 8.0255) in the Active group and (mean = 2.2632, SD = 6.0813) in the Active-Control group. This was statistically significant, indicating that the Active-Control group showed more improvement compared to the Active group (p = 0.02, t-test). The difference in rate of change (mean = -0.1637 , SD = 0.7826) in the Active group and (mean = 0.1908, SD = 0.5064) in the Active-Control group over time was statistically significant (p = 0.04, t-test).

| t-tests by group with at least 2 visits |
|---|

| Score change from baseline by group (baseline - last score) | | | | | |
|---|---|---|---|---|---|
| T stat | p-value | Mean score change from baseline in Active (n = 42) | SD of score change from baseline in Active (n = 42) | Mean score change from baseline in Active-Control (n = 19) | SD of score change from baseline in Active-Control (n = 19) |
| -2.3236 | 0.02 | -2.0714 | 8.0255 | 2.2632 | 6.0813 |
| **Rate of change from baseline by group (change from baseline / # of weeks from first-last visit)** | | | | | |
| T stat | p-value | Mean rate of change in Active (n = 42) | SD of rate change in Active (n = 42) | Mean rate of change in Active-Control (n = 19) | SD of rate of change percent in Active (n = 19) |
| -2.1154 | 0.04 | -0.1637 | 0.7826 | 0.1908 | 0.5064 |

GAD-7

Baseline (first) GAD-7 scores in the Active group were (mean = 8.5714, SD = 5.3472) and in the Active-Control group were (mean = 10.6842, SD = 4.6074); this was not significantly different (t = -1.5756, p = 0.12, t-test).

*Remitters and responders on the GAD-7 for last score in the analysis set*

Last GAD-7 scores in the Active group were (mean = 9.2381, SD = 6.9064) and in the Active-Control group were (mean = 8.2105, SD = 5.9496); this was not significantly different (t = 0.5934, p = 0.5562, t-test).

| GAD-7 t-tests by group - last visit | | | | | |
|---|---|---|---|---|---|
| **Mean last score comparisons by group** | | | | | |
| T stat | p-value | Mean in Active | SD in Active | Mean in Control | SD in Control |
| 0.5934 | 0.5562 | 9.2381 | 6.9064 | 8.2105 | 5.9496 |

31 patients (73.81%) went into remission based on the GAD-7 (Remission = last score < 5) in the Active group, and 13 patients in the Active-Control group (68.42%). This was not statistically significant (p = 0.76, Fisher’s exact). This indicates that both groups had similar improvement in anxiety.

| Remitters (n = 61) | | |
|---|---|---|
| | Remission | Not Remission |
| Active | 31 | 11 |
| Active-Control | 13 | 6 |
| **Fisher's exact test** | | |
| p-value = 0.76<br>95 percent confidence interval: (0.321808, 4.873295)<br>Odds ratio: 1.2949 | | |

Furthermore, 13 patients (30.95%) responded based on the GAD-7 (Responders have a (first score - last score)/first score > = 0.5) in the Active group, and 8 patients in the Active-Control group (47.37%). This was not statistically significant (p = 0.40, Fisher's exact). The low response rate, compared to the remission rate, is due to the low initial scores for anxiety, compared to depression.

| Responders | | |
|---|---|---|
| | Responder | Not Responder |
| Active | 13 | 29 |
| Active-Control | 8 | 11 |
| **Fisher's exact test** | | |
| p-value = 0.40<br>95 percent confidence interval: (0.1758574, 2.2310446)<br>Odds ratio: 0.6214 | | |

Finally, change from baseline to last score in GAD-7 were (mean = -0.6667, SD = 9.0059) in the Active group and (mean = 2.4737, SD = 8.6244) in the Active-Control group. This was not statistically significant (p = 0.14, t-test). The difference in rate of change (mean = -0.1617 , SD = 1.0663) in the Active group and (mean = 0.2193 , SD = 0.5881) in the Active-Control group over time was not statistically significant (p = 0.08, t-test).

| t-tests by group with at least 2 visits |
|---|

| Score change from baseline by group (baseline - last score) | | | | | |
|---|---|---|---|---|---|
| T stat | p-value | Mean score change from baseline in Active (n = 42) | SD of score change from baseline in Active (n = 42) | Mean score change from baseline in Active-Control (n = 19) | SD of score change from baseline in Active-Control (n = 19) |
| -1.4861 | 0.14 | -0.6667 | 9.0059 | 2.4737 | 8.6244 |
| **Rate of change from baseline by group (change from baseline / # of weeks from first-last visit)** | | | | | |
| T stat | p-value | Mean rate of change in Active (n = 42) | SD of rate change in Active (n = 42) | Mean rate of change in Active-Control (n = 19) | SD of rate of change percent in Active (n = 19) |
| -1.7906 | 0.08 | -0.1617 | 1.0663 | 0.2193 | 0.5881 |

It should be noted that the PHQ-9 and GAD-7 were collected weekly, and as such may demonstrate more heterogeneity, and represent more of an immediate state measure, than the MADRS and QIDS-SR-16 scores, which served as a more thorough review of overall symptoms.

9. STAR-P and STAR-C results (therapeutic alliance)

The standard STAR scales (STAR-C for clinicians, and STAR-P for patients) were used to measure the clinician-patient therapeutic alliances [52].
Subscores for both questionnaires were computed. For STAR-C, the subscores were 1) positive collaboration (items 1, 2, 5, 7, 10, 12); 2) emotional difficulties (items 4, 6, 9); and 3) positive clinician input (items 3, 8, 11) [52]. Comparisons were carried out using t-tests.
For STAR-P, the subscores were 1) positive collaboration (items 2, 3, 5, 6, 8, 11); 2) positive clinician input (items 1, 10, 12); and 3) non-supportive clinician input (items 4, 7, 9) [52]. In an exploratory analysis, two-tailed t-tests were computed for each subscore and total score for both questionnaires. No significant differences were found in total score (p = 0.13, Active = 40.88 ± 4.59, Active-Control = 43.26 ± 5.28) or subscore items for STAR-C (positive clinician input: p = 0.85, Active = 10.38 ± 1.53, Active-Control 10.47 ± 1.50; positive collaboration: p = 0.13, Active = 20.28 ± 2.91, Active-Control = 21.68 ± 3.09) with the exception of the emotional difficulties subscore (p = 0.03, Active = 10.23 ± 1.58, Active-Control = 11.11 ± 0.99). Examining individual items within this subscore, responses to the question “It is difficult for me to empathize with or relate to my patient’s problems” was significantly different between groups: p = 0.04 (non-corrected p-val). Active group mean = 2.85, Active-Control group mean = 3.37. However this was again by less than a point, and this exploratory analysis ignores within-clinician clustering of patients. No significant differences were found in overall score (p = 0.12, Active = 43.32 ± 8.25, Active-Control = 39.35 ± 6.62) or subscore items (positive clinician input: p = 0.16, Active = 10.16 ± 2.73, Active-Control = 8.94 ± 2.30; positive collaboration: p = 0.21, Active = 22.32 ± 4.61, Active-Control = 20.56 ± 3.84; non-supportive clinician input: p = 0.23, Active = 10.84 ± 1.95, Active-Control = 10.06 ± 1.95) for STAR-P. Overall, there is no indication that the inclusion of the AI had a negative impact on therapeutic alliance.

Figure 9.1: STAR-C Subscores by Group

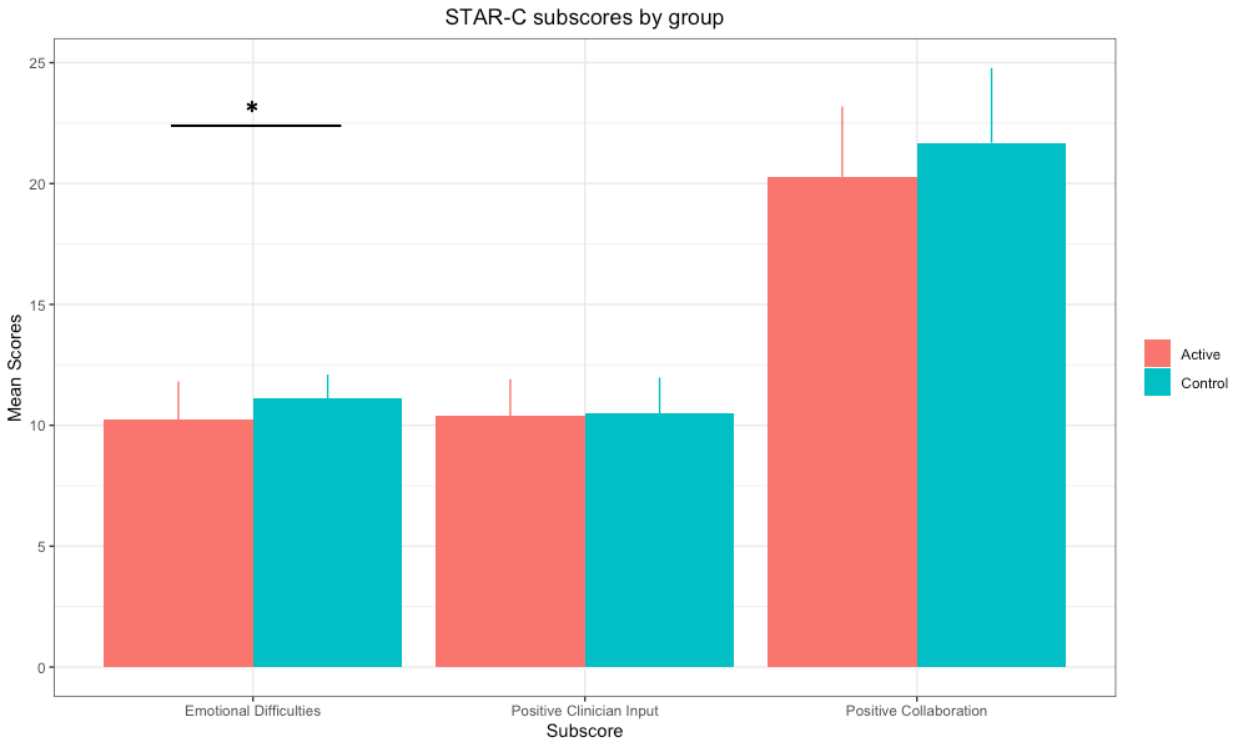

Fig. 9.1: STAR-C subscores by group - bar plots showing the means for each subscore by group, with the emotional difficulties subscore being significantly different between groups (emotional difficulties $p = 0.03$, positive clinician input $p = 0.85$, positive collaboration $p = 0.13$). Error bars represent the standard deviation.

Figure 9.2: STAR-P Subscores by Group

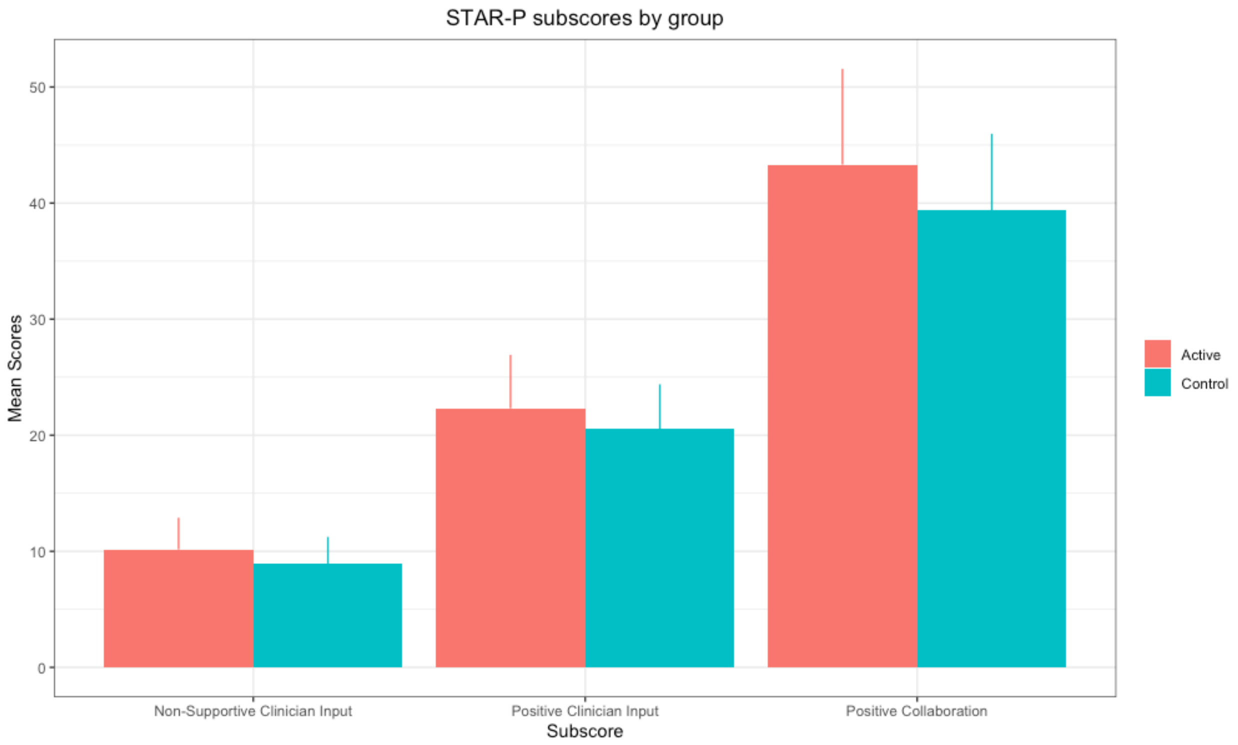

Fig. 9.2: STAR-P subscores by group - Bar plots showing the means for each subscore by group (non-supportive clinician input $p = 0.23$, positive clinician input $p = 0.16$, positive collaboration $p = 0.21$). Error bars represent the standard deviation.

Figure 9.3 STAR Total score by group

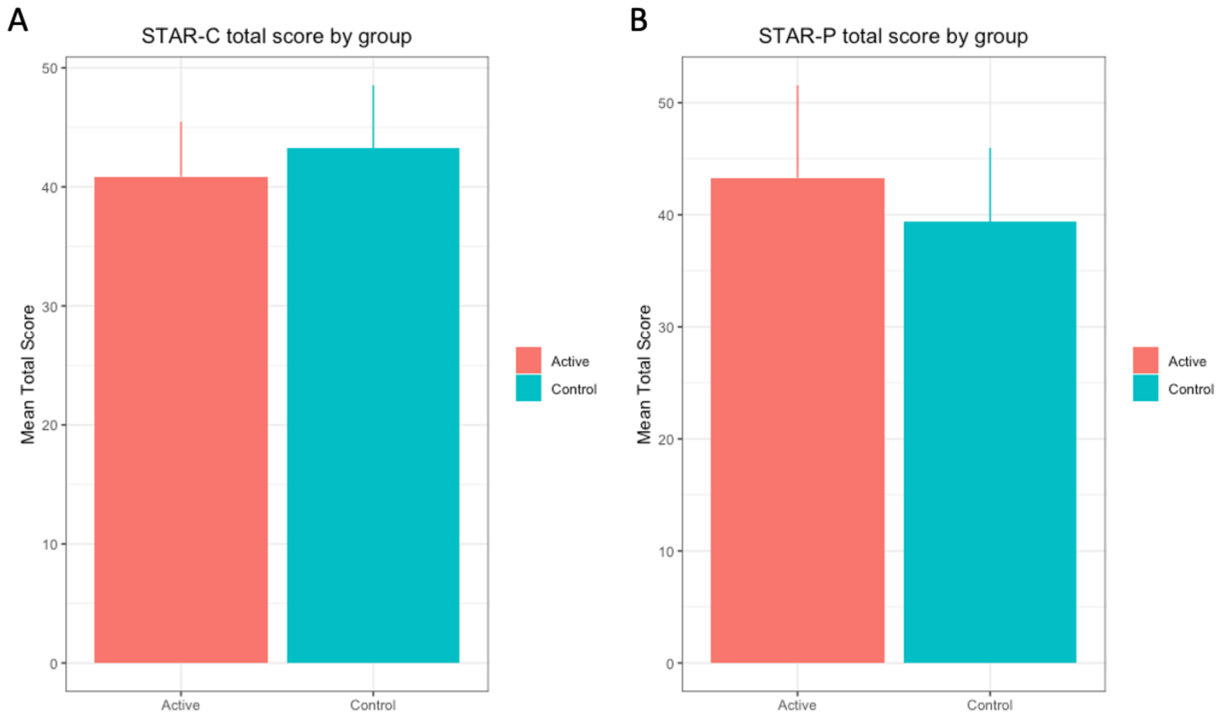

Figure 9.3: STAR Total score by group. Mean total scores by group for a) STAR-C (p = 0.13) and b) STAR-P (p = 0.12). Error bars represent standard deviation

### 10. Safety Outcomes and Adverse Event Narratives

*Safety Outcomes*

With respect to safety, adverse event rates (e.g., medication side effects) and serious adverse event rates (e.g., hospitalizations) were examined in the Safety population (all patients who completed at least the first treatment visit; this included 48 Active and 19 Active-Control patients). With respect to adverse events, 89 were reported for the Active group, a rate of 1.9 events per patient. 51 adverse events were reported in the Active-Control group, a rate of 2.7 events per patient. As such, the intervention was not associated with an increase in adverse event rate. There were 3 serious adverse events in the Active group, and none in the Active-Control group. All three events were determined to have been unrelated to the CDSS by the site's primary investigator. The three events included a visit to a psychiatric emergency department because of suicidal ideation; a brief hospitalization in a psychiatric short stay unit because of a panic attack and suicidal ideation; and a visit to general emergency department for a suicidal attempt gesture (overdose with 6 pills of 60mg duloxetine). All patients recovered from these events and none required prolonged hospitalization. As all three patients were known for cluster B personality traits or borderline personality disorder, the site PIs categorized these as expected events. Further details of the events and clinician perception of application safety can be found in the Supplementary Material.

*Adverse event narratives*

The first event was a visit to the ED because of suicidal ideation in the context of interpersonal and school stressors in a patient with depression and cluster B personality traits and a history of previous suicide attempts. The patient was sent to the ED by her clinician at the end of her first study visit. The second event was a patient who was experiencing a panic attack and suicidal thoughts; the site primary investigator requested that emergency services have him brought to the ED where he was briefly hospitalized in a psychiatric short stay unit. He was also known to have cluster B personality traits. In the report on the event, the patient noted being in a trial as a protective factor, as he did not want to hurt himself as it would prevent his data from helping others. These two patients were being seen in an emergency department followup clinic (i.e. they had been seen in the ED prior to starting followup with their study clinician and being recruited for the study) and as such were expected to have higher incidences of suicidal ideation and mood lability. The final patient was a patient with cluster B personality traits who is reported to have taken an overdose of her duloxetine as a suicide attempt or gesture (6 pills of 60 mg duloxetine); she was seen in a general hospital ED and discharged back to her outpatient psychiatric care, where she was noted by the treating clinician to have recovered from the incident rapidly.

11. Further details on avoiding contamination and expected effect size

Clinicians rather than patients were randomized as they were the ones receiving the decision support intervention, and to avoid contamination (Cook et al. 2016). This would occur if a clinician had patients in both the Active-Control and Active groups, but started using information gained in the Active group to treat patients in the Active-Control group. Given that the intervention essentially consists of information, the risk of contamination was judged to be high and therefore clinicians were randomized.

For expected effect size of 20% in remission rate difference between groups: in detail, investigators expected a 10% based on the AI treatment allocation (see [6,20–22] and 10% based on the fact that clinicians in the Active group will have access to a tool that provides measurement-based care and algorithm guided treatment (hence the hybrid nature of the tool reflected in the title); this 10% is based on previous studies of algorithm guided treatment (for example [49] and [50]). In these previous studies, fairly large differences between algorithm guided treatment and treatment as usual were found- roughly 30-40% differences in remission rate. Investigators estimated a reduced effect size of 10% because while the Active-Control group includes guideline training and access to measurement-based care assessments, it does not structure or force clinician decisions based on this.

12. Supplementary introduction

Investigators have developed and introduced the Aifred Clinical Decision Support System (hereinafter referred to as the CDSS). This is a digital platform which supports the implementation of guidelines (2016 CANMAT depression guidelines[15]), measurement-based care [15] in order to solve the treatment *management* problem, and which includes an AI (deep learning) powered module to assist in baseline treatment *selection* by providing predicted probabilities of remission for 10 commonly used first line antidepressants and combinations of these. For methodological details, see [6,20,21,31]. As representative a dataset as possible of the general adult population with moderate or greater severity MDD was obtained in order to assess potential model bias [38]. The choice to focus on first line treatments was made both due to availability of data needed to train the machine learning model and in order to maximize safety in this novel clinical application of AI. In the case of an incorrect model prediction (e.g. matching the patient to a sub-optimal first-line treatment) would still result in the patient obtaining care according to best practices, while a correct prediction could provide the patient with significant benefit by reducing the time required to achieve remission.

Prior to the current study, investigators performed extensive feasibility and ease of use testing of this CDSS in both simulation center and *in vivo* feasibility studies [16–19]. With *in silico* testing demonstrating that the AI component should help improve remission rates [6,20–22] and *in vivo* testing demonstrating that the platform was feasible, easy to use and likely safe[16–19] the current study was undertaken with the main objective of determining the effectiveness of the platform in improving depression treatment outcomes in patients with moderate to severe depression as well as to assess platform safety.

In a previous simulation center study, investigators demonstrated that clinicians could use the platform successfully in short (10 minute) interviews with standardized patients after less than half an hour of training and explanation [16,17]. Clinicians found the platform to be usable and trustworthy, and investigators found that they were more likely to prescribe in accordance to AI predictions when the patient was more severely depressed and when their trust in the model was higher. Trust was in turn related to how well they felt the interpretability report related to the patient in front of them. In a subsequent feasibility study in-clinic, investigators demonstrated that the platform did not lengthen appointment times; that it could be easily used both in-person and via telemedicine; and that it was rated as being feasible and clinically useful by the majority of patients and clinicians [19,53]. In addition, there were no serious adverse events related to the CDSS and the only adverse event noted was that one patient experienced transient anxiety while answering questionnaires.

### 13. Supplementary Methods

#### *Further details on patient recruitment criteria*

Psychiatric comorbidities (aside from bipolar disorder) were permitted as long as the clinician confirmed that there was an MDD they wished to focus on treating, and that depressive symptoms were not better explained by a comorbidity. Patients who did not have access to a computer or to the internet were offered a free computing device with internet connection for the duration of the study. It is also important to note that as diagnosis of MDD using the MINI requires the ruling out of non-psychiatric causes for depressive symptoms, referring clinicians were asked to rule out, in line with their best medical judgment, any non-psychiatric for depressive symptoms prior to referral to the study.

There were no exclusion or inclusion criteria related to input data for the AI model. When inputting data, patients and clinicians were required to fully complete the AI-related questionnaires before submitting (such that there was no missing data in any given AI-input). Patients and clinicians could also choose not to, or forget to respond to, these input questionnaires. In these cases the AI would not be able to display results, but the participants were not removed from the study given the desire to observe the impact of the CDSS in as naturalistic a setting as possible.

#### *Further details on setting*

Technical integration of the CDSS into trial settings was simple: the CDSS was hosted on an Azure server and was accessible via web browser or mobile phone application. As such, all that was required from sites was to allow the CDSS website to be accessed by clinicians from their computers; at most sites no information technology changes were necessary.

*Further details on measures*

Patients were also asked to complete a PHQ-9 and GAD-7 weekly once they had accounts on the CDSS as well as an additional Frequency, Intensity, Burden of Side Effects Rating (FIBSER) and Patient-Rated Inventory of Side Effects (PRISE-20) scales and a QIDS-SR-16 every other week until study end. Regular measures were intentionally administered more frequently than would normally be done in clinical practice in order to assess how much data patients were willing to provide; in a previous study with a similar schedule patients completed roughly ⅔ of the assigned weekly assessments [18]. At study end, they were asked to complete a second WHODAS as well as a customized end questionnaire asking them about their experience of the CDSS and in the study. Some patients were invited to participate in structured interviews with site staff (initially these were randomly selected, but near the end of the study all patients were invited as the study ended early).

Active clinicians were also asked to complete an end questionnaire about their experience in the study and of the CDSS as well as a structured end interview with site staff.

Patients were compensated for their time, and clinicians were compensated for their time where this was permitted by local site regulations.

*Further details on clinician training*

Clinicians were provided 1.5-2 hours of training by the medical monitor (DB) for this study. For those in the Active group, roughly 30 minutes were spent on explaining the AI model and learning to use the CDSS; the rest of the time was devoted to discussing the study protocol. While clinicians were not given information that would allow them to guess at the effectiveness of the AI model in order to preserve blinding, as described in investigators' companion paper on the AI model, clinicians were provided with training about the limitations of the AI.

*Further details on patient participation/Patient intervention*

Patient experiences differed only in their interaction with their clinicians, who had different information available based on their group assignment. However, these experiences were expected to be highly variable even within-group as clinicians, in order to maintain a naturalistic approach, were not required to use the information they were provided in a specific way with patients. Patients were asked to complete a number of baseline questionnaires as well as weekly sets of questionnaires (see below); however they were not required to complete their weekly questionnaires and were not removed from the study if they failed to do so. They received email reminders to complete their questionnaires. Patients often had access to the patient portal for up to 1-2 weeks prior to their first treatment visit (mean time 8.4 days), depending on clinician scheduling. Clinicians could see their patients more often than required, provided they alerted study staff so this could be recorded. Patients were not removed from the study if they needed to visit the emergency room or be hospitalized; rather, investigators attempted to complete the study as well as possible under these conditions.

*Further details on site recruitment efforts*

Significant efforts were made to recruit family-practice sites and family medicine units within participating sites. Unfortunately, the impacts of the COVID-19 pandemic and its aftermath reduced the capacity of family medicine units to participate.

It should be noted that it is likely that only those clinicians who desired to participate in clinical research and who had the time to participate would have been recruited into the study; white this may have some effect on result generalizability this is mitigated by the fact that in real-world practice only clinicians with the time and desire to adopt new technologies would be likely to use the AI-CDSS.

14. Further Supplementary results

*Further details on clinician recruitment and dropout*

50 clinicians were recruited, consistent with the recruitment target. Of these, 47 clinicians were trained and cleared to begin recruitment of patients. 3 clinicians were not able to begin recruitment due to early study termination. Of the 50 clinicians recruited, 26 were randomized in the Active group and 24 in the Active-Control group. 39 of these clinicians were psychiatrists (18 were randomized in the Active group and 21 in the Active-Control group); 2 were nurse practitioners specialized in psychiatry (1 was randomized in the Active group and 1 in the Active-Control group) and 9 were psychiatry residents (7 were included in the Active group and 2 in the Active-Control group). Of the 47 clinicians recruited who were able to recruit patients, 25 were randomized to the Active group and 22 to the Active-Control group. 27 clinicians recruited at least one patient (57%); 16 in the Active group (64%) and 11 in the Active-Control group (50%). 26 clinicians remained in the study until its termination (55%); 12 clinicians (25.5%), 4 from the Active group and 8 from the Active-Control group, withdrew from the study early (including one clinician who moved to a non-participating site), and all 9 residents (19%) reached the end of their rotation in the participating clinic during the study (note that they completed seeing any patients they had already recruited). Active and Active-Control clinicians spent the same mean number of months in the study (Active = 9.9 months; Active-Control 10.1 months). Active clinicians recruited more patients than Active-Control clinicians, with Active clinicians recruiting on average 2.2 patients/clinician and Active-Control clinicians recruiting 1.6 patients/clinician (including all patients consented).

*Further demographic data*

Participant demographics for the Analysis set are presented in Table 1. As can be seen, groups were similar with respect to age, sex, gender, racial and ethnic makeup, household income, education, psychiatric comorbidities, whether or not patients had recurrent depression, baseline MADRS, drug and alcohol use at baseline, and personality disorder screening at baseline. Given the importance of anxiety in predicting treatment outcomes in depression [54] investigators examined whether there were significant differences in the prevalence of generalized anxiety disorder between groups; no significant difference was found ($X^2$ = 0.7, p = 0.40). There were no differences in baseline MADRS severity: Active = 33, SD = 7.3; Active-Control = 30, SD = 5.8; F = 2.1, p = 0.15, ANOVA.

*Low frequency comorbidities and DAST-10/AUDIT scores*

| Low frequency MINI comorbidities |
|---|

| | | |
|---|---|---|
| - MDD with psychotic features current | 2 (4.8) | 0 (0) |
| - Obsessive compulsive disorder current | 2 (4.8) | 4 (21.1) |
| - Binge eating disorder current | 4 (9.5) | 2 (10.5) |
| - Antisocial personality disorder lifetime | 4 (9.5) | 0 (0) |
| AUDIT (Alcohol use) N(%) | n = 41 | n = 17 |
| Low risk (0-7 points) | 35(85.4) | 13(76.5) |
| Medium risk (8-15 points) | 4(9.7) | 3(17.6) |
| High risk (16-19 points) | 2(4.9) | 1(5.9) |
| Addiction likely (20+ points) | 0(0) | 0(0) |
| DAST-10 (Drug use) N(%) | n = 41 | n = 17 |
| 0 (no problems reported) | 13(31.7) | 4(23.5) |
| 1-2 (low level) | 22(53.7) | 11(64.7) |
| 3-5 (moderate level) | 3(7.3) | 2(11.8) |
| 6-8 (substantial level) | 3(7.3) | 0(0) |
| 9-10 (severe level) | 0(0) | 0(0) |

Demographics by Country are available but not presented as low sample sizes do not permit protection of patient confidentiality.

*Further information on patient recruitment and dropout*

Considering only patients who attended visit 1 (V1), 36/48 (75%) of Active patients completed all 12 weeks, and 18/19 (95%) Active-Control patients. completed visit 5 (V5); this was not statistically significant (p = 0.09, Fisher’s exact test).
The early termination of several patients was due to early study termination (3 active, 2 Active-Control). Removing these patients, 32/50 (64%) of Active and 18/20 (90%) of Active-Control patients completed the study when considering all patients who were eligible (a significant difference, p = 0.04, Fisher’s exact test), and 32/45 (71.1%) of Active and 18/18 (100%) of Active-Control patients completed the study when considering all patients who attended Visit 1 (p = 0.01, Fisher’s exact test). Overall, more patients in the Active than the Active-Control group

chose to leave the study (patient termination or dropout). Of these 14 Active patients, 5 dropped out of the study before visit 1 (35.7%). While detailed reasons for each of these dropouts is not available, reasons available included: a patient who explained they joined the study in order to have more rapid access to their clinician; a patient withdrew from the study due to inability to complete frequent study visits and questionnaires; a patient was not comfortable starting medication; a patient felt that questionnaires to fill were too invasive. It is important to note that, from the patient perspective, their experience in the Active and Active-Control groups were the same outside of their interactions with their clinicians during visits, which depended in turn on how the clinician chose to use the application. As such, it is difficult to interpret the numerical differences between groups as being a result of the intervention itself, though this remains possible. Other elements, such as between-site differences and variations in healthcare systems between the US and Canada, may be at play, especially as during randomization all clinicians at one of the VA sites were randomized to the Active group. 5 patients chose to withdraw or were lost to follow up in Canada (11.1% of the 45 patients in the safety population from Canada), and 10 patients chose to withdraw or were lost to follow up in the U.S. (45.5% of the 22 patients in the safety population from the U.S.); of these, 9/10 were from one Veterans Affairs site- where all the doctors happened to have been randomized to the Active group. As such, it is possible that country and site effects may account for some of the observed dropouts, rather than study group.

*Further details on medication use at and prior to baseline*

At baseline, groups had a similar burden of all prescribed medications (including both psychiatric and non psychiatric medications). For 57 patients for whom data was available, Active patients had a mean of 3.2 medications prescribed and in the Active-Control group this was 3.1. With respect to the number of antidepressants tried prior to study start, Active patients had tried an average of 0.64 per patient; for the Active-Control group this was 0.28.

### 14. AI-Consistent Treatment Subgroup Analysis and discussion of potential mechanisms of action

*Subgroup analysis for patients receiving AI consistent vs. inconsistent treatment*

As pre-specified, investigators examined outcomes for patients, across groups, who did or did not receive treatment, at some point during the study, consistent with the predictions of the AI. AI consistent treatment was defined as having been prescribed an antidepressant or combination of antidepressants consistent with one of the top 3 predicted treatments for each patient, as ranked by the AI predictions. The top 3 treatments were chosen given that often the predictions could be within a few percent of each other, and so clinicians may have had valid reasons for not choosing the top first or second treatment which outweighed the small percent increase in remission it would have entailed. AI results were available for, or could be recovered based on completed questionnaires, for 56 patients (92% of the sample). Of these, 20 patients (35.7%) had AI consistent treatment (note that, due to complexity of prescribing in this study of patients in secondary and tertiary care settings, augmentation treatments or treatments for comorbidities were ignored when determining AI consistency unless these conflicted with the AI predictions). Of 38 Active patients with AI predictions available, 16 (42%) had AI consistent treatment. Interestingly, the rate at which Active clinicians chose AI-consistent treatments was close to the consistent treatment rate estimated in a previous simulation center study [16]. In addition, given that 74% of clinicians in the active group actually saw the AI results at the first or second visit, this represents a significant proportion of the choices made by active group clinicians. Of the 18 patients in the Active-Control group, 4 patients had AI consistent treatment

(22.2%). This suggests that the clinicians' used the AI results in making medication choices. Amongst those with AI consistent treatment, the remission rate was 20% (n = 4/20), similar to the rate in those with AI inconsistent treatment (n = 7/36, 19.4%). The response rate was 35% (n = 7/20) in the AI-consistent group, compared to 30.6% (n = 11/36) in the AI-inconsistent group- a 4.4% numerical difference. While this difference in response rate was not statistically significant ($X^2 = 0.11$, p = 0.73), it is important to note that the sample was likely underpowered to detect this difference. The results suggest a possible small benefit for patients receiving AI consistent treatment.

*Active group only*

Restricting the subgroup analysis to the Active group only (exploratory analysis), there were 16 of 38 patients who received AI-consistent treatment (42%). Of those receiving AI consistent treatment, 4 remitted (25%) compared to 7 of the 22 not receiving AI consistent treatment (31.8%); this was not significant ($X^2 = 0.21$, p = 0.65). With respect to responders, there were 6 responders in the AI consistent group (37.5%) and 8 in the AI not consistent group (36.4%); this was not significant ($X^2 = 0.01$; p = 0.94).

Given these subgroup results, which were underpowered given the sample size, there are several potential reasons for the improved outcomes in the Active group. While the subgroup analysis does not support that the improved outcomes were primarily due to optimized treatment selection using the AI, there may have been some advantage provided by the AI in terms of treatment selection, either in terms of optimal treatment or in terms of providing clinicians and patients more information about potential treatments which improved shared decision making[19]. As some patients may have had treatment changes prior to visit 1 it is possible that the CDSS had a more significant effect in terms of treatment management than treatment selection for some patients. It is also possible that the treatment selection which was focused on first line medications was less relevant for this secondary/tertiary care population, meaning that the AI and CDSS were more helpful in helping clinicians and patients consider options than to specifically match a patient to a treatment. Future models with more treatment options and more variation in first-rank treatments may be of further utility in this population. While the number of treatments and dose changes were similar between groups (see other sections of Supplementary Material), it is possible that the information provided by the AI or the clinical algorithm improved the effectiveness or timing of these changes. The increased rate of MADRS score improvement in the Active group does potentially speak to more effective treatment management by Active group clinicians during the study time period, despite the fact that Active-Control group clinicians had access to the same raw data, suggesting an effect of the CDSS platform. In addition, having access to the CDSS platform may have provided Active clinicians with superior organization and presentation of all of the patient's data, facilitating and improving the quality of their decision making. Further work is needed to explore these potential mechanisms. In addition, in future studies with larger sample size, different definitions of AI-consistent treatment could be explored, beyond the simple top 3 definition used here, which may not capture certain nuances clinicians employed when considering the AI results. Importantly, benefits were achieved without requiring data other than patient- and clinician-reported symptoms and demographics for the clinical algorithm and AI predictions, improving the immediate feasibility of the platform; in the future, the incorporation of more data types and relevant biomarkers may further improve the ability of this and similar systems to improve outcomes[7].

Analysis of AI model errors was not possible  due to the lack of mechanistic ground truth. For example, if the model predicted that a given molecule should have been effective for a patient and this was not the case, there is no clear mechanism we could point to which would explain why the patient did not respond to the treatment; the same is true in the case where a patient responded to a treatment the AI did not predict they would respond to. Addressing this would require further elucidation of mechanism underlying treatment response.

### 15. Supplemental discussion

It is important to discuss the potential generalizability of these results. Sites were diverse in nature and demographics show that patients were diverse in terms of their backgrounds and comorbidities. All treating clinicians were psychiatrists, psychiatry residents, or specialized nurse practitioners. This occurred despite best efforts to recruit primary care providers, which proved to be difficult due several factors (lack of embedded research staff, time-consuming clinic-level onboarding in busy clinical practices, and primary care service adaptation to the post COVID-19 environment). However, previous work has shown that the CDSS is feasible in primary care [19], [18], and AI training data included patients in both primary and specialized services [22]. Given that the majority of MDD is treated by primary care physicians, and that patients in primary care are more likely to have less treatment resistant or recurrent depression, future work will need to confirm similar if not improved results in primary care[27]. Indeed, previous work demonstrated that primary care clinicians found the CDSS to be more useful than psychiatrists did; given the complexity of MDD management, the CDSS could be a valuable tool in primary care [16,17]. In addition, given the lack of safety concerns identified in this and previous studies [18,19], it would seem reasonable to introduce the CDSS to primary care in future work. Finally, while the clinical algorithm based on the guidelines might be applicable across many jurisdictions, the AI model was trained on data mostly from European and North American populations and as such would need to be validated and potentially re-trained before being used outside of these populations given potentially different patterns of symptom expression in different cultures [28–30].

Further analyses based on this dataset will examine qualitative and quantitative data about clinician and patient perceptions of the platform. Similar methods as the ones presented here could potentially be used to assess clinical decision support in other disease areas.

It would be important to be able to identify which elements of the intervention are most responsible for the clinical improvements seen. This could have been accomplished by adding further arms to the trial, but this was not possible. To compensate, the design matched the Active and Active-Control interventions closely, such that the main differences between groups were the clinical algorithm and the AI predictions. The clinical algorithm, in turn, was approximated in the Active-Control group by the guideline training and questionnaire data provided to clinicians. The objective of the study was to determine the impact of the CDSS as a unitary intervention; future implementation research could focus on separating the platform's component parts in order to study their independent effects.

Efforts were underway to improve recruitment in the Active-Control group prior to the premature end of the study.
