## Supplementary material for "Artificial Intelligence in Depression – Medication Enhancement (AID-ME): A Cluster Randomized Trial of a Deep Learning Enabled Clinical Decision Support System for Personalized Depression Treatment Selection and Management": Table1

|  | Active group (n = 42) | Active-Control group (n = 19) |
| --- | --- | --- |
| Mean age (SD) | 44.0 (15.2) | 39.3 (12.4) |
| Sex - n female (%) | 20 (47.6) | 10 (52.6) |
| Mean baseline MADRS (SD) | 33 (7.3) | 30 (5.8) |
| Race | n = 40 | n = 18 |
| - White | 30 (75.0) | 13 (72.2) |
| - Other** | 10 (25.0) | 5 (27.8) |
| Mean yearly household income (USD) | n = 37 | n = 15 |
| - Mean (SD) | 42,206.87 (29103.3) | 44,817.10 (17975.4) |
| Highest level of education achieved* | n = 40 | n = 17 |
| - Some high school/ high school diploma or equivalent (GED) | 5 (15) | 5 (27.8) |
| - Some university or college | 12 (30.0) | 4 (22.2) |
| - Bachelor’s degree | 11 (27.5) | 6 (33.3) |
| - Graduate or Professional Degree | 7 (17.5) | 1 (5.6) |
| - Trade/technical training or other | 5 (12.5) | 1 (5.6) |
| Graduated high school | 39 (97.5) | 17 (94.4) |
| Currently employed | n = 39 | n = 18 |
|  | 18 (41.5) | 12 (66.7) |
| Marital Status |  |  |
| - Single | 22 (55.0) | 12 (66.7) |
| - Partnered | 18 (45.0) | 6 (33.3) |
| Mean number of medications (all indications) prescribed at baseline (n = 57) | 3.18 | 3.06 |
| Adverse Childhood Experiences (mean, (SD)) | 2.62 (2.59) | 3.61 (2.59) |
| MINI Comorbidities: n (%) |  |  |
| - Suicidality (current- past month) | 22 (53.5) | 13 (68.4) |
| - High suicidality score category | 15 (35.7) | 6 (31.6) |
| - Generalized anxiety disorder current | 15 (35.7) | 9 (47.4) |
| - Social anxiety disorder current | 11 (26.2) | 4 (21.1) |
| - Posttraumatic Stress Disorder current | 10 (23.8) | 1 (5.3) |
| - Panic Disorder current | 7 (16.7) | 2 (10.5) |
| - Alcohol Use Disorder Past 12 months | 6 (14.3) | 6 (31.6) |
| - Agoraphobia current | 5 (12) | 2 (10.5) |
| - Substance Use Disorder (Non-Alcohol), Past 12 months | 5 (11.9) | 2 (10.5) |
| SAPAS-SA (Personality disorder screening) N(%) | n = 41 | n = 17 |
| Those meeting cutoff score of 3 or more for positive screening | 25 (61) | 8 (47.1) |

*Table 1: Baseline Clinical and Demographic Characteristics per Group*

*Note: participants could select more than one option; the graduated high school entry was constructed based on the available data.

** Lower count rows have been collapsed into the ‘other’ category in order to preserve confidentiality
