## Supplementary material for "Artificial Intelligence in Depression – Medication Enhancement (AID-ME): A Cluster Randomized Trial of a Deep Learning Enabled Clinical Decision Support System for Personalized Depression Treatment Selection and Management": Table2

| *Outcome* | Active | Active-Control | p-value |
| --- | --- | --- | --- |
| Remission | 12 (28.6%) | 0 (0%) | 0.01 |
| Response | 17 (40.5%) | 3 (15.8%) | 0.06 |
| Mean change from baseline | 12 (SD = 13.5) | 4.9 (SD = 10.9) | 0.05 |
| Percent change from baseline | 35 (SD = 41.1) | 13.2 (36.2) | 0.05 |
| Slope of improvement (amount of change per week) | 1.26 (SD = 1.63) | 0.37 (SD = 0.91) | 0.03 |

*Table 2: Summary of Outcomes*
