## Supplementary material for "Artificial Intelligence in Depression – Medication Enhancement (AID-ME): A Cluster Randomized Trial of a Deep Learning Enabled Clinical Decision Support System for Personalized Depression Treatment Selection and Management": Table3

| **Visit** | **App Access (%)** | **Algorithm Access (%)** | **AI Result Access (%)** |
| --- | --- | --- | --- |
| **Treatment Visit 1 (Day 0)** | 0.88 | 0.69 | 0.67 |
| **Treatment Visit 2 (W2)** | 0.80 | 0.80 | 0.07 |
| **Treatment Visit 3 (W4-6)** | 0.74 | 0.74 | 0.00 |
| **Treatment Visit 4 (W8)** | 0.71 | 0.68 | 0.03 |
| **Treatment Visit 5 (W12)** | 0.56 | 0.47 | 0.00 |

*Table 3: Clinician Platform Usage by Visit* App access = proportion of clinicians in the active group who logged into the app at each visit; Algorithm Access = proportion of clinicians who went beyond logging in and access the clinical algorithm module; AI Result Access = proportion of clinicians who accessed the AI results at the end of one session of the clinical algorithm, which was intended to occur at Visit 1 or 2.
